# Dual foundation models for accelerometry predict future health

**DOI:** 10.64898/2026.07.24.26358894

**Authors:** Marcus Dige, Niels R. Lorenzen, Magnus Ruud Kjær, Angus Burns, Poul Jennum, Emmanuel H. During, James Zou, Emmanuel Mignot, Andreas Brink-Kjaer

## Abstract

Wrist accelerometers are ubiquitous and capture activity, sleep, and cardiorespiratory motion, but how this relates to future disease across the phenome is unclear. We encoded one week of UK Biobank accelerometry from 97,696 participants using two frozen self-supervised models and trained a multilabel survival model on these embeddings with participant age and sex for 390 outcomes. In 5,253 held-out participants, mean concordance was 0.688. A single component explained 76% of predicted risk variance and was associated with future disease burden and mortality. Nevertheless, disease-specific scores added discrimination beyond this shared axis for 85 of 101 well-powered outcomes. A higher-powered full-cohort out-of-fold analysis identified prodromal neurodegenerative signatures, strongest for Parkinson’s disease (five-year time-dependent AUROC 0.90; 428 cases), with limited attenuation after lead-time washout. Daytime movement contributed most, whereas sleep-related and genetic information contributed selectively. These findings establish one week of wrist movement as a scalable, low-cost representation of future health for wearable-based risk assessment.

## Introduction

Many diseases present with prodromal symptoms or even overt clinical manifestations years before formal diagnosis. These often manifest by changes in overall behaviour, sleep, or vitality that are difficult to ascertain during the short time span that encompasses a typical clinical visit. Wrist-worn wearable devices offer a low-burden option for long-term health data collection. Wrist accelerometry devices, for example, record movement and rest patterns over long periods of time and are worn by hundreds of millions of individuals [1]. The feasibility of characterising cardiorespiratory dynamics through wrist acceleration has further been demonstrated [2, 3, 4]. However, their clinical interpretation has largely been restricted to summaries such as activity volume, sleep duration, and circadian rhythmicity, or to models developed for individual diseases.

Evidence from population cohorts indicates that the relationship between wrist accelerometry and health is considerably broader. In UK Biobank, accelerometer-derived moderate-to-vigorous physical activity was associated with the future incidence of 373 of 697 evaluated conditions, with associations spanning all major disease categories [5]. Other analyses have linked individual dimensions of wrist recordings, including diurnal temperature rhythmicity and objectively measured sleep and rest-activity characteristics, to future disease across several organ systems [6, 7]. Together, these findings suggest that free-living wrist recordings contain information about a general state of health that extends beyond any single behaviour or clinical condition. However, such analyses examine a limited set of prespecified summaries separately. They therefore cannot determine whether the associations across diseases arise predominantly from one shared physiological-risk gradient, or whether the raw signal also contains distinct patterns associated with specific future diseases.

At the other end, disease-focused studies have shown that richer representations of wrist movement can identify clinically meaningful pre-diagnostic patterns. Wrist accelerometry has detected movement changes years before Parkinson’s disease diagnosis [8, 9, 10], contributed to models of incident Alzheimer’s disease [11], and revealed early movement signatures of knee osteoarthritis [12]. These studies demonstrate that wrist signals can contain disease-relevant information not captured by conventional activity summaries. However, because each study defines its own features, model, population and clinical outcome, the resulting signatures cannot readily be compared across diseases. It consequently remains unclear whether apparent disease prediction mostly reflects common correlates of poor health (such as reduced activity, disrupted sleep or frailty) or whether condition-specific information remains after this general health component is accounted for.

Self-supervised learning provides a means of addressing this question by learning reusable representations directly from large collections of unlabelled accelerometer recordings. Yuan et al. used the UK Biobank’s [13] roughly 700,000 person-days of accelerometer data as an unlabelled pretraining corpus for self-supervised learning, producing models that generalised across external datasets for human activity recognition [14] and enabled epidemiological studies of sleep [15]. Emerging foundation models such as the Pretrained Actigraphy Transformer used masked-autoencoder pretraining on activity data from the National Health and Nutrition Examination Survey (NHANES) [16] to predict mental-health outcomes [17]. More broadly, our recent work on a foundation model of multimodal polysomnography (PSG) showed that a common physiological representation can be systematically evaluated against future diseases across the phenome [18]. Yet accelerometry foundation models have primarily been evaluated on behavioural recognition, sleep phenotyping, or selected clinical endpoints. Their broader future-health content, and in particular the extent to which that content is shared across diseases or specific to individual conditions, has not been systematically characterised.

We previously developed two self-supervised foundation models of wrist accelerometry using about 109,000 UK Biobank recordings. The Human Activity (HA) model learns frequency-resolved representations of common free-living behaviours and achieved a balanced accuracy of approximately 0.85 in an external 24-hour video-annotated dataset [19]. AcceleRest was designed to represent sleep-related and cardiorespiratory information from wrist movement, which was validated against gold standard PSG labels of sleep stages with a macro F1 score of 0.69 [20]. Together, these encoders provide complementary representations of daytime behaviour and night-time physiology.

Here, rather than developing a separate specialised wearable model for each clinical condition, we used these frozen representations to ask how future-health information is organised across the phenome. We examined whether disease risks share a general wrist-derived physiological component, whether condition-specific information remains beyond that component, and which movement, sleep-related and genetic sources contribute to different disease predictions.

## Results

UK Biobank participants wore a research-grade wrist accelerometer (Axivity Ax3) for up to one week [13], which we encoded with two frozen self-supervised foundation models [19, 20]. From 97,696 participants (participant-disjoint train, validation and test split; Methods), a daily embedding was read by a rolling-window Cox proportional-hazards model with age and sex to estimate the time to first onset of each disease (Figure 1a; Methods). The primary metric was the held-out concordance index (C-index) for every outcome. Disease outcomes were phecodes with at least 0.30% prevalence plus all-cause mortality, a panel of 390 outcomes (388 with incident test cases); for rare outcomes we additionally report a full-cohort cross-validated out-of-fold readout as a secondary, higher-power estimate, labelled as such throughout.

**Figure 1:**
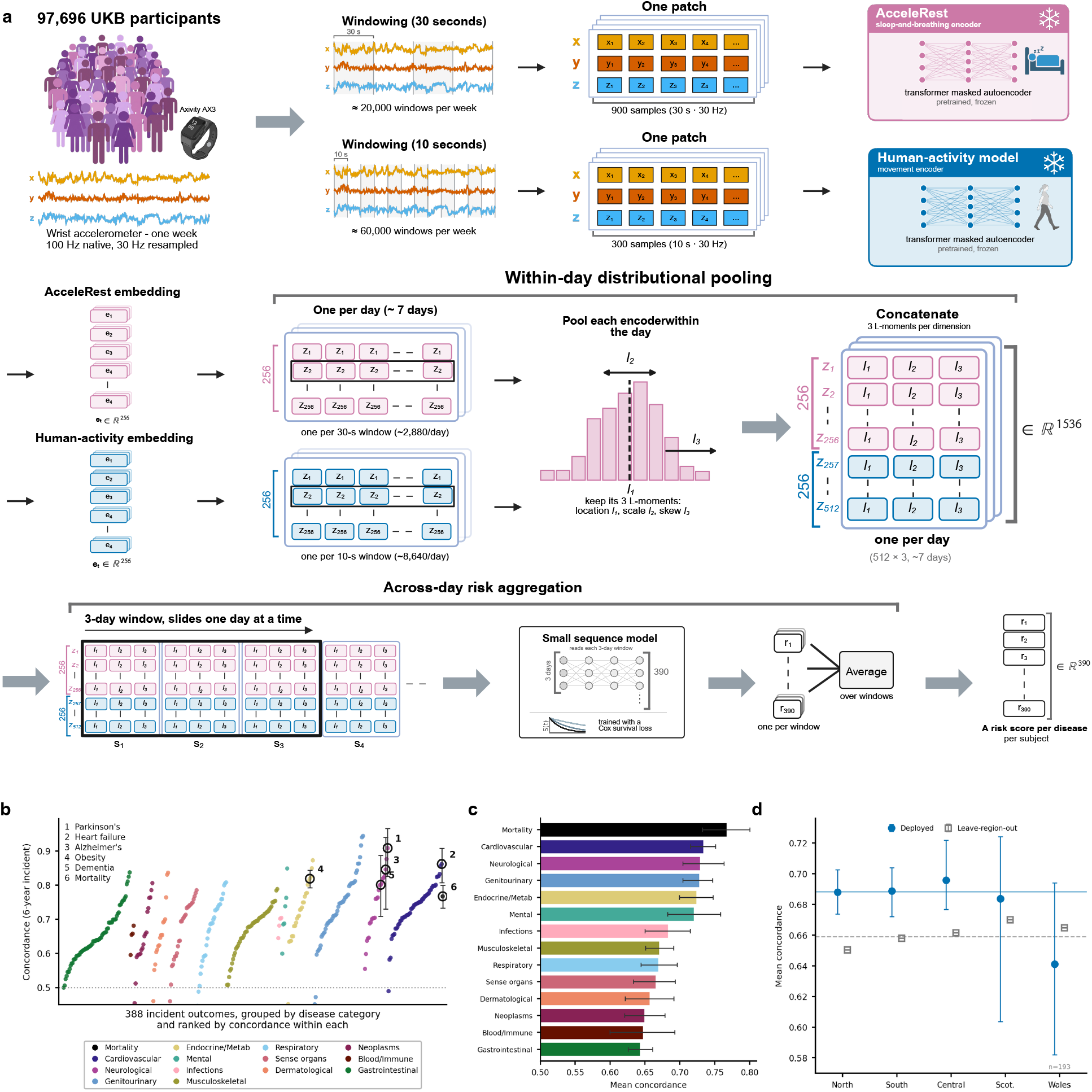
General-purpose wrist representations stratify incident disease risk within UK Biobank. **a**, Analysis overview: one week of wrist accelerometry is embedded by two frozen self-supervised encoders, aggregated within and across days, and read by a single survival model that outputs a six-year risk for every outcome. **b**, Per-outcome concordance coloured by disease category and ranked within category (the horizontal axis is an ordering); the dotted line marks chance, and six clinically important outcomes are ringed and numbered (key, top left). Points are point estimates; per-disease 95% confidence intervals are shown in Supplementary Table 6. **c**, Mean concordance within each disease category; every category ranks above chance. **d**, Transportability: mean concordance on each held-out UK Biobank region for the deployed model (filled circles) and under leave-one-region-out refitting (open squares), against the pooled reference (lines). Mean concordance across the panel is 0.688.

### General-purpose wrist representations embed future-health information

Wrist activity embedding predicted incident disease broadly (Figure 1b). Mean concordance over the 388 evaluable outcomes was 0.69 (95% CI 0.679–0.697), and every disease category ranked above chance, led by cardiovascular, neurological, and genitourinary diseases (0.73 for each category; Figure 1c). All-cause mortality, used as a non-specific indicator of general health, reached a concordance of 0.767 (95% CI 0.729–0.800).

We selected individual examples by both raw concordance and incremental value over a demographic baseline: including Parkinson’s disease with C-index of 0.91 (95% CI 0.83–0.97, Δ*C* = 0.144, 25 incident cases), Alzheimer’s disease with C-index of 0.85 (0.72–0.94, Δ*C* = 0.088, 15 cases), chronic obstructive pulmonary disease with C-index of 0.81 (95% CI 0.76–0.85, Δ*C* = 0.110, 92 cases), pulmonary embolism with C-index of 0.71 (95% CI 0.622–0.784, Δ*C* = 0.086, 38 cases), and acute kidney failure with C-index of 0.80 (95% CI 0.768–0.829, Δ*C* = 0.055, 136 cases). Cardiometabolic and sleep-related outcomes were also well predicted, including heart failure (C-index 0.86, 95% CI 0.81–0.91, 37 incident cases), sleep apnoea (0.81, 0.75–0.86, 57 cases), type 2 diabetes (0.78, 0.74–0.81, 159 cases) and atrial fibrillation (0.75, 0.72–0.78, 183 cases). Per-disease concordances with confidence intervals and published comparators are given in Supplementary Table 3.

The breadth of predictions was not a feature of a single population (Figure 1d): mean concordance was stable across the four large UK Biobank regions (0.68–0.70 versus 0.69 pooled) and held under leave-one-region-out refitting (0.65–0.67 versus 0.659), providing internal geographical replication but not external validation. The signal was also not an artefact of near-term detection: excluding incident cases diagnosed within two years of recording changed mean concordance only from 0.688 to 0.685, though 19 outcomes dropped by more than 0.05, for which this contribution cannot be excluded (Supplementary Table 7).

### The predictions define a Shared Actigraphic Risk Component and disease-specific signatures

Although our accelerometry model predicts disease across every organ system, per-disease risk scores were far from independent (Figure 2). Across 101 outcomes with at least 50 incident cases, the predicted six-year risks were strongly correlated across diseases (mean Spearman correlation 0.70, 95% CI 0.690–0.704). This structure was not an artefact of demographics or of comorbidity. Residualising the risk of each disease on age, age squared and sex changed mean pairwise correlation by only 0.007. Further, incident co-occurrence was low (mean phi coefficient 0.045) and explained only 10% of the correlation (Figure 2a), and predicted risks stayed correlated at 0.56 even among disease pairs that never co-occurred.

**Figure 2:**
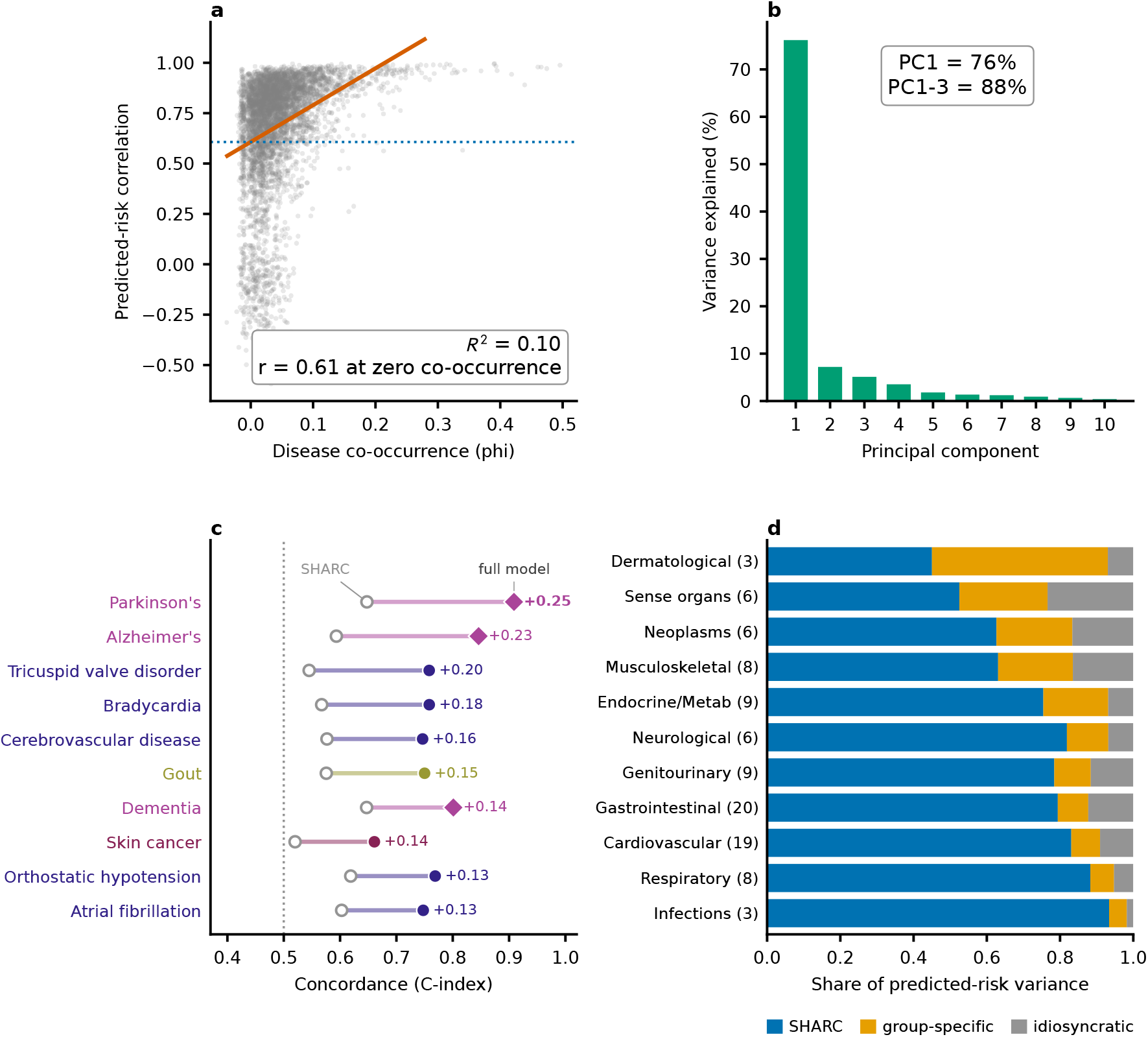
The predicted disease risks define a Shared Actigraphic Risk Component (SHARC) with disease-specific signatures. Data for 101 outcomes with at least 50 incident test cases (5,253 test participants). **a**, Each grey point is one disease pair: predicted-risk correlation (vertical) against incident co-occurrence (comorbidity, phi coefficient, horizontal). The solid line is the fit and the dotted line value at zero co-occurrence. Comorbidity explains only 10% of the correlation (*R*^2^ = 0.10) and the fit reaches 0.61 at zero co-occurrence, so pairs that rarely co-occur stay strongly correlated. **b**, Fraction of predicted-risk variance explained by each successive principal component: one component accounts for 76% (the first three, 88%). **c**, Per-disease discrimination beyond the SHARC for selected outcomes (bars, markers and labels coloured by disease category), ordered by the gain in concordance from the SHARC alone (open circles) and to the full wrist model (filled markers), joined by a bar labelled with the gain; the dotted line marks chance and diamonds mark neurodegenerative outcomes with fewer than 50 incident cases. For Parkinson’s disease the SHARC alone gives *C≈*0.65, yet the full model is far more discriminative. Related panels are in Extended Data Figure 8 and Supplementary Figure 5. **d**, For each organ system (a phecode category with at least three well-powered diseases), the share of standardised predicted-risk variance carried by the SHARC (blue), a single group-specific residual component (orange; the leading principal component of the within-system risk residuals once the SHARC is projected out) and an idiosyncratic remainder (grey; the residual disease-level variance captured by neither), so the three shares sum to one. Systems are ordered by their group-specific share.

Because predicted risks rise with age and differ by sex, we first residualised each disease’s predicted risk on age, age squared and sex, so that the shared structure would reflect variation beyond these demographics. The leading principal component of these standardised residuals explained 76% of their variance (three components, 88%; Figure 2b). Nearly every disease loaded positively onto it (97% positive). We term this the Shared Actigraphic Risk Component (SHARC). By construction, it is uncorrelated with age and sex, so it serves as an estimate of overall physiological risk independently of the factors. Participants with higher SHARC scores subsequently accumulated more incident diagnoses: mean future disease burden increased across score deciles (Spearman *r* = 0.22, 95% CI 0.20–0.25; Extended Data Figure 8). Because the SHARC is age-and sex-adjusted by construction, we tested whether it still tracked mortality within demographic strata. Within four strata defined by sex and median age, observed six-year all-cause mortality was higher in the upper half of the SHARC than in the lower half in every stratum (for example 4.2% versus 1.0% in older women and 6.9% versus 3.7% in older men), although deaths were sparse in the younger strata (12 among the younger women and 17 among the younger men; Supplementary Table 1). The SHARC tracked lower physical activity most strongly. Its sharpest association was with the 95th percentile of acceleration, a measure of the movement peaks (Spearman *r* = *−*0.52), and it held across mean acceleration, moderate-to-vigorous activity and the UK Biobank overall-acceleration average (Spearman *r* between *−*0.47 and *−*0.50). It was also associated with body-mass index (BMI) (Spearman *r* = 0.48, 95% CI 0.46–0.50) and sedentary behaviour (Spearman *r* = 0.31, 95% CI 0.29–0.34). These activity associations persisted after adjustment for body-mass index (partial Spearman *r* between *−*0.39 and *−*0.46; Extended Data Figure 10).

Importantly, the risk estimates carried predictive information beyond the SHARC. For each well-powered outcome, we added the predicted hazard of the outcome to the SHARC score and measured the out-of-fold gain in concordance. This gain was positive with a subject-bootstrap 95% confidence interval above zero for 85 of the 101 outcomes (84%; Figure 2c).

We made this organ-system structure explicit by partitioning each system’s predicted-risk variance into the SHARC, a single group-specific component (the dominant pattern of variation shared within that organ system once the SHARC is removed) and an idiosyncratic per-disease remainder (Figure 2d). The SHARC accounted for most of the variance in nearly every system (mean 73%, the exception being dermatological), while the group-specific component was largest for dermatological, sense-organ, neoplastic and musculoskeletal disease.

Individual outcomes could also gain markedly over the SHARC, including movement and neurodegenerative disorders, though these rest on few events. Specifically, Parkinson’s disease gained 0.25 in concordance (95% CI 0.188–0.312), Alzheimer’s disease, 0.23 (0.065–0.363), and dementia 0.14 (0.061–0.220) over the SHARC. Among the well-powered outcomes, atrial fibrillation gained 0.13 (0.098–0.168) (Figure 2c). The per-disease SHARC loadings are shown in Extended Data Figure 8. As such, the wrist accelerometer signal carries information beyond the SHARC (Extended Data Figure 9).

### The wrist representations encompass demographics and activity summary metrics

A central question was whether our wrist embeddings carry information beyond age, sex, BMI and activity summary metrics. The wrist embedding on its own reached a mean concordance of 0.684; our primary model added age and sex, giving the 0.688 reported above, and a jointly trained model that also included body-mass index reached 0.689. Age, sex and body-mass index together thus added only +0.005. The wrist embedding alone already outperformed an age, sex and body-mass-index baseline (*C* = 0.639) and a ridge model using the standard UK Biobank activity summaries (*C* = 0.577) (Figure 3a).

**Figure 3:**
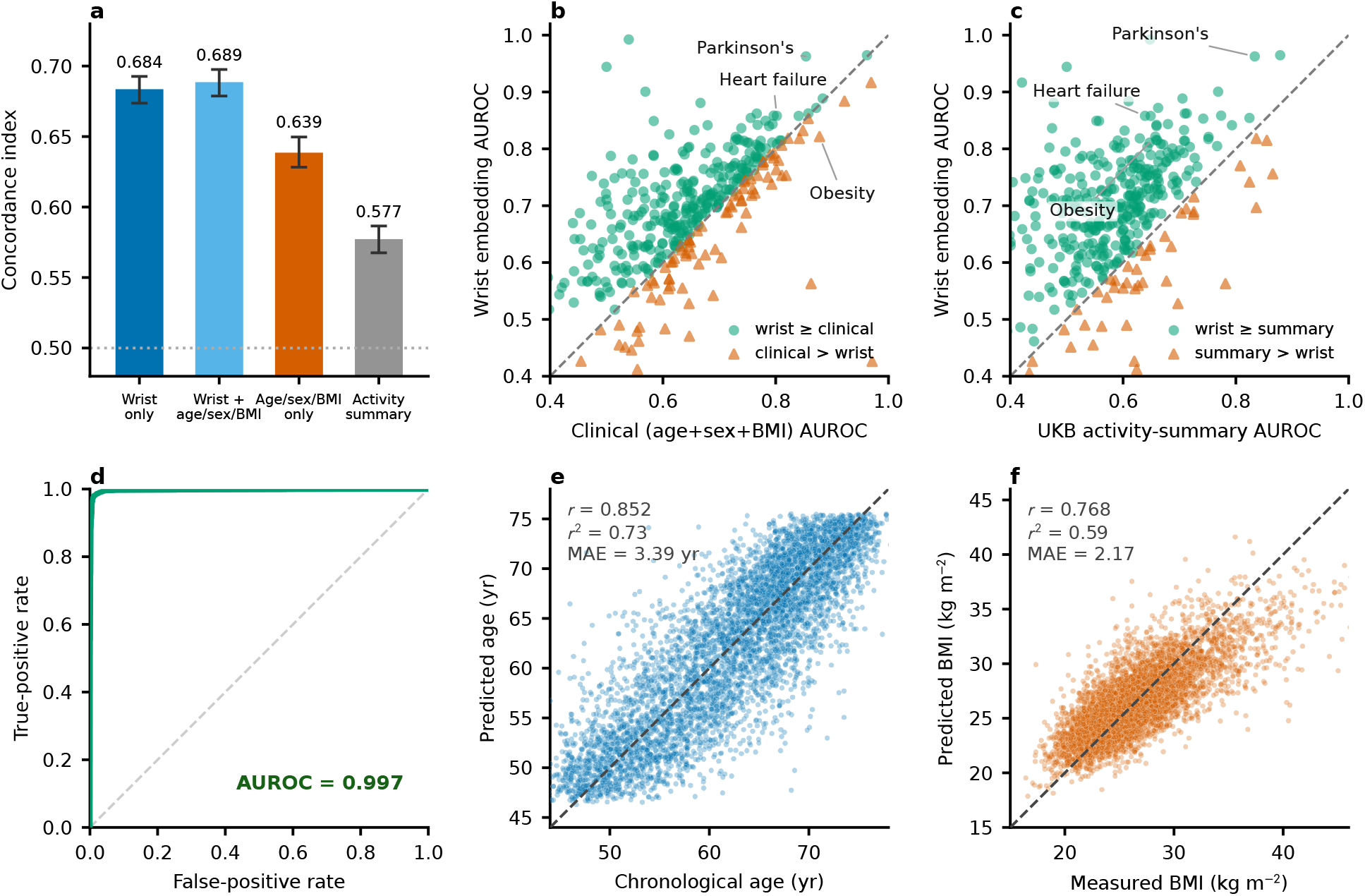
The wrist embedding already encodes age, sex and body-mass index and is the primary predictor. **a**, Mean concordance across the 388 evaluable outcomes for the wrist embedding alone (blue), the wrist embedding with age, sex and body-mass index added (cyan), an age, sex and body-mass-index clinical baseline with no wrist signal (orange), and a ridge model on the 100 UK Biobank activity-summary fields (grey). The wrist embedding alone exceeds the clinical baseline by 0.045 and the activity-summary baseline by 0.106, while adding the three demographic variables to the wrist model raises concordance by 0.005. Error bars are 95% confidence intervals from a bootstrap over the disease panel. Differences between models are assessed as paired contrasts, so the two wrist intervals overlap while their paired difference excludes zero (paired +0.005, 95% CI 0.001–0.009). The dotted line marks chance (concordance 0.5). **b**, Per-disease six-year incident AUROC of the wrist embedding versus an age, sex and body-mass-index baseline; the embedding’s AUROC exceeded the baseline’s for 78% of diseases (300 of 384, +0.053 on average). Points above the dashed diagonal favour the embedding; Parkinson’s disease and heart failure (above the diagonal) and obesity (below it) are labelled. **c**, The same per-disease comparison against a ridge Cox model built on the 100 accelerometer summary fields UK Biobank derives and distributes (Methods); the x-axis is that model’s six-year AUROC and the y-axis the wrist embedding’s. The embedding’s AUROC exceeded this baseline’s for 89% of diseases (340 of 384, +0.114 on average). **d**–**f**, The same wrist signal recovers the three demographics on the held-out test set: **d**, Receiver-operating characteristic for sex classification (AUROC 0.997); **e**, chronological age regression (Pearson *r* = 0.852, mean absolute error 3.39 years); and **f**, body-mass index (BMI) regression (Pearson *r* = 0.768, mean absolute error 2.17 kg m*^−^*^2^).

Wrist embeddings alone beat a linear age, sex, and BMI baseline on 78% of the 384 diseases with an estimable AUROC (300 of 384; mean 0.69 versus 0.64, +0.053 on average, 95% CI 0.044–0.063; Figure 3b, with the per-category breakdown in Supplementary Table 2). The same was true for a demographic model including quadratic and interaction terms. Here, the wrist hazard still raised per-disease concordance by +0.026 on average (near zero for a matched random control Extended Data Figure 1).

We further compared the model to commonly used activity summary metrics, including overall acceleration average and standard deviation, day-of-week and hour-of-day acceleration averages, and acceleration-intensity distribution [13] (the full 100-field list is in Supplementary Table 8). Wrist embeddings beat this 100-field summary baseline in 89% of diseases (340 of 384) and by +0.114 AUROC on average (95% CI 0.101–0.125) (Figure 3c). Adding these summary features to the accelerometry embeddings also did not increase the mean concordance. A feature-source comparison confirmed that the learned representation, rather than the pooling or the downstream model, carried the accessible signal (Extended Data Figure 4).

Even reduced to a single age-and sex-adjusted axis, the SHARC was as discriminative on average as the full 100-field activity panel (mean six-year AUROC 0.60 versus 0.58 across the 384 outcomes; a tie on the well-powered subset).

The limited gain from adding demographic information to the wrist models suggest that this could already be encoded in the wrist embeddings. Indeed, age, sex, and BMI could be linearly estimated from the frozen representation trained for disease prediction (*R*^2^ = 0.62, AUROC 0.99 and *R*^2^ = 0.47, respectively; see Supplementary Figure 1), with stronger performance obtained through dedicated models as reported in (Figure 3d–f).

### Genetic susceptibility complements wrist-derived risk estimates for selected diseases

We investigated whether genetic information distilled into polygenic risk scores (PRS) could improve disease prediction performance when combined with our accelerometer embeddings. We used the UK Biobank Standard PRS [21], computed from external genome-wide studies, i.e., they were not derived using UK Biobank data. For each disease with a matching score available (Supplementary Table 5), we added the PRS to the frozen wrist-embedding hazard and measured changes in c-index. This small model weights the polygenic score against the wrist model’s hazard. That hazard is an honest out-of-sample prediction only on the held-out test set. On the training data it is in-sample and over-optimistic, so a combiner fitted there would learn weights that are wrong for the honest hazard the model is judged on. We therefore fitted the combiner on the test set, with its weights learned by cross-validation on held-out folds of participants and applied to the rest, so that no participant’s own record set the weights that scored them. The primary analysis was restricted to participants of European genetic ancestry, where PRS are most portable.

Surprisingly, although the PRS were good predictors for some diseases, adding the PRS did not significantly increase predictive power for most diseases beyond the accelerometry model (Supplementary Table 4). Genetic information primarily improved prediction for cardiometabolic and autoimmune diseases. Atrial fibrillation prediction C-index rose by 0.036 (95% CI 0.011–0.060, 163 incident cases), type 2 diabetes by 0.022 (0.002– 0.040, 128 cases) and coronary artery disease by a similar margin, though only atrial fibrillation survived controlling for false discovery among well-powered outcomes. The largest per-disease increments in C-indices were for autoimmune diseases but rested on few events: coeliac disease increased from 0.59 to 0.87 (12 cases) and type 1 diabetes from 0.41 to 0.57 (11 cases). Autoimmune diseases had the largest mean gain across all 26 matched outcomes by +0.042 (95% CI +0.021 to +0.069; Extended Data Figure 3), falling to +0.027 once the two largest gains (coeliac disease and type 1 diabetes) were removed.

For Parkinson’s disease specifically, where the wrist model was a strong predictor, the PRS alone was weak (c-index of 0.54), and adding it did not improve concordance (*−*0.001). This also held for high-specificity screening. Here, adding the matched polygenic score did not sharpen screening for Parkinson’s disease at stringent operating points, and added little across the matched diseases (Supplementary Figure 3, Supplementary Table 11).

The predictive effects did not depend on ancestry. Repeating the analysis using all ancestries gave essentially the same mean gain (+0.046), indicating European restriction was conservative rather than decisive.

Genetics had to be matched to the disease. Adding the full panel of polygenic scores indiscriminately did not help. Feeding all 36 scores to every outcome reduced mean concordance by 0.040, likely because most scores are irrelevant to other diseases and add noise. Retraining the accelerometry model with the full PRS panel as joint inputs left overall mean concordance unchanged (0.6875 versus 0.6880 for the wrist alone), yet the same cardiometabolic outcomes still improved inside that model (type 2 diabetes +0.036, atrial fibrillation +0.033, heart failure +0.013). A single model cannot route each score to the disease it predicts, so the disease-matched PRS analysis is the main result (Supplementary Figure 2).

### Daytime activity drives most disease predictions

To see which signals drive each prediction, the held-out concordance of every outcome was decomposed into the contribution of each information source (Figure 4). We measured each contribution as the change in concordance when a wrist component was removed from the model, or when genetics or demographics were added to the wrist model. Within the wrist embeddings, daytime activity was the strongest predictor (mean contribution to concordance 0.021 across the phenome), ahead of night-time activity and the sleep-and-breathing information. The mixture was disease-specific: Parkinson’s disease and pulmonary embolism drew on the movement model, whereas chronic obstructive pulmonary disease, sleep apnoea and persistent atrial fibrillation relied on the sleep-and-breathing embedding, while heart failure was a notable exception that drew on night-time rather than daytime activity. Across the 332 outcomes with at least ten incident events, each wrist component reached significance far more often than the demographic covariates: daytime activity for 174, night-time activity for 122, night-time sleep-and-breathing for 91, and daytime sleep-and-breathing for 74.

**Figure 4:**
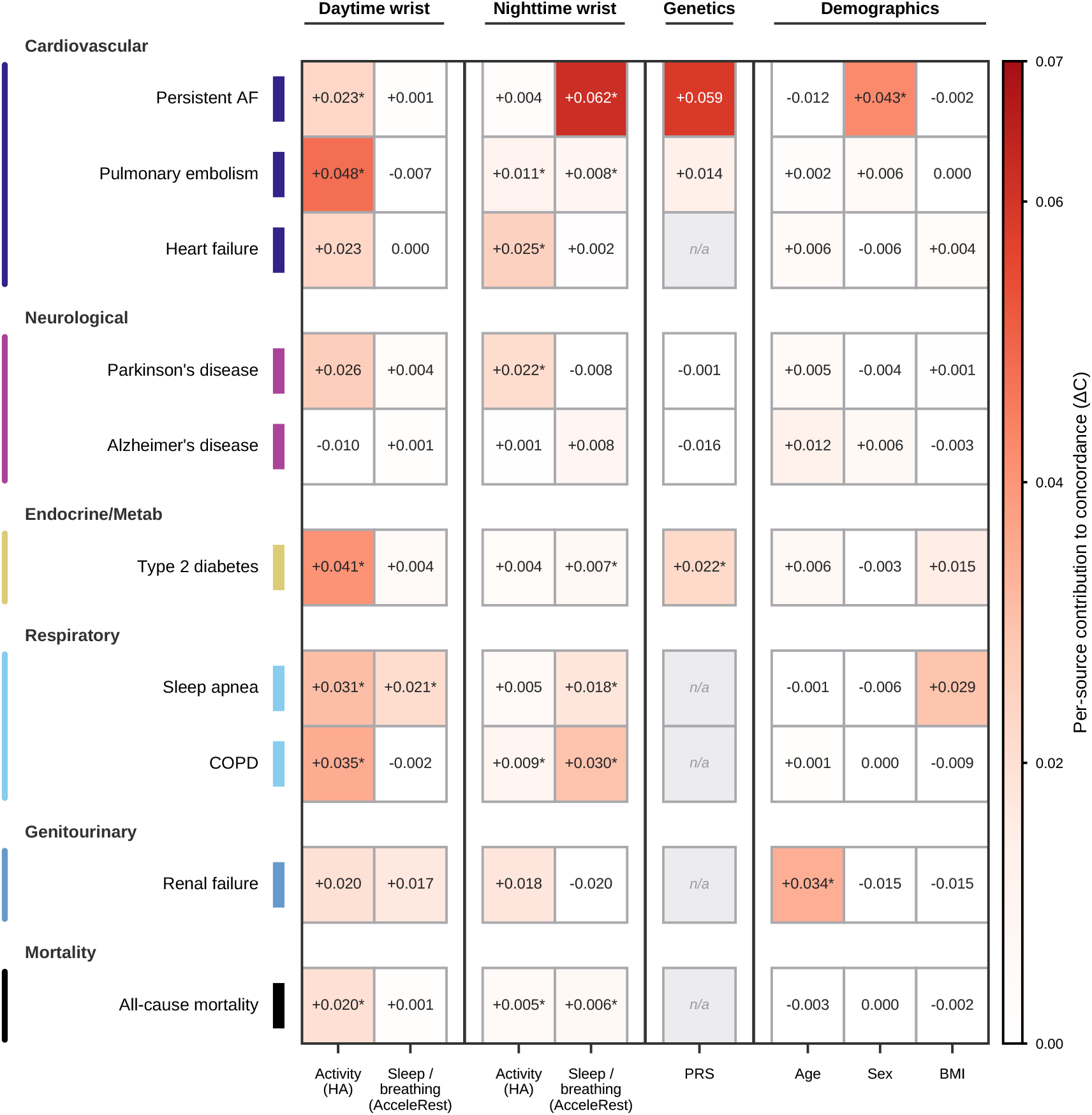
Per-disease source attribution: which signals drive each prediction. For ten diseases (rows, grouped and coloured by organ system), the contribution of each information source (columns) to held-out concordance. For the four wrist sources (the activity model (HA) and the sleep-and-breathing model (AcceleRest), each split into daytime and night-time by a fixed clock window, 09:00–21:00 and 21:00–09:00, with both encoders reading the full recording, day and night), the value is the drop in concordance when that component is removed; for genetics (a matched polygenic risk score, PRS) and demographics (age, sex and body-mass index), it is the increment in concordance when the source is added to the full wrist model. Colour encodes the contribution on a shared scale (white indicates none; values at or below zero indicate no contribution). Each cell prints the point estimate; an asterisk marks a statistically significant contribution, and n/a marks a disease with no matched polygenic score. Significance is defined per source. For the wrist components, the conditional-permutation attribution passes Benjamini–Hochberg false-discovery-rate control across the disease panel. For the polygenic score, the 95% confidence interval excludes zero. For age, sex and body-mass index, the contribution exceeds the 95th percentile of a matched covariate-permutation null (Methods). Genetics is evaluated on European-ancestry participants only. Per-source values for all outcomes are in Extended Data Figure 7.

In this unified attribution, genetics and the demographic covariates behaved as the dedicated analyses above indicated. The matched polygenic score contributed to cardiometabolic outcomes such as type 2 diabetes, and more broadly across the fuller genetically matched panel (Extended Data Figure 7), but added nothing for neurological outcomes beyond the wrist embedding. Age, sex and body-mass index each reached significance for only a minority of outcomes, spanning many organ systems. Surprisingly, BMI contributed more to musculoskeletal conditions than to the classic weight-linked metabolic diseases.

### The wrist embedding identifies prodromal neurodegeneration years before diagnosis

The strongest prodromal disease-specific signatures were found for neurodegenerative diseases, most notably Parkinson’s disease. On the held-out test set, Parkinson’s disease reached a concordance of 0.91 (25 incident cases), our primary estimate. Because these incident cases are few, we additionally quantified the prodromal signal with a cross-validated, out-of-fold risk estimate based on the frozen embedding across the entire cohort (428 incident cases), reporting discrimination on the entire at-risk population stratified by lead time as a secondary, higher-power estimate. The incident base rates were 0.44%, 0.34% and 0.38% for Parkinson’s disease, Alzheimer’s disease and dementia respectively. Encoders were trained in a self-supervised fashion on the UK Biobank. Thus, the disease labels, not the representation, were held out.

Among incident cases, median time from the wrist accelerometry recording to Parkinson’s-disease diagnosis was 4.9 years (IQR 2.8–6.7). On the full at-risk cohort (97,564 participants), the prediction reached a population time-dependent AUROC of 0.90 (95% CI 0.873–0.923) at five years and 0.87 (0.850–0.895) at seven years (Figure 5a,b). Excluding diagnoses within one, two or three years of recording reduced the seven-year AUROC only modestly, from 0.872 to 0.849 (Figure 5c). With the prodromal definition (diagnosis more than two years after recording), average precision (the area under the precision–recall curve) was 0.18 (95% CI 0.13–0.23) for the 282 incident cases. A model with no predictive skill would have an average precision equal to the outcome prevalence, 0.29%, so this corresponds to a 61-fold enrichment over chance. This pattern was consistent across lead-time washouts (Extended Data Figure 2). The same model could also be used to screen for Parkinson’s disease already present at the time of wear, the strongest signal for detecting prevalent disease in the phenome (cross-validated AUROC 0.916; Extended Data Figure 6). It reached an average precision of 0.64, corresponding to a 140-fold enrichment (Extended Data Figure 2).

**Figure 5:**
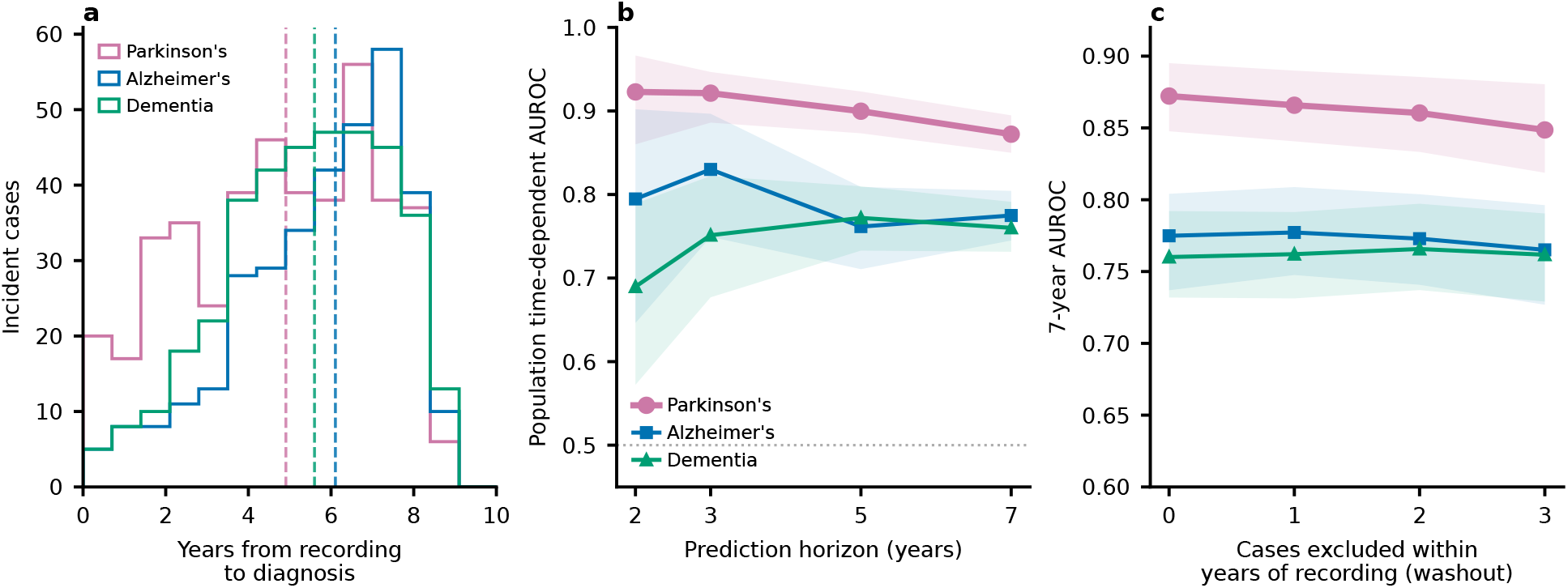
A prodromal neurodegeneration signature is detectable years before diagnosis. A secondary full-cohort cross-validated out-of-fold readout of the frozen wrist embedding was scored to predict future neurodegenerative disease across the UK Biobank cohort. **a**, Distribution of the time from wrist recording to diagnosis among incident cases, for Parkinson’s disease (428 cases, median 4.9 years), Alzheimer’s disease (333 cases, 6.1 years) and dementia (376 cases, 5.6 years); dashed lines mark the medians. **b**, Population time-dependent AUROC against the prediction horizon for the three outcomes; the dotted line marks chance (AUROC 0.5). **c**, Lead-time washout: seven-year AUROC after excluding cases diagnosed within up to three years of the recording, for each outcome. In **b** and **c**, shaded bands are 95% confidence intervals from subject-level bootstrap resampling.

The model also predicted other neurodegenerative disorders, albeit less strongly. On the same secondary cross-validated readout, the seven-year AUROC was 0.77 for Alzheimer’s disease and 0.76 for dementia, while the corresponding primary held-out concordances were 0.85 and 0.80. For both classes, discrimination was essentially unchanged under lead-time washout (Figure 5c), indicating these signals are also pre-diagnostic rather than near-onset disease markers. For participants who developed Parkinson’s disease, Alzheimer’s disease or dementia, the model preferentially over-ranked them for the other two conditions rather than for diseases of other systems, and this preference persisted after controlling for the SHARC, though these estimates rest on few events (Supplementary Table 10). The conditions carried by the model’s high-specificity false positives were clinically coherent and robust to comorbidity-burden adjustment (Supplementary Figure 4, Supplementary Table 9).

Across the 101 well-powered outcomes the predicted six-year risks were well calibrated (median calibration slope 1.03) and satisfied proportional hazards after false-discovery correction (Extended Data Figure 5a,b,e). Because the model’s baseline hazard was not retained, these curves assess calibration shape and should not be read as external validation of absolute risk. Building on this prodromal signal (Figure 5), flagging the top 10% by predicted risk captured 77% of incident Parkinson’s disease. In decision-curve analysis, the wrist readout gave greater net benefit than treating everyone, treating no one, or a demographic baseline across clinically relevant thresholds (Extended Data Figure 5c,d).

## Discussion

One week of free-living wrist accelerometry contained information about future disease across organ systems within UK Biobank. Three findings organise the study. First, a common representation stratified risk broadly, with a mean concordance of 0.688 across 388 evaluable outcomes. Second, most cross-disease variation was concentrated in one shared wrist-derived health-risk gradient, the SHARC. Third, disease-specific information remained beyond this gradient for most adequately powered outcomes, with the clearest example arising in prodromal neurodegeneration. These findings shift the interpretation of actigraphy from a collection of isolated activity and sleep summaries towards a representation containing both general and condition-specific health information.

In this work, we show that the combination of two frozen self-supervised actigraphy foundation models, one trained to infer gross movement patterns, the other to infer and amplify subtle respiratory signals during sleep, was able to jointly predict the incidence of a wide range of diseases across the phenome. The estimated risks of most diseases were found to lie along a dominant gradient of shared risk, predicting accumulation of morbidity and mortality. After removing the effects of age and sex on this dominant gradient we termed it the Shared Actigraphic Risk Component (SHARC) and propose that this be studied in more diverse populations (notably with a wider age range). Once validated, a component of this kind could in principle be read from any triaxial accelerometer and used as a guide for behavioural interventions, with the goal of lowering overall future morbidity. The SHARC correlated with several activity and sleep metrics, besides BMI and prevalent disease. Part of the predictive power likely reflects the model recognising complex activity and sleep patterns, captured complementarily by the two foundation models. A single-disease study could not have isolated this shared component and might have mistaken it for a disease-specific biomarker.

Despite the SHARC explaining a majority of the predicted risk for most diseases, most still had non-trivial group-level and idiosyncratic components. The strongest disease-specific signal was found for prodromal neurodegeneration. Most notably, our model flagged future Parkinson’s disease a median of 4.9 years before diagnosis, with a held-out concordance of 0.91 and a five-year time-dependent AUROC of 0.90 in the higher-powered out-of-fold analysis of 428 incident cases. Discrimination remained strong after excluding diagnoses occurring within three years of recording, supporting a persistent prodromal or risk signature rather than near-onset detection alone. This is biologically plausible because prodromal synucleinopathy affects multiple aspects of daily movement and sleep, including bradykinesia, gait, tremor and REM sleep behaviour disorder [22, 23].

These findings substantially extend the study by Schalkamp et al., which first established UK Biobank accelerometry as a strong predictor of future Parkinson’s disease [8]. Under a matched prodromal definition, our general-purpose representation achieved an average precision of 0.18 compared with 0.07 for the bespoke accelerometry model, and 0.64 compared with 0.14 for prevalent disease (Extended Data Figure 2). Thus, encoders not pretrained specifically for Parkinson’s disease, combined with an outcome-specific readout, captured a markedly stronger ranking signal than previous engineered approaches. This complements our parallel work showing that REM sleep behaviour disorder can be detected from wrist actigraphy across cohorts [9, 24] and that an actigraphy-derived RBD score predicts future Parkinson’s disease independently of, and synergistically with, polygenic risk [10]. Together, these studies position passive wrist accelerometry as a scalable means of enriching populations for prodromal Parkinson’s disease research and prevention trials. Notably, the night-time Parkinson’s signal arose primarily from the HA movement encoder rather than AcceleRest, suggesting that abnormal nocturnal movement was more informative than the sleep-and-cardiorespiratory component. Alzheimer’s disease and dementia showed weaker but persistent pre-diagnostic signals, potentially reflecting both shared neurodegenerative changes and overlap in diagnostic ascertainment.

Our per-disease prediction results (Supplementary Table 3) hold up well against other published wearable and accelerometer predictions. Beyond the prodromal Parkinson’s comparison above, our broad-spectrum model was competitive with models built for single health outcomes, matching or exceeding previous specific models for Alzheimer’s disease (C 0.85 vs. 0.68) [11], type 2 diabetes (C 0.78 vs. 0.77) [25], and one for all-cause mortality which included a full clinical panel besides accelerometry (C 0.77 vs. 0.77) [26], as well as a model for heart failure using more than 400 clinical biomarkers and spirometry variables (C 0.86 vs. 0.83) [27].

We tested the inclusion of genetic susceptibility for those outcomes where PRS were available and found that they contributed most to cardiometabolic and autoimmune diseases the accelerometry model predicted less well. This suggests some potential for future combined genetic-wearable screening with genetics providing a fixed susceptibility and accelerometry providing a momentary risk estimate.

We found that pooling the frozen embeddings from the two foundation models was a sufficient starting point for disease prediction. Using more complex architectures including sequence models for the prediction head did not improve the performance. The same was true for finetuning the pretrained encoders or adding hand-engineered features. This indicates that the relevant information for disease prediction is already captured in the representations of the frozen transformer models in a way that encompasses sequence information.

We showed that our dual-foundation model approach could be used to extract complementary information for disease prediction from the same wrist accelerometry signal with performance exceeding that of simple demographics or standard actigraphy summary metrics. While many modern consumer wearables include additional sensor modalities such as photoplethysmography (PPG) or ECG, they all include an accelerometer, and here we provide a direction to extract health information from the raw signal beyond standard activity metrics. Future studies may explore how these accelerometry models interact with physiological information derived from PPG or ECG. Here, we relied solely on the representations of previously developed models. Future studies could also explore ways to develop foundation models for accelerometry specifically with future disease prediction in mind.

This study includes several limitations. First, the lack of external cohorts to evaluate our model limits our claim to model generalisability to other devices and populations. Second, the performance for several outcomes, including Alzheimer’s disease and dementia, with relatively few incident cases, carried large confidence intervals. Third, outcomes were defined from hospital-inpatient and death records, which are under-ascertaining conditions managed only in primary care, for example dementia, psychiatric, metabolic and respiratory diseases. In the UK Biobank, adding primary-care records roughly doubles ascertained counts of dementia and Parkinson’s diseases; about half of dementia cases and 39% of stroke cases are recorded only in primary care [28, 29, 30]. Missed diagnoses enter the analysis as mislabelled non-cases and bias discrimination towards the null, so the true associations are likely stronger than reported. Fourth, the cohort is not population representative. The UK Biobank is known to suffer from ascertainment bias, being more highly educated and more healthy than the general population [31]. It lacks young, healthy-volunteers and the very old [32] in which the incidence of dementia roughly doubles every five years of age [33] and stroke incidence rises steeply [34]. Similarly, consent and wear-compliance requirements deplete prevalent dementia and disabling stroke, leaving recorded dementia here rarer than Parkinson’s disease.

Previously, in-clinic polysomnography has been shown to enable prediction of a similar wide array of diseases [18]. Others have also shown the power of molecular profiling of blood by metabolomics [35] or proteomics [36]. Importantly however, these other measures require a clinic visit or a laboratory assay. Here, we achieved this using a simple wearable accelerometer, a sensor type already worn by millions of consumers and which is deployable by simple letter mail. Our results suggest that simple wearable-based screenings could be used to enrich prevention trials with high-risk participants.

## Methods

### Study design, participants and outcomes

We conducted a retrospective prognostic modelling study using wrist-accelerometry and linked health record data from the UK Biobank [13, 37]. UK Biobank has general ethical approval for its studies from the NHS National Research Ethics Service (17 June 2011, reference 11/NW/0382), and all participants gave informed consent. This research was conducted under UK Biobank Application Number 62249. Between February 2013 and December 2015, UK Biobank invited participants to wear an Axivity AX3 research-grade triaxial accelerometer on the dominant wrist continuously for seven days, sampling at 100 Hz over a *±*8 g dynamic range [13]. Our analysis cohort comprised the participants with usable accelerometry that entered the disease model: 87,652 in training, 4,772 in validation, and 5,272 in the held-out test set (97,696 in total), 56% female, with a mean age of 62 years (s.d. 8) at the time of wear.

Hospital-inpatient ICD-10 diagnoses and causes of death were mapped to phecodes [38]. Phecodes meeting the prespecified prevalence threshold of 0.30% were retained (389 phecodes) and combined with all-cause mortality, yielding 390 outcomes. For each disease, participants with a diagnosis recorded on or before accelerometer wear were excluded from that disease’s incident risk set. An incident event was the first recorded diagnosis after wear. Two phecodes had no incident events in the held-out test set, leaving 388 evaluable outcomes.

Participants were assigned to mutually exclusive training, validation, and test sets before downstream disease-model development. The split contained 87,652 participants (96,016 recordings) in training, 4,772 (5,331 recordings) in validation, and 5,272 (7,460 recordings) in test. Of the 5,272 test participants, 5,253 had sufficient valid wear to generate a week-level embedding and formed the common evaluation cohort; 19 did not.

### Accelerometry processing and frozen representations

Recordings were resampled to 30 Hz and calibrated to local gravity [39] following the UK Biobank accelerometry protocol [13]. Non-wear was detected with a rolling standard-deviation criterion (10-second windows, a per-axis standard-deviation tolerance of 0.013 g, and 45-minute patience) and handled as a mask: the signal was preserved and propagated as missing through all downstream pooling, so that no non-wear epoch contributed to an embedding summary. Of the 115,398 recordings processed, 5,980 failed gravity calibration and 6 were unreadable, leaving 109,412 usable recordings. We did not apply a maximum-non-wear or minimum-wear-time threshold at the recording level; wear sufficiency was instead enforced during day-level pooling. A day contributed to a participant’s daily embedding only if it contained at least 100 valid (non-masked) patches in each encoder’s grid, about 50 minutes for AcceleRest (30-second patches) and about 17 minutes for the Human Activity model (10-second patches), and only days valid in both encoders were retained. Participants with fewer than three such valid days produced no week-level embedding and did not enter the analysis cohort (19 of the 5,272 test participants). Across the whole cohort, 108,807 of the 109,412 usable recordings met this requirement and produced a week-level embedding; these came from 97,696 participants, who formed the analysis cohort, and the remaining 605 recordings were excluded.

Two pretrained accelerometer foundation models were used as frozen encoders. The HA model is a transformer masked autoencoder over 10-second patches with a 50-minute context window, pretrained with a frequency-aware log-scale spectrogram reconstruction loss [19]. AcceleRest is a transformer masked autoencoder over 30-second patches with a 128-minute context window, pretrained with a respiratory-amplification reconstruction objective [20]. Each emits a 256-dimensional embedding per patch, with a one-patch sliding stride. Both encoders were pretrained by self-supervision on about 109,000 UK Biobank recordings and were not fine-tuned for this work.

We compared two ways of reducing each day’s sequence of window embeddings to a fixed vector: the per-day mean, and a distributional pooling that retains the first three sample L-moments (location, scale and the third L-moment, a measure of asymmetry) of each embedding dimension within the day [40]. Each encoder is reduced separately within the day, and the two daily summaries are concatenated. The distributional pooling is motivated by within-day variation being largely orthogonal to the daily mean, and is the primary model.

### Multilabel disease prediction model

The daily embedding sequence was passed to a rolling-window transformer over three-day windows [41], trained with a multilabel Cox proportional-hazards loss across the panel; predictions over all valid windows were averaged at inference [42]. The transformer had two pre-normalization layers, model dimension 256, four attention heads, feed-forward dimension 1,024, and dropout 0.1, with a learned per-day positional embedding and class-token pooling, and took the 1,536-dimensional daily vector as input. It was optimised with AdamW (learning rate 10*^−^*^4^, weight decay 10*^−^*^3^, batch size 64) under a cosine schedule for up to 100 epochs, with early stopping on the validation loss (patience 20). Four models with random seeds 42, 123, 456, and 789 were trained and used as an ensemble by averaging their hazards. The primary model concatenated participant age (in years) and sex (as a binary indicator) to the pooled representation before the output head; a wrist-embedding-only variant omits them.

### Primary evaluation and statistical analysis

The primary discrimination metric was Harrell’s concordance index (C-index) [43, 44], evaluated per participant over all observed follow-up. Six-year incident AUROC was used as a secondary fixed-horizon metric. For the conventional six-year AUROC analysis, participants censored before the horizon were excluded. We report mean C-index across the 388 evaluable outcomes, category-level summaries and per-outcome estimates with event counts and confidence intervals. Inferential emphasis was placed on the 101 outcomes with at least 50 incident test events; estimates based on fewer events were treated as exploratory.

### Cross-fitted additions to frozen predictions

When evaluating information added to a frozen disease risk score, we used event-stratified within-test cross-fitting [45]. A ridge-penalised Cox model combined the frozen score with the added covariate block and scored each participant using a model fitted without that participant. The number of folds was ten for outcomes with at least 50 events, five for at least 20 events and three for at least 10 events. The ridge penalty was max(0.1, 10*/n*_ev_). This procedure was used for demographic, activity-summary, polygenic-risk-score and SHARC-complement analyses. It was fitted in the held-out test cohort because the deep risk score was out of sample only there; a combiner fitted to in-sample training risks would estimate its weights against an over-optimistic predictor.

### Uncertainty and multiple testing

Unless otherwise stated, confidence intervals were obtained from 1,000 participant-level bootstrap resamples. Each replicate resampled participants once and recomputed all relevant outcomes, enabling paired intervals for model differences. Per-outcome multiplicity was controlled using the Benjamini–Hochberg false-discovery-rate procedure [46]. Analyses based on correlations used percentile bootstrap intervals where reported.

A disease-panel bootstrap was used only for explicitly labelled summaries of variability across outcomes, including the comparison of SHARC and conventional activity summaries.

Analyses were implemented in Python 3.12.1 using numpy 2.2.5, scipy 1.15.2, pandas 2.2.3, scikit-learn 1.5.2, PyTorch 2.5.1 and lifelines 0.30.3.

### Comparator, demographic and genetic analyses

#### Demographic comparator

The demographic comparator was a per-disease ridge Cox model containing age, sex and body-mass index, fitted in validation participants and evaluated in test participants. A jointly trained wrist-plus-demographic model concatenated the same covariates to the pooled wrist representation. A flexible demographic sensitivity model contained age, age squared, sex, body-mass index, body-mass index squared and their pairwise interactions. The out-of-fold increment from adding the frozen wrist score to this flexible model was compared with a matched permuted-score control.

#### UK Biobank activity-summary comparator

We used 100 accelerometer summary fields distributed by UK Biobank and generated with the Oxford accelerometer-processing pipeline [13]: overall acceleration mean and standard deviation, seven day-of-week averages, 24 hour-of-day averages and a 67-bin acceleration-intensity distribution. Data-quality, calibration, wear-time and device fields were excluded. Features were standardised using validation-set parameters and entered into per-disease ridge Cox models fitted in validation participants and evaluated in test participants. Their incremental value after addition to the frozen wrist score was evaluated by cross-fitting; because the 100 fields are collinear, the added block was reduced to leading principal components within each fold. The complete field list is provided in Supplementary Table 8.

#### Demographic information encoded by the wrist representation

To quantify demographic information already present in the disease-prediction representation, ridge models for age and body-mass index and logistic regression for sex were fitted on validation embeddings and evaluated once in the test set. Separate dedicated wrist estimators of age, sex and body-mass index were selected using training and validation participants and evaluated once in test participants. The dedicated age estimator additionally used four daypart summaries and a correlation loss.

#### Polygenic risk scores

For diseases with a corresponding score, we used the UK Biobank Standard PRS release derived from external genome-wide association studies [21]. Phecodes were linked to PRS traits using a curated crosswalk (Supplementary Table 5). The primary analysis was restricted to participants of European genetic ancestry (field 22006). Each disease-matched PRS was combined with the frozen wrist score using the event-stratified cross-fit Cox procedure, and the increment in C-index was estimated with participant-bootstrap intervals and false-discovery-rate correction across matched diseases. A participant-permuted PRS served as a null control.

Sensitivity analyses repeated disease-matched fusion in participants of all ancestry groups, added the complete panel of available PRS to every disease, and retrained the disease model with the full PRS panel as joint inputs. High-specificity analyses compared sensitivity at fixed specificity before and after adding the matched PRS.

### Shared and disease-specific risk analyses

#### Cross-disease correlation and comorbidity

Among the 101 outcomes with at least 50 incident test events, pairwise Spearman correlations were computed between outcome-specific predicted risk scores. Each outcome score was residualised on age, age squared, and sex, and the residuals were standardised before analysis. Incident disease co-occurrence was quantified by the phi coefficient between event indicators. We regressed demographic-residualised predicted-risk correlation on incident co-occurrence across disease pairs.

#### Shared Actigraphic Risk Component

The leading principal component of the standardised demographic-residualised outcome scores was oriented so that higher values represented greater overall predicted risk and was termed the SHARC (Shared Actigraphic Risk Component). We recorded per-disease loadings and participant-level SHARC scores and related the scores to subsequent incident-disease burden and six-year mortality. Mortality was compared between the upper and lower halves of the SHARC within four strata defined by sex and a median-age split.

#### Organ-system and disease-specific components

For each phecode category containing at least three well-powered outcomes, the standardised predicted-risk variance was partitioned into three components: variance carried by the SHARC; variance carried by the leading principal component of the within-category residual scores after projecting out the SHARC; and the remaining disease-specific variance. The fractions were normalised to sum to one within each category. To test whether individual outcome scores contained information beyond the SHARC, the outcome-specific frozen risk score was added to the SHARC using the cross-fit Cox procedure, and the out-of-fold increment in C-index was estimated with participant-bootstrap intervals. A leave-disease-out SHARC, reconstructed without the target outcome, was used as a sensitivity analysis.

#### SHARC descriptor associations

To characterise what the SHARC captures physiologically, we correlated each participant’s SHARC score with a panel of wrist-derived descriptors spanning movement volume and intensity, the non-parametric circadian rest–activity rhythm, sleep architecture and timing, and a wrist respiratory measure, together with body-mass index, prevalent and incident disease burden (the number of the 101 well-powered diseases a participant had at the time of wear and developed within six years, respectively), and six-year mortality as reference anchors. Movement descriptors were computed from the ENMO time series (mean acceleration, its 95th percentile, and the fraction of wear time in sedentary, light, moderate, vigorous and moderate-to-vigorous intensity bands using rounded operational thresholds of 30, 100 and 400 mg adapted from Hildebrand et al. [47]); circadian descriptors were the non-parametric M10, L5, relative amplitude, interdaily stability and intradaily variability with the M10 and L5 onset clock times; and sleep descriptors derived from a sleep fine-tuned model [20] (total sleep time, efficiency, fragmentation and the Sleep Regularity Index) with a wrist apnoea–hypopnoea analogue (detected respiratory events per hour of sleep) were derived from the sleep-stage and respiratory outputs. The Sleep Regularity Index is the percentage probability of being in the same state, asleep or awake, at two time points 24 hours apart, rescaled so that 100 indicates identical sleep and wake times each day and 0 a random pattern [48]. UK Biobank’s distributed overall acceleration was included as an external anchor. Each descriptor was computed per recording and averaged to one value per participant over the held-out test cohort (*n* between 5,209 and 5,253, depending on descriptor missingness). For each descriptor we report the Spearman correlation with the SHARC and a 95% confidence interval from 2,000 participant bootstrap resamples. To test whether these associations are independent of adiposity, we additionally computed, for the wrist descriptors, a body-mass-index-adjusted partial Spearman correlation, residualising the ranks of the SHARC and of the descriptor on the rank of body-mass index before correlating (Extended Data Figure 10).

#### SHARC as a single-axis predictor

The SHARC was evaluated as a fixed per-disease predictor using the same six-year AUROC protocol as the 100-field activity-summary comparator. It received no per-disease refitting. The mean AUROC difference across outcomes was summarised using 2,000 bootstrap resamples over diseases, and metric harmonisation was checked by recomputing wrist-model AUROC from the same labels.

### Source-attribution analyses

Encoder and modality attributions used on-manifold conditional permutation ablation [49] on the frozen model with no retraining, using ten permutation seeds per cell across four independently trained model seeds; the day/night attributions used the same ablation with five permutation seeds per cell across the four model seeds and additionally remove-and-retrain [50], retrained once per cell and model seed. Daytime and night-time are fixed local-clock windows (09:00–21:00 and 21:00–09:00), applied identically to every participant rather than individualised to each person’s sleep period. Both encoders run over the full recording, day and night alike; the daytime and night-time sources are formed by pooling each encoder’s per-patch embeddings within these two windows, so the labels denote when a signal occurs, not a restriction on what the model reads. As faithfulness checks, replacing the trained weights with a random initialisation reduced concordance to chance and collapsed the attributions, and permuting the outcome labels reduced concordance to 0.50; every reported value was reproduced by a second, from-scratch reimplementation, sharing only the survival-analysis library, to within 0.001. A source’s contribution was denoted as significant when, for the wrist components, its conditional-permutation attribution passed Benjamini–Hochberg false-discovery-rate control across the disease panel; for the polygenic score, its 95% confidence interval excluded zero; and for age, sex and body-mass index, it exceeded the 95th percentile of a matched covariate-permutation null.

### Prodromal neurodegeneration analyses

Because incident neurodegenerative events on the held-out test set are few, we quantified the prodromal signal with a cross-validated readout of the frozen embedding across the whole cohort. An L2-regularised logistic readout was trained on the concatenated frozen wrist embedding by five-fold cross-validation with subject-level folds, and each participant was scored out of fold, so no participant’s diagnosis entered its own score; the encoders are self-supervised on UK Biobank, so the readout, not the representation, is the held-out element. We report the population time-dependent AUROC and average precision on the full at-risk cohort at the natural incident base rate, with inverse-probability-of-censoring weighting [51, 52], and a lead-time washout that excludes cases diagnosed within one to three years of the recording. Incident timing came from first-diagnosis dates, and prevalent cases were excluded.

### Calibration and model robustness

#### Calibration and proportional hazards

Absolute six-year risks for calibration plots were reconstructed using an event-stratified within-test cross-fit Breslow baseline [53], with the frozen linear predictor entered as an offset and the baseline cumulative hazard fitted out of fold. We report calibration slopes and curve shape in the held-out UK Biobank test set. Calibration slopes were estimated only for outcomes with at least five incident test events. Because the primary model’s original baseline hazard was not retained, these analyses do not estimate calibration-in-the-large of an externally transported model. Proportional hazards were assessed per outcome using scaled Schoenfeld-residual tests, with false-discovery-rate correction.

#### Internal geographical replication

UK Biobank assessment centres were grouped into geographical regions that were not used as model inputs. We evaluated the primary deployed model among held-out test participants in each region. Separately, a regularised Cox head was fitted on the frozen pooled representation while leaving one region out of downstream training and evaluated in that region. This leave-region-out analysis was restricted to the 60 best-powered outcomes and represents internal geographical replication rather than external validation.

#### Diagnostic washout

To assess sensitivity to near-term diagnosis, each incident outcome was re-evaluated after excluding cases diagnosed within two years of accelerometer wear. Frozen test-set scores were retained and only case inclusion changed. We summarised the mean and median change in C-index across the 388 evaluable outcomes and tabulated outcomes whose C-index decreased by more than 0.05.

#### Clinical utility

We quantified pre-diagnostic utility for Parkinson’s disease using the cross-validated cohort readout recalibrated to absolute six-year risk. We computed a time-to-event decision curve [54] (net benefit against threshold probability, with observed incidence in the flagged subgroup) comparing the wrist readout with screening everyone, screening no one, and an age, sex and body-mass-index baseline, and screening yield (sensitivity, specificity, positive predictive value, number-needed-to-screen and fold-enrichment of the incident rate) at the top-10% and 95%-specificity operating points, on the unenriched cohort at the natural base rate, with subject-level bootstrap intervals.

### Prevalent-disease screening

We tested whether the frozen embedding detects disease already present at the time of the recording. An outcome was counted as prevalent when its diagnosis was recorded at or before the wrist recording, within a seven-day window; controls had no record of the outcome. Because prevalent cases of the diseases of interest are rare (132 for Parkinson’s disease, 16 for Alzheimer’s disease and 21 for dementia across the whole cohort), a held-out test set would contain too few of them for a stable per-disease estimate. As for the prodromal readout, we therefore scored the whole cohort out of fold rather than the held-out test set: every case is scored by a model fitted only on other participants, and because the encoders are self-supervised, the readout rather than the representation is the held-out element. For each outcome, we ran a same-distribution five-fold cross-validation, with folds assigned by a deterministic hash of the participant identifier, and pooled the out-of-fold predictions over all prevalent cases and a fixed pooled control set. Because a prevalent diagnosis is a cross-sectional present-or-absent label with no onset time, the readout was a classifier rather than the survival model used for incident prediction. The classifier was an L2 logistic regression on the frozen 1,536-dimensional L-moment wrist embedding, averaged across all valid days of each recording (scikit-learn, lbfgs solver, up to 2,000 iterations, class-weighted), with no demographics, per-fold standardisation, and the regularisation strength (*C ∈ {*0.01, 0.1, 1*}*) selected on a validation carve-out. We report the AUROC with a 1,000-resample bootstrap confidence interval. The demographic baseline was a logistic model on age, sex and body-mass index over the same folds. We screened the 632 outcomes with enough prevalent cases, and counted an outcome as a robust win when its wrist confidence interval cleared the baseline.

### False-positive analyses

To characterise how the model over-ranks patients for diseases they do not have, we residualised each disease’s predicted six-year risk on age, age^2^ and sex and converted it to a cohort percentile. For each disease, we averaged, over the participants who truly had it, the percentile the model assigned for every other disease, giving a directed confusion value. Diseases with at least ten incident test cases (314 of 390) contributed a true-disease profile, while the diseases they could be over-predicted on spanned all 390 scored outcomes.

For each of ten prespecified diseases, we defined the false positives as test participants whose six-year risk exceeds the disease’s 95% specificity threshold but who never had that disease recorded, either before or after accelerometer wear. At this operating point, we reported positive predictive value and the implied number of false positives per 100 flagged participants. For every other outcome, we computed its enrichment among the false positives, the rate of that outcome (ever diagnosed, prevalent, or incident) in the false-positive group divided by its rate in the whole test cohort. Candidate outcomes were restricted to those with at least 30 incident cases and present in at least 10 false-positive participants. Each enrichment carried a 95% confidence interval from 2,000 bootstrap resamples of the false-positive group, and the six outcomes shown per disease were those with the highest lower confidence bound, a conservative ranking that keeps a rare outcome with a wide interval from dominating.

To test whether these misdiagnoses reflect disease-specific structure rather than the false positives simply being frailer, we recomputed each enrichment with adjustment for comorbidity burden (Supplementary Table 9). As a descriptive effect size we standardised directly on burden: we counted each participant’s prevalent diagnoses across the panel, split the never-target participants into deciles of this count, and computed a burden-matched enrichment *E^∗^* as the outcome rate among the false positives divided by the rate expected from the true negatives at the false positives’ burden distribution. Because a prevalent outcome is itself part of the burden count, we computed the enrichment for ever-diagnosed, prevalent-and incident-only outcomes but report only the incident-only comparison, in which the outcome is a future event and which is least mechanically coupled to the prevalent-comorbidity measure; the ever-diagnosed and prevalent counts are small and are not shown. For formal inference, we fitted, per outcome, a logistic regression of the outcome on false-positive status, adjusting for age, sex, and comorbidity burden, and report the adjusted odds ratio, the estimator that directly removes age, which the matching does not. For Parkinson’s and Alzheimer’s disease, we pre-specified, before analysis, a list of prodromal and associated conditions from the literature and report every one with its adjusted odds ratio and a Benjamini–Hochberg false-discovery rate within each disease family, so that a null cannot be hidden. As a robustness check, we repeated the matching on deciles of the SHARC; because the SHARC is computed on demographic-residualised risks, it is nearly orthogonal to age and only weakly correlated with realised comorbidity, so it probes robustness to the SHARC rather than to raw comorbidity burden. The false-positive bootstrap interval on *E^∗^* holds the control rates fixed and is therefore approximate, so significance is taken from the adjusted odds ratio.

### Representation and aggregation comparisons

We performed a limited set of descriptive comparisons to quantify the information added by successive representation and aggregation choices. With the downstream survival model, age and sex covariates, participant split, and evaluation procedure held fixed, we compared hand-engineered wrist summaries, mean-pooled frozen embeddings, and distributionally pooled frozen embeddings based on the first three L-moments. We additionally evaluated one alternative sequence model that retained the temporal ordering of the patch embeddings. These analyses were evaluated on the test cohort and are therefore interpreted as descriptive comparisons of the implemented pipeline, rather than as an independent validation of architecture selection.

### The age-pretrained sequence model

The sequence model applies a two-layer rotary transformer to each encoder’s ordered per-patch embeddings, with four attention heads and a six-head masked attention pool; the two encoder streams are combined by averaging their outputs. It was pretrained to estimate chronological age under an inverse-frequency-weighted L1 loss, with each recording tiled into six-hour blocks and 75% of the blocks masked at random. Because the masked patches are dropped before the transformer, an additive real-elapsed-time positional encoding re-injects the true elapsed time between the retained patches. The pretrained body was then fine-tuned to the disease panel. Both stages used AdamW. Pretraining ran for up to 25 epochs at learning rate 10*^−^*^4^ (batch size 16), early-stopped on the validation age error. Fine-tuning used a 4-epoch linear-probe phase at learning rate 10*^−^*^3^ followed by up to 8 epochs of full fine-tuning at 10*^−^*^4^ (batch size 12), early-stopped on the validation loss. It uses a 20-bin discrete-time logistic hazard rather than the Cox partial likelihood, because the in-batch Cox risk set starves at the small batch sizes a per-patch sequence requires; we therefore report both the Cox mean-pool bar and a covariate-matched mean-pool baseline, and compare the sequence model to a mean pool of the same per-patch embeddings on the same subjects and diseases (an eid-and-phecode intersection; Extended Data Figure 4d). It ran on the complete per-patch embedding cache, on the full 390-outcome panel of the canonical split, matching the main model; the comparison is internally matched.

#### Feature-source ladder

To quantify how much the learned representation contributes, we held the survival model, the split, the covariates, and the evaluation fixed, and replaced only the daily feature block (Extended Data Figure 4a). The learned arm used the per-day mean of the frozen embedding. The summary-statistic arm used a standard panel of twelve hand-engineered wrist measures (mean acceleration and its dispersion, time in moderate-to-vigorous activity, the nonparametric rest–activity measures M10, L5, relative amplitude, intradaily variability and interdaily stability, cosinor amplitude, acrophase and mesor, and a predicted-wake fraction), z-scored on the training set. All but the predicted-wake fraction are computed from the raw accelerometer signal; the predicted-wake fraction is the mean wake probability from the AcceleRest sleep-stage predictions. The untrained-encoder arm re-initialised both encoder architectures with random weights, matched to each architecture’s own initialisation, and ran the identical extraction and pooling on the real signal. The matched-dimensionality random arm replaced the embedding with independent Gaussian noise of the same shape. Every arm trained the same rolling-window Cox head with the same recipe and was evaluated identically; the random and demographic floor coincide because all arms retain the age and sex covariates.

#### Context titration

With the trained model fixed, we truncated its input at inference, with no retraining. For days of wear, we restricted each recording to its first *k* calendar days and ran the model’s standard inference, averaging all valid three-day windows within those days; for *k <* 3, below the model’s window, we fed a single masked partial window and flag these points as an out-of-distribution extrapolation. For hours per day, we re-pooled each day’s per-window embeddings into the daily distributional summary using only the first *H* hours of the noon-to-noon day, then ran the same fixed model. The full-context points reproduce the headline concordance exactly.

## Data Availability

This study was conducted under UK Biobank Application Number 62249. UK Biobank data are available to approved researchers through the UK Biobank Access Management System (https://www.ukbiobank.ac.uk). In line with UK Biobank policy, individual-level data derived from UK Biobank, including trained model weights and participant-level embeddings, are treated as equivalent to individual-level data and cannot be shared, egressed or published, and are available only to approved researchers within the UK Biobank environment. The aggregate data underlying the figures are provided with the manuscript. Code to reproduce the analyses and figures will be made available in a public repository upon publication.

## Data availability

The UK Biobank data are available to approved researchers through the UK Biobank Access Management System (https://www.ukbiobank.ac.uk); this study used the resource under Application Number 62249.

## Code availability

The accelerometer foundation-model encoders are previously published [19, 20]. Upon acceptance, the code to reproduce the analyses and figures will be made accessible in a public repository. In line with UK Biobank policy, trained model weights and participant-level embeddings derived from UK Biobank data are treated as equivalent to individual-level data and cannot be shared, egressed, published, or commercialised; they are available only to approved researchers within the UK Biobank environment.

## Extended Data

This section preserves every approach taken in the project, including those that did not improve on the frozen representation, together with the supporting analyses and the full per-disease table.

## Author contributions

M.D. and N.R.L. contributed equally to this work. M.D. conceived and designed the study, developed the code, performed all analyses, curated the data, produced the figures and wrote the original draft. N.R.L. developed and provided the pretrained accelerometer foundation models and the cohort data preparation, and contributed methodology and expertise. M.R.K. contributed methodology and analysis guidance. A.B. contributed sleep-medicine and genetics expertise and to the interpretation of the results. P.J. provided clinical guidance and supervision, and E.H.D. contributed clinical and neurological expertise. J.Z. contributed methodology and machine-learning expertise. E.M. and A.B.-K. jointly supervised the study: E.M. conceived the study and provided resources, A.B.-K. contributed methodology, and both guided the analysis and interpretation. All authors reviewed and edited the manuscript.

## Acknowledgements

This research has been conducted using the UK Biobank Resource under Application Number 62249. This work uses data provided by patients and collected by the NHS as part of their care and support. We thank the UK Biobank participants and the UK Biobank team. Analyses used the computing resources of the laboratory of E.M. at Stanford University. This research was funded by Takeda and by unrestricted funds and gifts to E.M.

## Supplementary Information

**Supplementary Figure 1.**
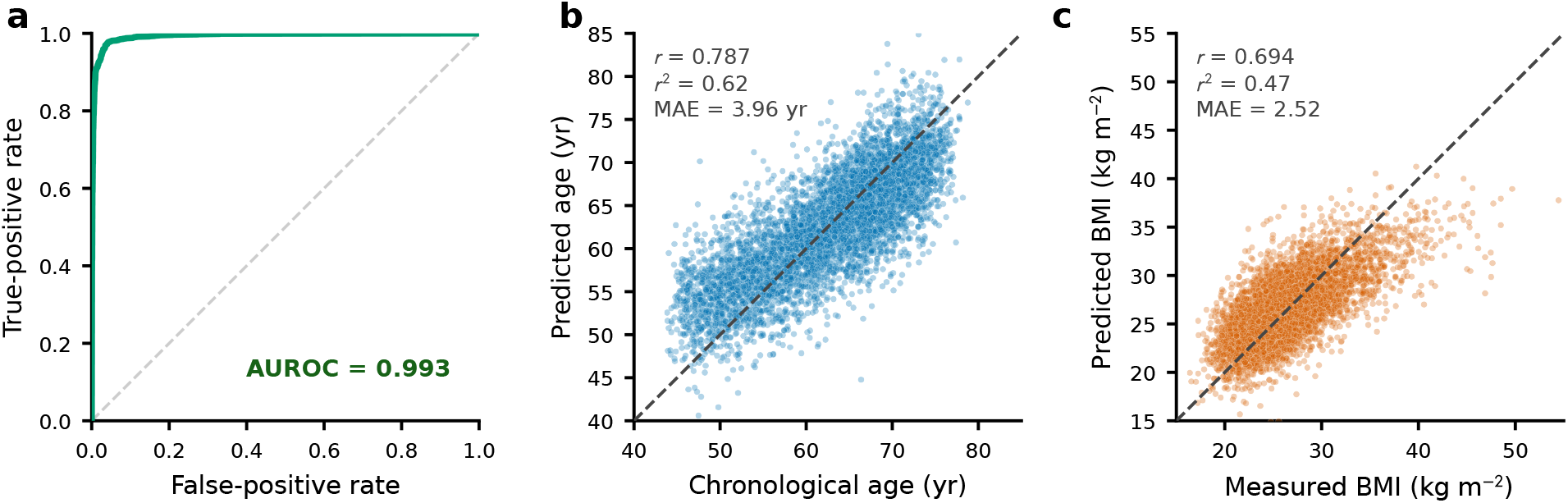
Why demographics add little: the disease-prediction embedding already encodes age, sex and body-mass index. Sex (**a**), age (**b**) and body-mass index (**c**) read directly from the frozen wrist embedding used for disease prediction, by a linear model fit on the validation split and scored on the held-out test set (Methods). **a**, Sex from a receiver-operating-characteristic curve. **b**, Chronological age and **c**, body-mass index as predicted versus measured values, with the Pearson correlation, its square and the mean absolute error; the dashed line marks equality. The embedding linearly recovers age (*r*^2^ = 0.62), sex (AUROC 0.99) and body-mass index (*r*^2^ = 0.47). Because the embedding already carries these variables, adding age and sex back to the disease model raises the mean concordance by only +0.0045 (95% CI +0.0005 to +0.0085) and the mean six-year AUROC by +0.0006; dedicated wrist estimators recover the same three variables more accurately still (Figure 3d–f).

**Supplementary Figure 2.**
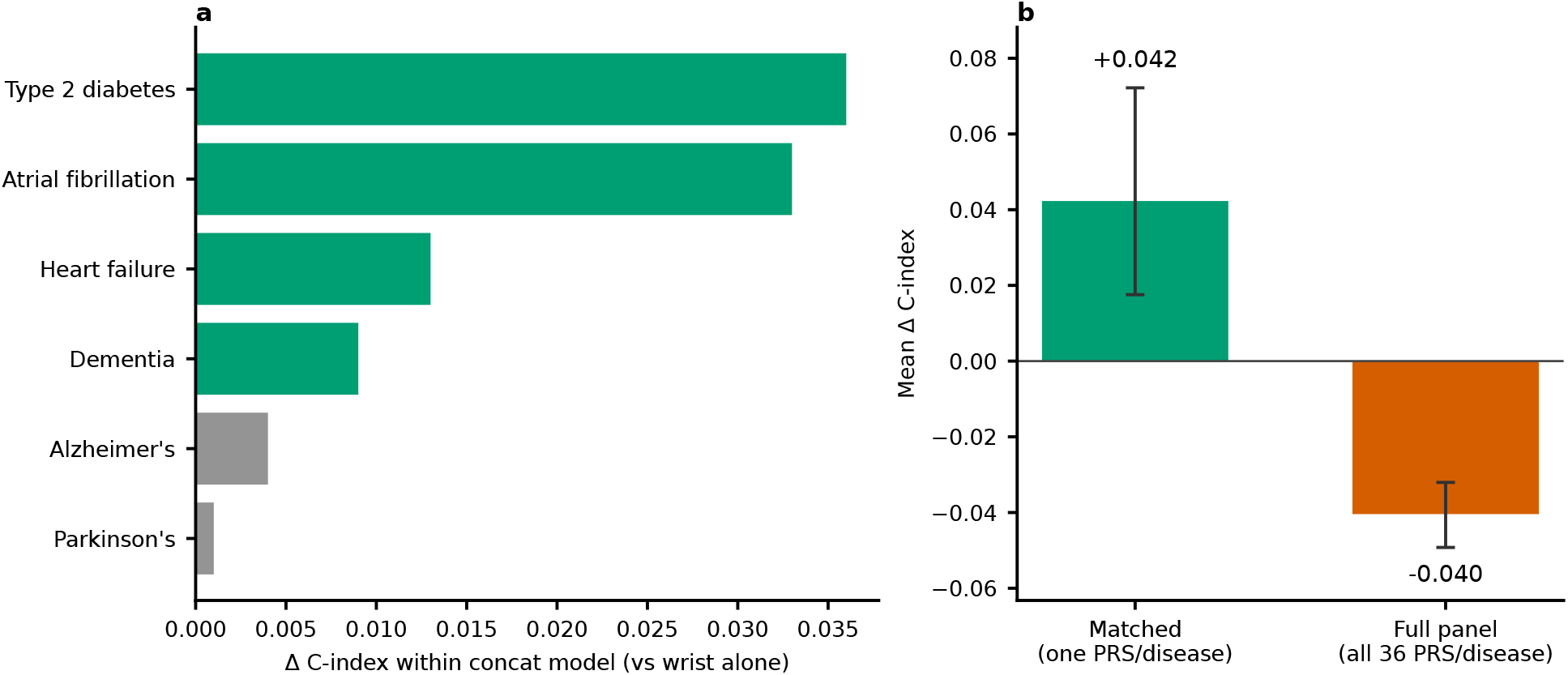
Genetics must be matched to the disease. (a) Retraining the embedding model with the full panel of 36 polygenic scores as joint inputs leaves the overall mean concordance unchanged (0.6875 versus 0.6880 for the wrist alone), yet the cardiometabolic outcomes still improve inside that single model. A model with one fixed score vector cannot route each score to the disease it predicts. (b) The disease-matched analysis, with one score per disease, gains +0.042, whereas feeding all 36 scores to every outcome as features loses 0.040 to noise. Held-out test set. Error bars in panel b are 95% confidence intervals from a phecode-panel bootstrap over 31 matched and 332 full-panel diseases; panel a shows point estimates.

**Supplementary Figure 3.**
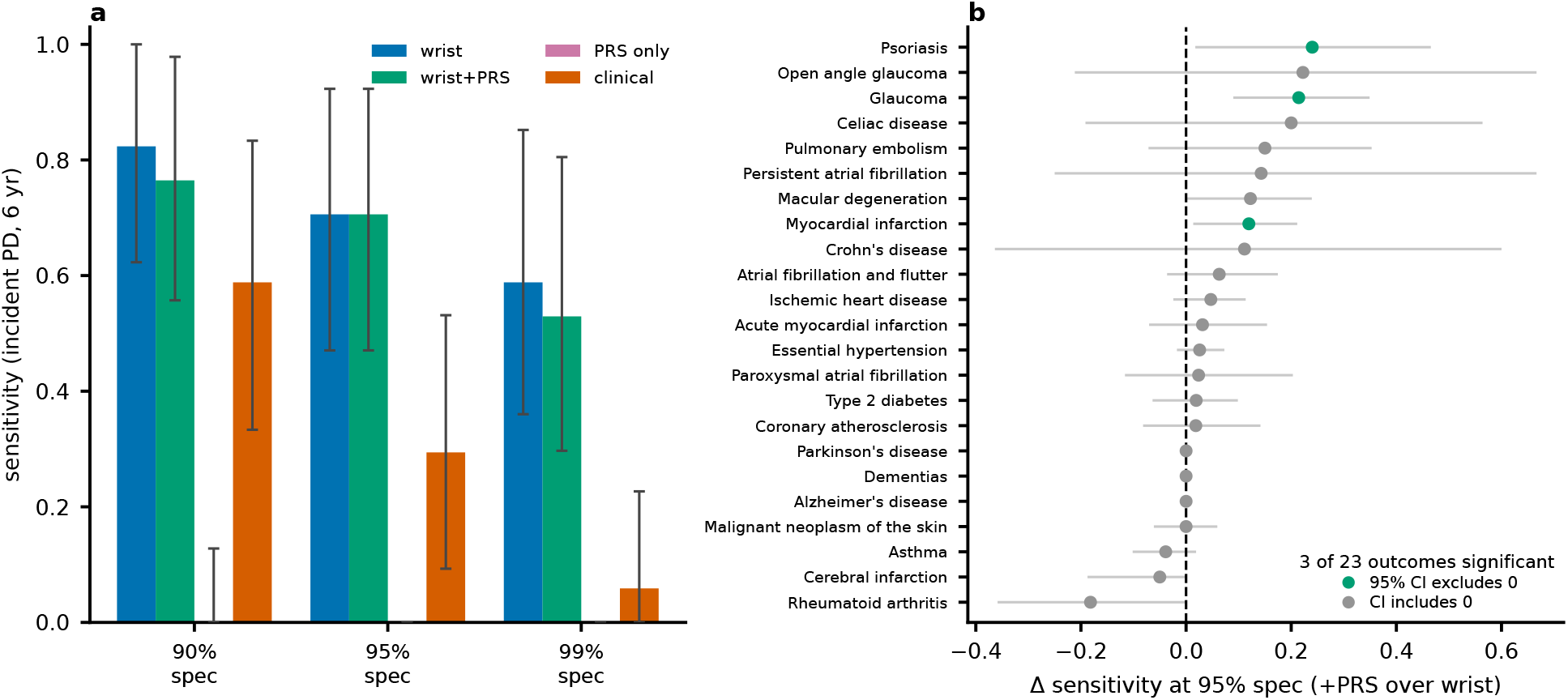
A polygenic risk score does not sharpen high-specificity screening, including for Parkinson’s disease. Incident six-year screening on the European-ancestry test set. **a**, Sensitivity for Parkinson’s disease at fixed specificities for the wrist model, the wrist model combined with the Parkinson’s polygenic score through a leakage-safe out-of-fold Cox fusion, the polygenic score alone, and an age, sex and body-mass-index baseline. The wrist model reaches an area under the curve of 0.96; the polygenic score alone is at chance (0.48) and flags no cases at any operating point; adding it to the wrist model does not raise sensitivity. Error bars in panel a are 95% subject-bootstrap confidence intervals, which are wide given the small number of incident cases. **b**, Change in sensitivity at 95% specificity from adding the matched polygenic score to the wrist model, per matched disease, with paired subject-bootstrap 95% intervals; green marks intervals that exclude zero. The score helps a small number of cardiometabolic and autoimmune outcomes at moderate specificity (3 of 23) and none at 99% specificity, and it does not help Parkinson’s or Alzheimer’s disease. A cross-sectional prevalent-detection companion (accompanying table) reaches the same conclusion. This area under the curve is a six-year fixed-horizon incident AUROC on the European-ancestry test set, based on 17 incident Parkinson’s cases, and it is a different estimator from the full-cohort time-dependent AUROC of 0.90 reported in the main text.

**Supplementary Figure 4.**
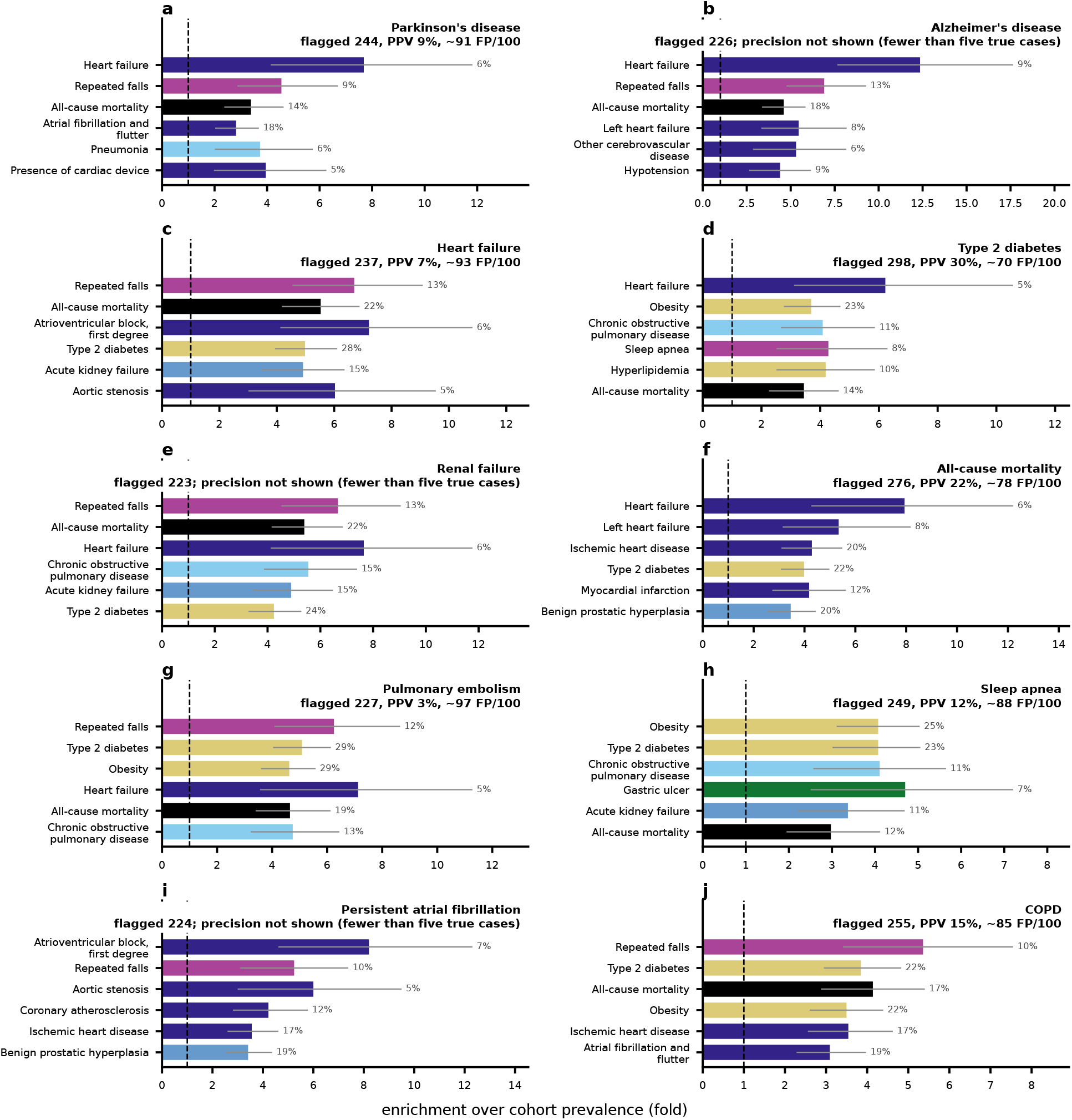
What the model’s false positives actually have. For each of ten diseases (**a**–**j**, European-ancestry test set), the false positives are participants flagged at the disease’s 95% specificity threshold who never have it, prevalent or incident (Methods). Each panel title gives the precision at this operating point (positive predictive value and false positives per 100 flagged), which is low because these outcomes are rare; for diseases with fewer than five true cases at this threshold the precision is not shown, in line with UK Biobank’s small-numbers policy. Bars show the conditions the false positives actually carry, on an axis of enrichment: the false-positive rate of a condition divided by its cohort rate (a value of 4 is fourfold; the dashed line marks the base rate). The number after each bar is the share of false positives with the condition, each bar has a 95% bootstrap interval, and the six shown are those with the highest lower bound. Colour is the condition’s clinical category. The misdiagnoses are clinically coherent: Parkinson’s-disease false positives carry a cardiac and frailty pattern, sleep-apnoea false positives the metabolic-syndrome cluster, and type-2-diabetes false positives cardiometabolic disease. A companion analysis (Supplementary Table 9) confirms this is not merely because flagged participants are frailer: matching them to controls of the same comorbidity burden barely changes the enrichments (rank correlation 0.95).

**Supplementary Figure 5.**
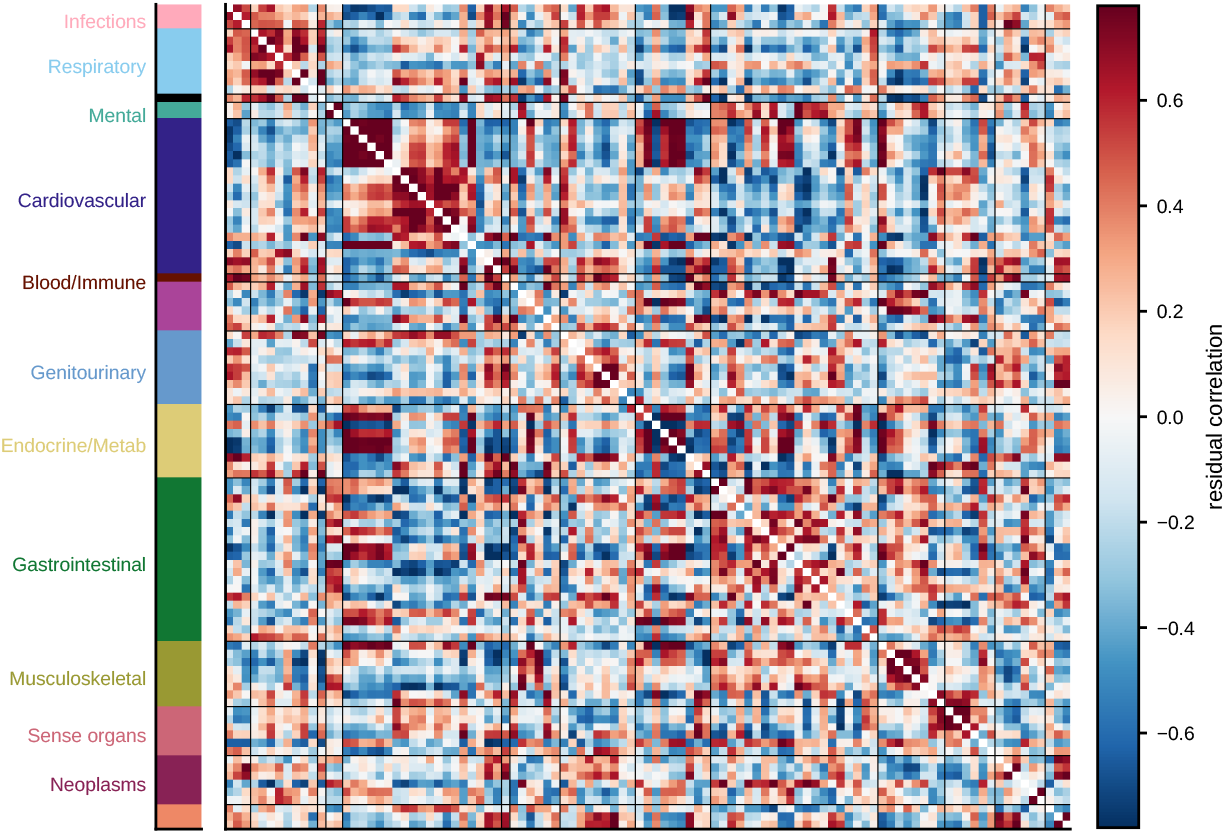
Residual disease-risk correlation after the SHARC is removed. Pairwise Pearson correlation of the predicted six-year risks, for the 101 outcomes with at least 50 incident cases, after the SHARC is projected out, ordered by organ system (colour bar at left; Methods). The dense predicted-risk correlation (Extended Data Figure 8d) collapses into weak, block-diagonal structure concentrated within organ systems; the SHARC therefore carries most of the cross-disease correlation. This is the qualitative companion to the per-disease residual concordance in Figure 2c.

**Supplementary Table 1.** The Shared Actigraphic Risk Component separates six-year mortality within sex and age strata. Observed six-year all-cause mortality among the 5,253 held-out test participants, within four strata defined by sex and a median-age split and, within each stratum, by the upper versus lower half of the SHARC score. For each half, *n* is the number of participants and Deaths the number of six-year deaths. Mortality <u>is higher in the upper half in every stratum; event counts are small in t</u>he younger strata.

| Stratum | Upper-half SHARC |  |  | Lower-half SHARC |  |  |
| --- | --- | --- | --- | --- | --- | --- |
| | $n$ | Deaths | Mortality | $n$ | Deaths | Mortality |
| Older women | 689 | 29 | 4.2% | 688 | 7 | 1.0% |
| Older men | 626 | 43 | 6.9% | 625 | 23 | 3.7% |
| Younger women | 786 | 7 | 0.9% | 785 | 5 | 0.6% |
| Younger men | 527 | 11 | 2.1% | 527 | 6 | 1.1% |

**Supplementary Table 2.**
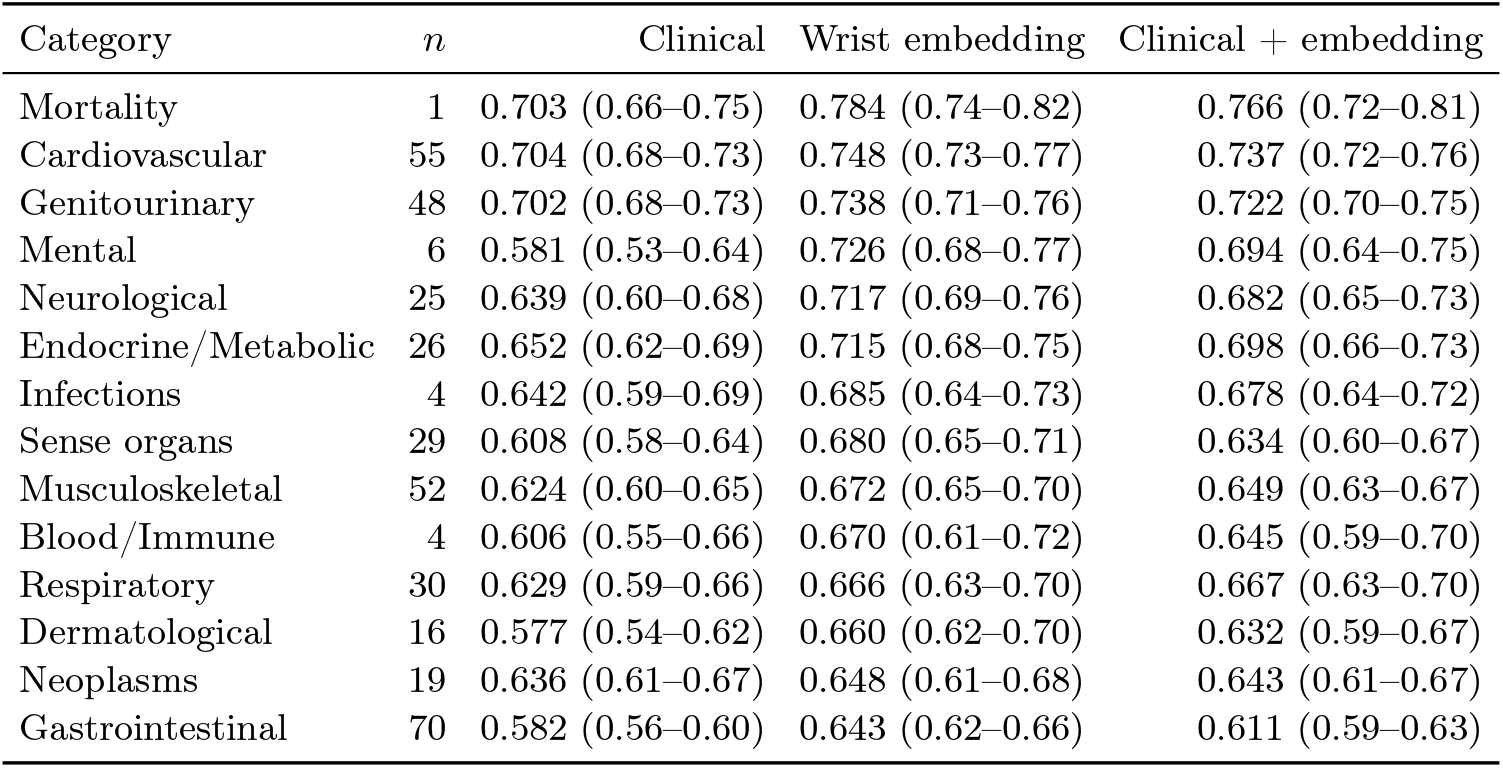
Per-category six-year AUROC: age, sex and body-mass-index baseline versus wrist embedding. Mean AUROC within each disease category for the age, sex and body-mass-index baseline, the wrist embedding, and a per-disease combiner of the clinical covariates with the wrist embedding, each with a 95% confidence interval from a subject-level cluster bootstrap (1,000 resamples of the shared test participants, recomputing every disease and averaging within the category). The combiner is biased downward by per-outcome estimation cost; the jointly trained clinical-plus-embedding model reaches a mean AUROC of 0<u>.692.</u> *<u>n</u>* <u>is the number of outcomes with a valid six-year AUROC in the category.</u>

**Supplementary Table 3.**
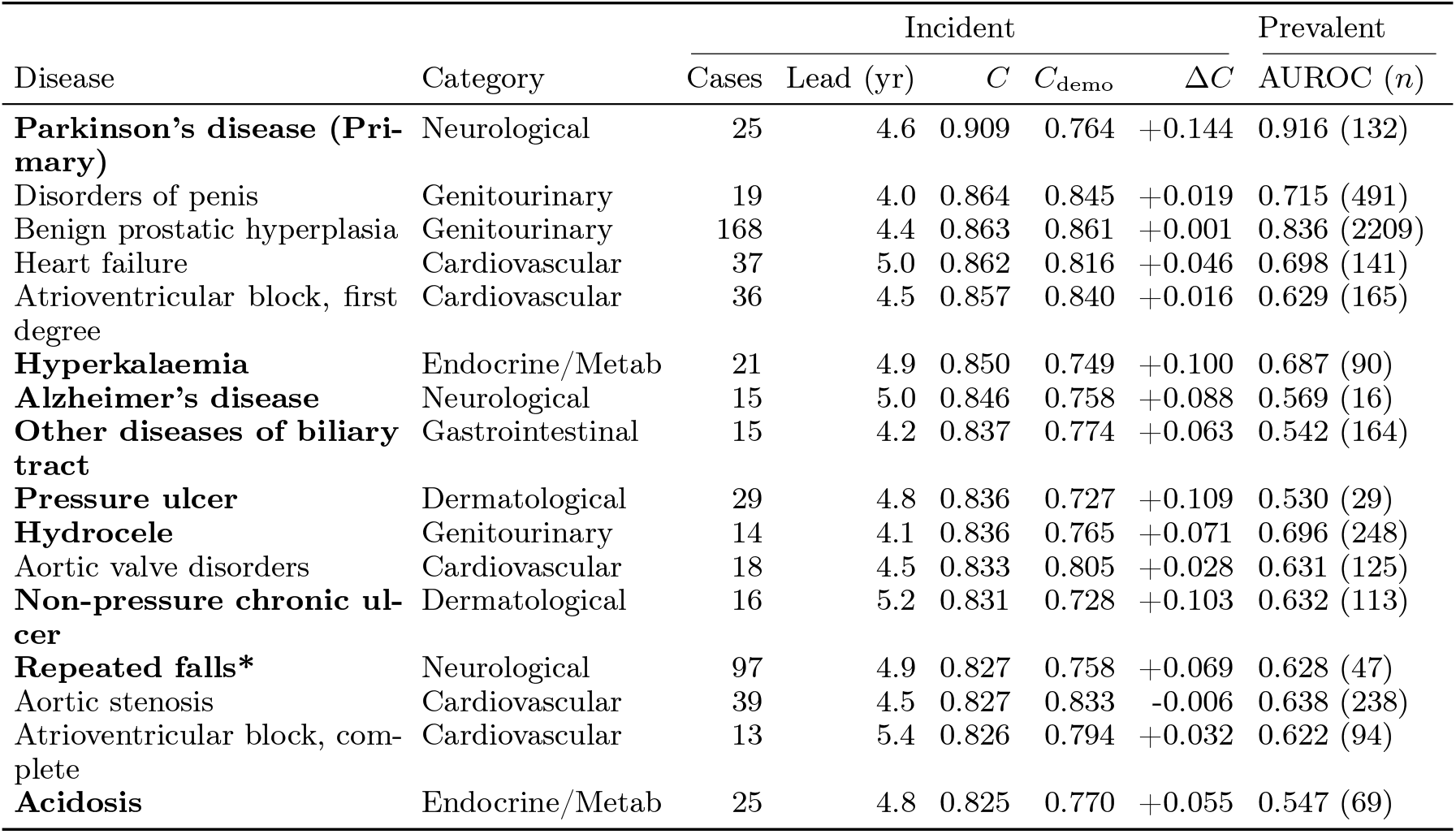
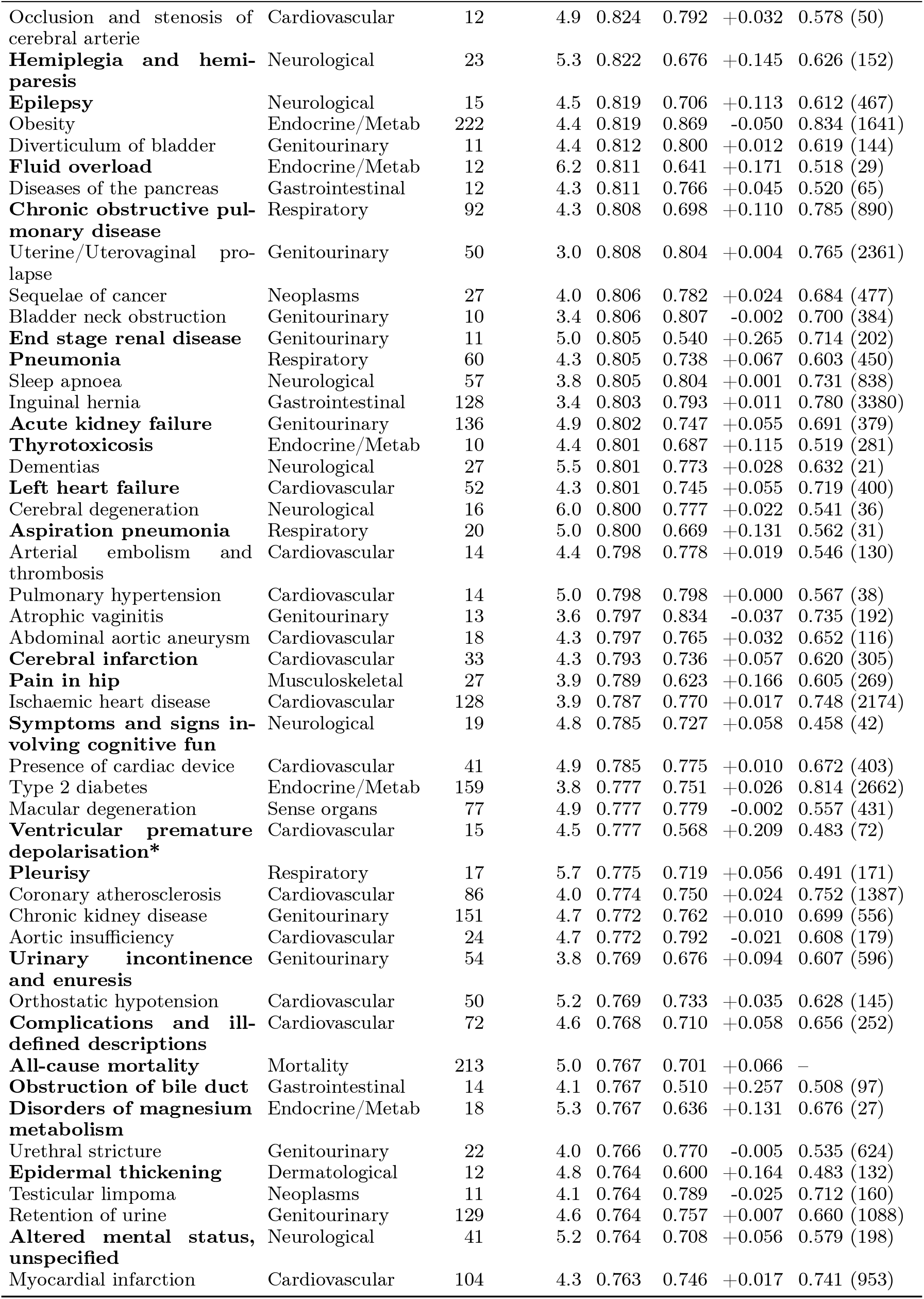
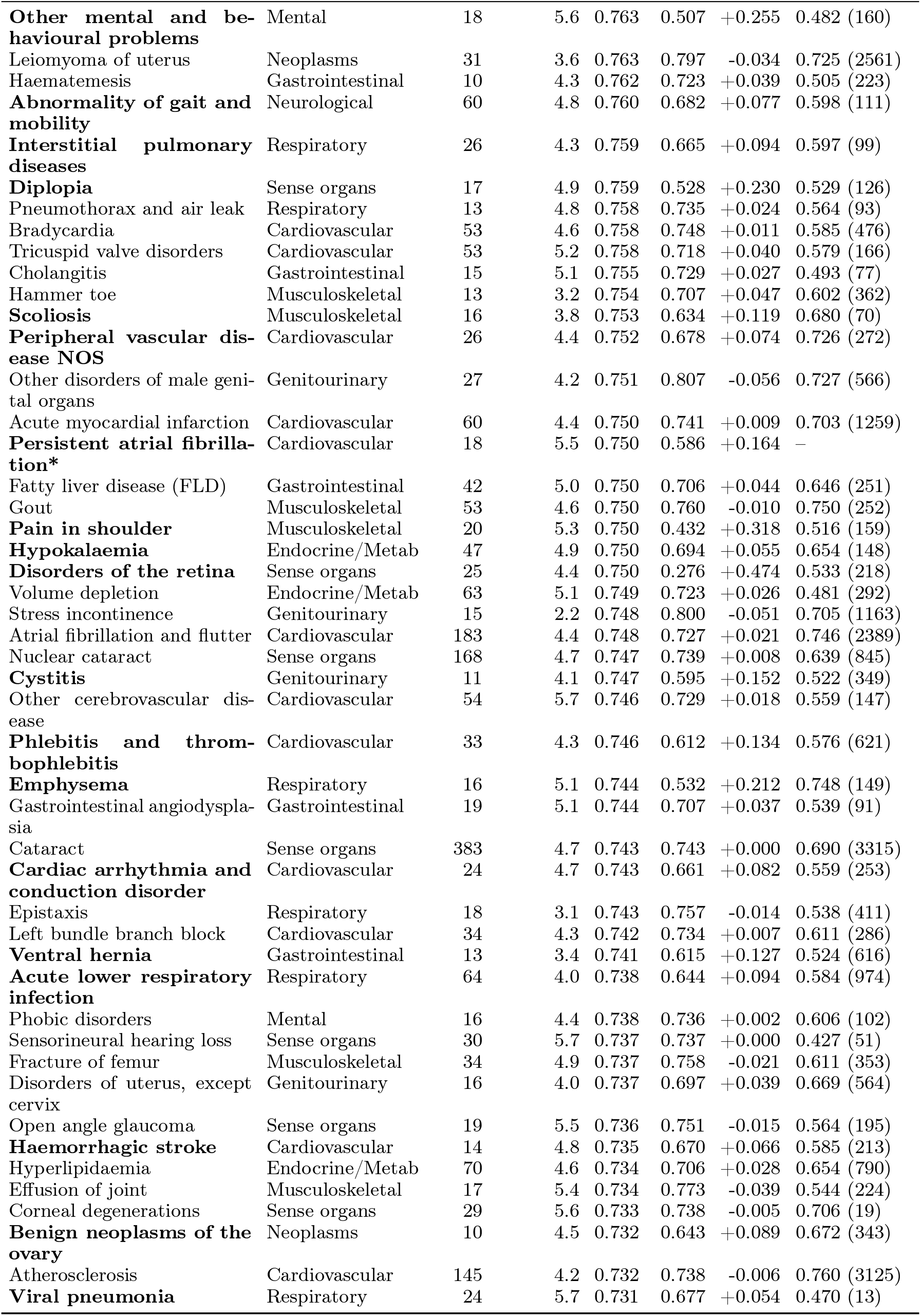
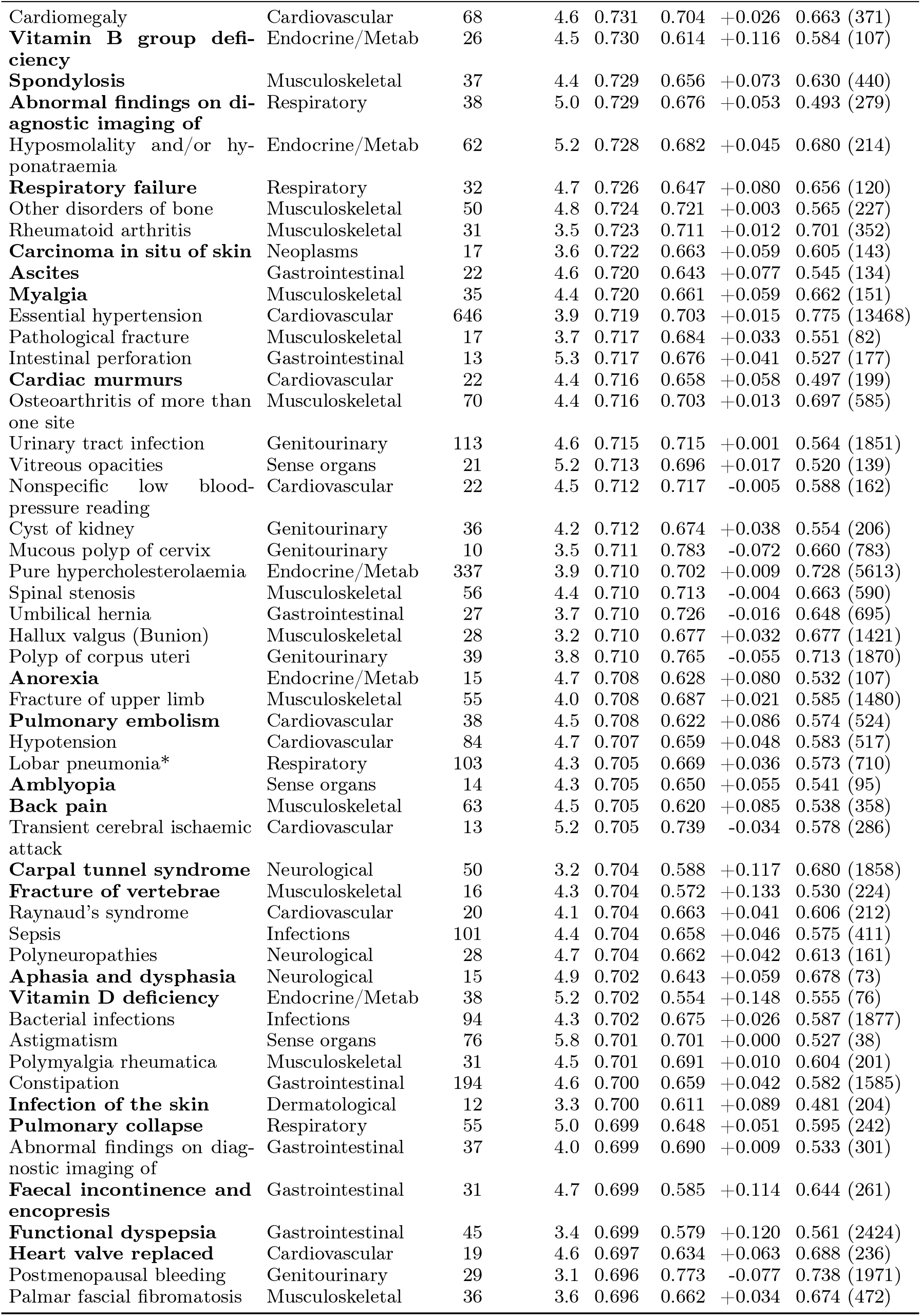
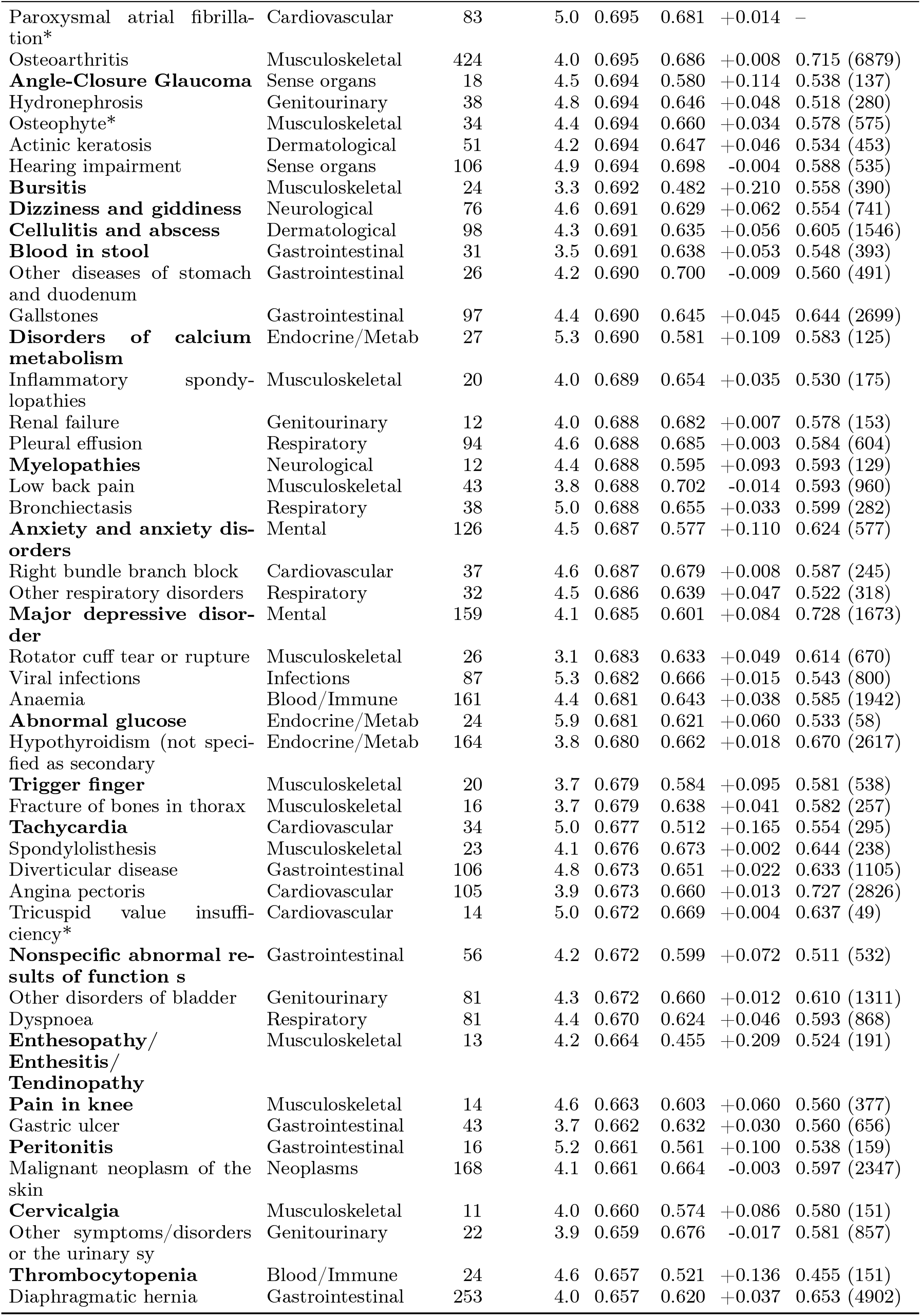
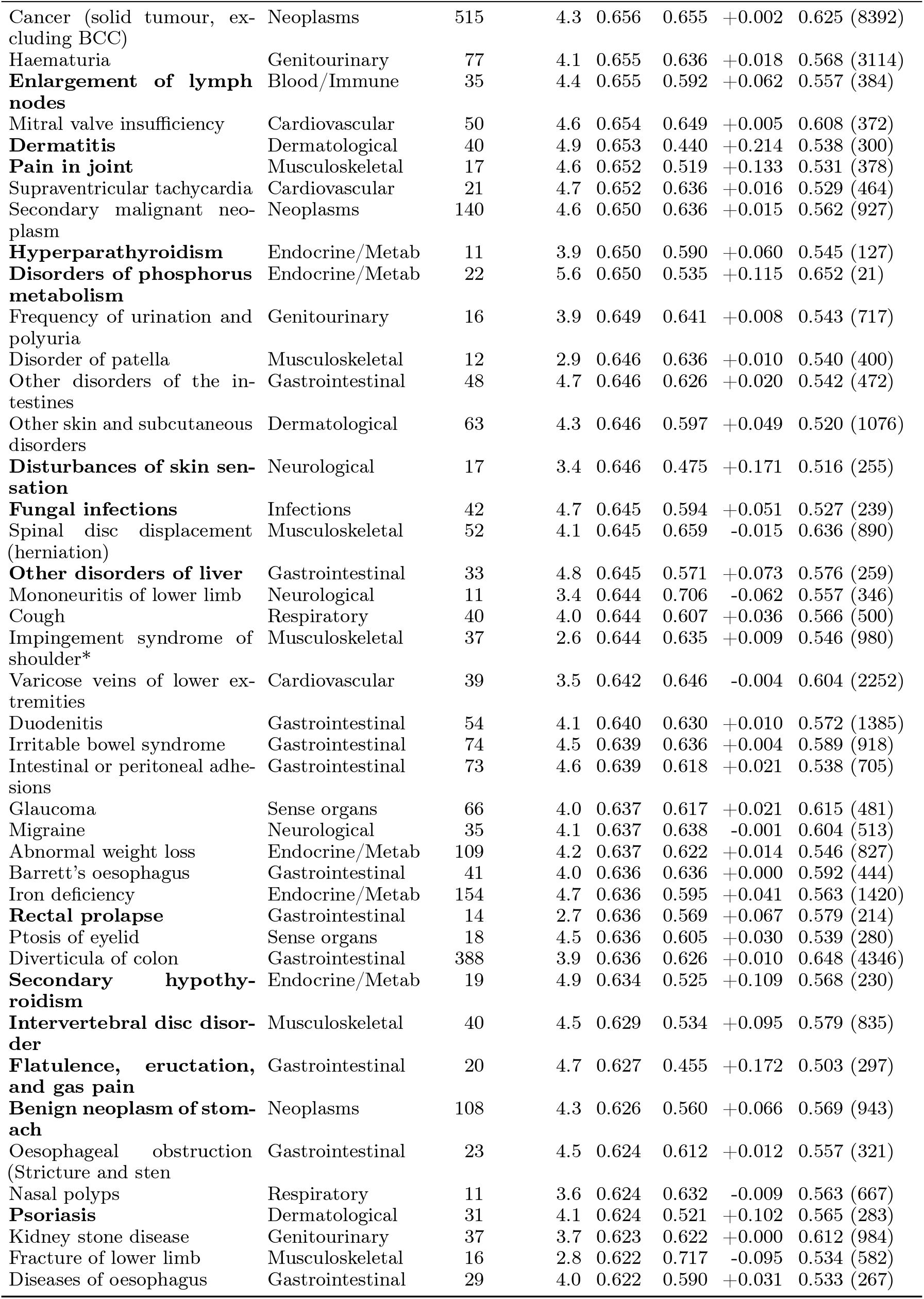
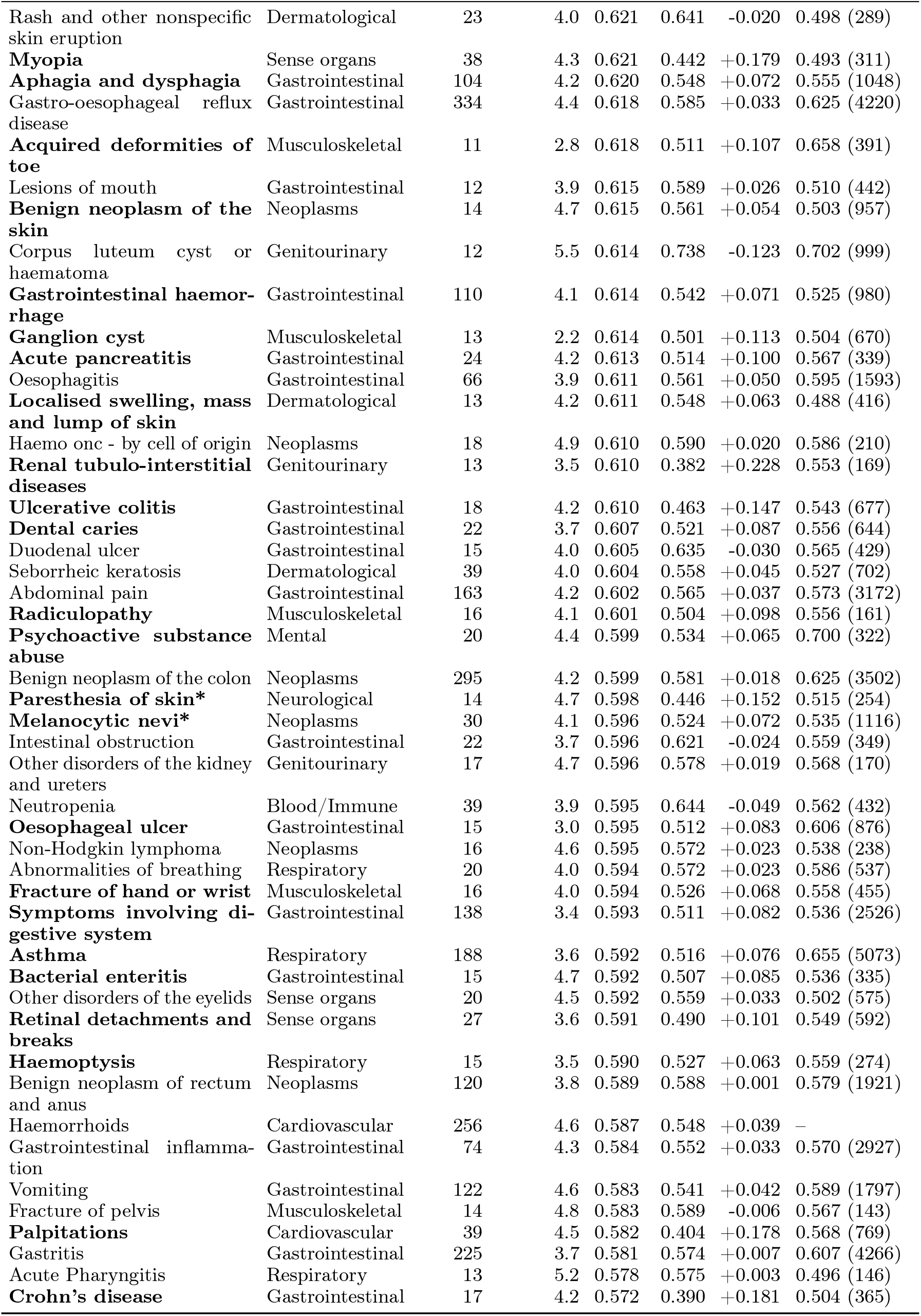
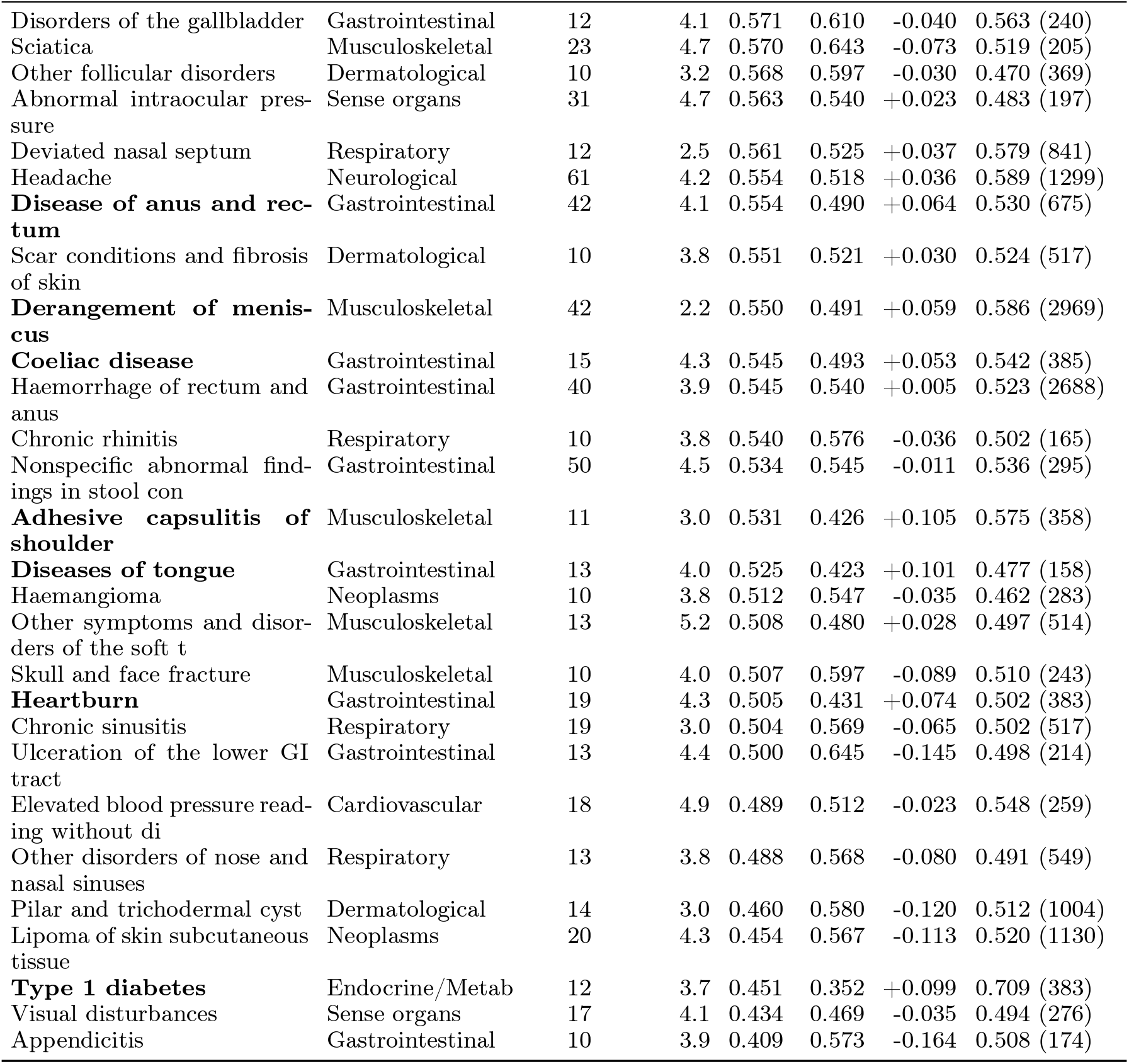
Per-disease incident prediction, prevalent-disease screening and a demographic baseline. Outcomes with at least 10 incident test cases, ordered by incident concordance. The header groups the columns into an *Incident* block (incident prediction) and a *Prevalent* block (prevalent-disease screening). *Cases* is the number of incident test cases; *Lead* is the mean time from the wrist recording to diagnosis among those cases (years). *C* is the incident concordance of the wrist embedding; *C*_demo_ is an age, sex and body-mass-index Cox baseline for the same outcome; Δ*C* is their difference. The *Prevalent* column is the wrist embedding’s prevalent-disease screening AUROC (same-distribution cross-validation), with the number of prevalent test cases in parentheses; a dash marks outcomes with no viable screening cell, and all-cause mortality has no prevalent state. Diseases in **bold** improve on the demographic baseline by more than 0.05 in concordance (Δ*C >* 0.05).

| Disease | Category | Incident |  |  |  | Prevalent |  |
| --- | --- | --- | --- | --- | --- | --- | --- |
| | | Cases | Lead (yr) | $C$ | $C_{\text{demo}}$ | $\Delta C$ | AUROC ($n$) |
| **Parkinson’s disease (Primary)** | Neurological | 25 | 4.6 | 0.909 | 0.764 | +0.144 | 0.916 (132) |
| Disorders of penis | Genitourinary | 19 | 4.0 | 0.864 | 0.845 | +0.019 | 0.715 (491) |
| Benign prostatic hyperplasia | Genitourinary | 168 | 4.4 | 0.863 | 0.861 | +0.001 | 0.836 (2209) |
| Heart failure | Cardiovascular | 37 | 5.0 | 0.862 | 0.816 | +0.046 | 0.698 (141) |
| Atrioventricular block, first degree | Cardiovascular | 36 | 4.5 | 0.857 | 0.840 | +0.016 | 0.629 (165) |
| **Hyperkalaemia** | Endocrine/Metab | 21 | 4.9 | 0.850 | 0.749 | +0.100 | 0.687 (90) |
| **Alzheimer’s disease** | Neurological | 15 | 5.0 | 0.846 | 0.758 | +0.088 | 0.569 (16) |
| **Other diseases of biliary tract** | Gastrointestinal | 15 | 4.2 | 0.837 | 0.774 | +0.063 | 0.542 (164) |
| **Pressure ulcer** | Dermatological | 29 | 4.8 | 0.836 | 0.727 | +0.109 | 0.530 (29) |
| **Hydrocele** | Genitourinary | 14 | 4.1 | 0.836 | 0.765 | +0.071 | 0.696 (248) |
| Aortic valve disorders | Cardiovascular | 18 | 4.5 | 0.833 | 0.805 | +0.028 | 0.631 (125) |
| **Non-pressure chronic ulcer** | Dermatological | 16 | 5.2 | 0.831 | 0.728 | +0.103 | 0.632 (113) |
| **Repeated falls\*** | Neurological | 97 | 4.9 | 0.827 | 0.758 | +0.069 | 0.628 (47) |
| Aortic stenosis | Cardiovascular | 39 | 4.5 | 0.827 | 0.833 | -0.006 | 0.638 (238) |
| Atrioventricular block, complete | Cardiovascular | 13 | 5.4 | 0.826 | 0.794 | +0.032 | 0.622 (94) |
| **Acidosis** | Endocrine/Metab | 25 | 4.8 | 0.825 | 0.770 | +0.055 | 0.547 (69) |
| | | Cases | Lead (yr) | $C$ | $C_{\text{demo}}$ | $\Delta C$ | AUROC ($n$) |
| Occlusion and stenosis of cerebral arterie | Cardiovascular | 12 | 4.9 | 0.824 | 0.792 | +0.032 | 0.578 (50) |
| **Hemiplegia and hemiparesis** | Neurological | 23 | 5.3 | 0.822 | 0.676 | +0.145 | 0.626 (152) |
| **Epilepsy** | Neurological | 15 | 4.5 | 0.819 | 0.706 | +0.113 | 0.612 (467) |
| Obesity | Endocrine/Metab | 222 | 4.4 | 0.819 | 0.869 | -0.050 | 0.834 (1641) |
| Diverticulum of bladder | Genitourinary | 11 | 4.4 | 0.812 | 0.800 | +0.012 | 0.619 (144) |
| **Fluid overload** | Endocrine/Metab | 12 | 6.2 | 0.811 | 0.641 | +0.171 | 0.518 (29) |
| Diseases of the pancreas | Gastrointestinal | 12 | 4.3 | 0.811 | 0.766 | +0.045 | 0.520 (65) |
| **Chronic obstructive pulmonary disease** | Respiratory | 92 | 4.3 | 0.808 | 0.698 | +0.110 | 0.785 (890) |
| Uterine/Uterovaginal prolapse | Genitourinary | 50 | 3.0 | 0.808 | 0.804 | +0.004 | 0.765 (2361) |
| Sequelae of cancer | Neoplasms | 27 | 4.0 | 0.806 | 0.782 | +0.024 | 0.684 (477) |
| Bladder neck obstruction | Genitourinary | 10 | 3.4 | 0.806 | 0.807 | -0.002 | 0.700 (384) |
| **End stage renal disease** | Genitourinary | 11 | 5.0 | 0.805 | 0.540 | +0.265 | 0.714 (202) |
| **Pneumonia** | Respiratory | 60 | 4.3 | 0.805 | 0.738 | +0.067 | 0.603 (450) |
| Sleep apnoea | Neurological | 57 | 3.8 | 0.805 | 0.804 | +0.001 | 0.731 (838) |
| Inguinal hernia | Gastrointestinal | 128 | 3.4 | 0.803 | 0.793 | +0.011 | 0.780 (3380) |
| **Acute kidney failure** | Genitourinary | 136 | 4.9 | 0.802 | 0.747 | +0.055 | 0.691 (379) |
| **Thyrotoxicosis** | Endocrine/Metab | 10 | 4.4 | 0.801 | 0.687 | +0.115 | 0.519 (281) |
| Dementias | Neurological | 27 | 5.5 | 0.801 | 0.773 | +0.028 | 0.632 (21) |
| **Left heart failure** | Cardiovascular | 52 | 4.3 | 0.801 | 0.745 | +0.055 | 0.719 (400) |
| Cerebral degeneration | Neurological | 16 | 6.0 | 0.800 | 0.777 | +0.022 | 0.541 (36) |
| **Aspiration pneumonia** | Respiratory | 20 | 5.0 | 0.800 | 0.669 | +0.131 | 0.562 (31) |
| Arterial embolism and thrombosis | Cardiovascular | 14 | 4.4 | 0.798 | 0.778 | +0.019 | 0.546 (130) |
| Pulmonary hypertension | Cardiovascular | 14 | 5.0 | 0.798 | 0.798 | +0.000 | 0.567 (38) |
| Atrophic vaginitis | Genitourinary | 13 | 3.6 | 0.797 | 0.834 | -0.037 | 0.735 (192) |
| Abdominal aortic aneurysm | Cardiovascular | 18 | 4.3 | 0.797 | 0.765 | +0.032 | 0.652 (116) |
| **Cerebral infarction** | Cardiovascular | 33 | 4.3 | 0.793 | 0.736 | +0.057 | 0.620 (305) |
| **Pain in hip** | Musculoskeletal | 27 | 3.9 | 0.789 | 0.623 | +0.166 | 0.605 (269) |
| Ischaemic heart disease | Cardiovascular | 128 | 3.9 | 0.787 | 0.770 | +0.017 | 0.748 (2174) |
| **Symptoms and signs involving cognitive fun** | Neurological | 19 | 4.8 | 0.785 | 0.727 | +0.058 | 0.458 (42) |
| Presence of cardiac device | Cardiovascular | 41 | 4.9 | 0.785 | 0.775 | +0.010 | 0.672 (403) |
| Type 2 diabetes | Endocrine/Metab | 159 | 3.8 | 0.777 | 0.751 | +0.026 | 0.814 (2662) |
| Macular degeneration | Sense organs | 77 | 4.9 | 0.777 | 0.779 | -0.002 | 0.557 (431) |
| **Ventricular premature depolarisation\*** | Cardiovascular | 15 | 4.5 | 0.777 | 0.568 | +0.209 | 0.483 (72) |
| **Pleurisy** | Respiratory | 17 | 5.7 | 0.775 | 0.719 | +0.056 | 0.491 (171) |
| Coronary atherosclerosis | Cardiovascular | 86 | 4.0 | 0.774 | 0.750 | +0.024 | 0.752 (1387) |
| Chronic kidney disease | Genitourinary | 151 | 4.7 | 0.772 | 0.762 | +0.010 | 0.699 (556) |
| Aortic insufficiency | Cardiovascular | 24 | 4.7 | 0.772 | 0.792 | -0.021 | 0.608 (179) |
| **Urinary incontinence and enuresis** | Genitourinary | 54 | 3.8 | 0.769 | 0.676 | +0.094 | 0.607 (596) |
| Orthostatic hypotension | Cardiovascular | 50 | 5.2 | 0.769 | 0.733 | +0.035 | 0.628 (145) |
| **Complications and ill-defined descriptions** | Cardiovascular | 72 | 4.6 | 0.768 | 0.710 | +0.058 | 0.656 (252) |
| **All-cause mortality** | Mortality | 213 | 5.0 | 0.767 | 0.701 | +0.066 | — |
| **Obstruction of bile duct** | Gastrointestinal | 14 | 4.1 | 0.767 | 0.510 | +0.257 | 0.508 (97) |
| **Disorders of magnesium metabolism** | Endocrine/Metab | 18 | 5.3 | 0.767 | 0.636 | +0.131 | 0.676 (27) |
| Urethral stricture | Genitourinary | 22 | 4.0 | 0.766 | 0.770 | -0.005 | 0.535 (624) |
| **Epidermal thickening** | Dermatological | 12 | 4.8 | 0.764 | 0.600 | +0.164 | 0.483 (132) |
| Testicular limpoma | Neoplasms | 11 | 4.1 | 0.764 | 0.789 | -0.025 | 0.712 (160) |
| Retention of urine | Genitourinary | 129 | 4.6 | 0.764 | 0.757 | +0.007 | 0.660 (1088) |
| **Altered mental status, unspecified** | Neurological | 41 | 5.2 | 0.764 | 0.708 | +0.056 | 0.579 (198) |
| Myocardial infarction | Cardiovascular | 104 | 4.3 | 0.763 | 0.746 | +0.017 | 0.741 (953) |

| Disease | Category | Incident |  |  |  |  | Prevalent |  |
| --- | --- | --- | --- | --- | --- | --- | --- | --- |
| | | Cases | Lead (yr) | $C$ | $C_{\text{demo}}$ | $\Delta C$ | AUROC ($n$) | |
| **Other mental and behavioural problems** | Mental | 18 | 5.6 | 0.763 | 0.507 | +0.255 | 0.482 (160) |  |
| Leiomyoma of uterus | Neoplasms | 31 | 3.6 | 0.763 | 0.797 | -0.034 | 0.725 (2561) |  |
| Haematemesis | Gastrointestinal | 10 | 4.3 | 0.762 | 0.723 | +0.039 | 0.505 (223) |  |
| **Abnormality of gait and mobility** | Neurological | 60 | 4.8 | 0.760 | 0.682 | +0.077 | 0.598 (111) |  |
| **Interstitial pulmonary diseases** | Respiratory | 26 | 4.3 | 0.759 | 0.665 | +0.094 | 0.597 (99) |  |
| **Diplopia** | Sense organs | 17 | 4.9 | 0.759 | 0.528 | +0.230 | 0.529 (126) |  |
| Pneumothorax and air leak | Respiratory | 13 | 4.8 | 0.758 | 0.735 | +0.024 | 0.564 (93) |  |
| Bradycardia | Cardiovascular | 53 | 4.6 | 0.758 | 0.748 | +0.011 | 0.585 (476) |  |
| Tricuspid valve disorders | Cardiovascular | 53 | 5.2 | 0.758 | 0.718 | +0.040 | 0.579 (166) |  |
| Cholangitis | Gastrointestinal | 15 | 5.1 | 0.755 | 0.729 | +0.027 | 0.493 (77) |  |
| Hammer toe | Musculoskeletal | 13 | 3.2 | 0.754 | 0.707 | +0.047 | 0.602 (362) |  |
| **Scoliosis** | Musculoskeletal | 16 | 3.8 | 0.753 | 0.634 | +0.119 | 0.680 (70) |  |
| **Peripheral vascular disease NOS** | Cardiovascular | 26 | 4.4 | 0.752 | 0.678 | +0.074 | 0.726 (272) |  |
| Other disorders of male genital organs | Genitourinary | 27 | 4.2 | 0.751 | 0.807 | -0.056 | 0.727 (566) |  |
| Acute myocardial infarction | Cardiovascular | 60 | 4.4 | 0.750 | 0.741 | +0.009 | 0.703 (1259) |  |
| **Persistent atrial fibrillation\*** | Cardiovascular | 18 | 5.5 | 0.750 | 0.586 | +0.164 | - |  |
| Fatty liver disease (FLD) | Gastrointestinal | 42 | 5.0 | 0.750 | 0.706 | +0.044 | 0.646 (251) |  |
| Gout | Musculoskeletal | 53 | 4.6 | 0.750 | 0.760 | -0.010 | 0.750 (252) |  |
| **Pain in shoulder** | Musculoskeletal | 20 | 5.3 | 0.750 | 0.432 | +0.318 | 0.516 (159) |  |
| **Hypokalaemia** | Endocrine/Metab | 47 | 4.9 | 0.750 | 0.694 | +0.055 | 0.654 (148) |  |
| **Disorders of the retina** | Sense organs | 25 | 4.4 | 0.750 | 0.276 | +0.474 | 0.533 (218) |  |
| Volume depletion | Endocrine/Metab | 63 | 5.1 | 0.749 | 0.723 | +0.026 | 0.481 (292) |  |
| Stress incontinence | Genitourinary | 15 | 2.2 | 0.748 | 0.800 | -0.051 | 0.705 (1163) |  |
| Atrial fibrillation and flutter | Cardiovascular | 183 | 4.4 | 0.748 | 0.727 | +0.021 | 0.746 (2389) |  |
| Nuclear cataract | Sense organs | 168 | 4.7 | 0.747 | 0.739 | +0.008 | 0.639 (845) |  |
| **Cystitis** | Genitourinary | 11 | 4.1 | 0.747 | 0.595 | +0.152 | 0.522 (349) |  |
| Other cerebrovascular disease | Cardiovascular | 54 | 5.7 | 0.746 | 0.729 | +0.018 | 0.559 (147) |  |
| **Phlebitis and thrombophlebitis** | Cardiovascular | 33 | 4.3 | 0.746 | 0.612 | +0.134 | 0.576 (621) |  |
| **Emphysema** | Respiratory | 16 | 5.1 | 0.744 | 0.532 | +0.212 | 0.748 (149) |  |
| Gastrointestinal angiodysplasia | Gastrointestinal | 19 | 5.1 | 0.744 | 0.707 | +0.037 | 0.539 (91) |  |
| Cataract | Sense organs | 383 | 4.7 | 0.743 | 0.743 | +0.000 | 0.690 (3315) |  |
| **Cardiac arrhythmia and conduction disorder** | Cardiovascular | 24 | 4.7 | 0.743 | 0.661 | +0.082 | 0.559 (253) |  |
| Epistaxis | Respiratory | 18 | 3.1 | 0.743 | 0.757 | -0.014 | 0.538 (411) |  |
| Left bundle branch block | Cardiovascular | 34 | 4.3 | 0.742 | 0.734 | +0.007 | 0.611 (286) |  |
| **Ventral hernia** | Gastrointestinal | 13 | 3.4 | 0.741 | 0.615 | +0.127 | 0.524 (616) |  |
| **Acute lower respiratory infection** | Respiratory | 64 | 4.0 | 0.738 | 0.644 | +0.094 | 0.584 (974) |  |
| Phobic disorders | Mental | 16 | 4.4 | 0.738 | 0.736 | +0.002 | 0.606 (102) |  |
| Sensorineural hearing loss | Sense organs | 30 | 5.7 | 0.737 | 0.737 | +0.000 | 0.427 (51) |  |
| Fracture of femur | Musculoskeletal | 34 | 4.9 | 0.737 | 0.758 | -0.021 | 0.611 (353) |  |
| Disorders of uterus, except cervix | Genitourinary | 16 | 4.0 | 0.737 | 0.697 | +0.039 | 0.669 (564) |  |
| Open angle glaucoma | Sense organs | 19 | 5.5 | 0.736 | 0.751 | -0.015 | 0.564 (195) |  |
| **Haemorrhagic stroke** | Cardiovascular | 14 | 4.8 | 0.735 | 0.670 | +0.066 | 0.585 (213) |  |
| Hyperlipidaemia | Endocrine/Metab | 70 | 4.6 | 0.734 | 0.706 | +0.028 | 0.654 (790) |  |
| Effusion of joint | Musculoskeletal | 17 | 5.4 | 0.734 | 0.773 | -0.039 | 0.544 (224) |  |
| Corneal degenerations | Sense organs | 29 | 5.6 | 0.733 | 0.738 | -0.005 | 0.706 (19) |  |
| **Benign neoplasms of the ovary** | Neoplasms | 10 | 4.5 | 0.732 | 0.643 | +0.089 | 0.672 (343) |  |
| Atherosclerosis | Cardiovascular | 145 | 4.2 | 0.732 | 0.738 | -0.006 | 0.760 (3125) |  |
| **Viral pneumonia** | Respiratory | 24 | 5.7 | 0.731 | 0.677 | +0.054 | 0.470 (13) |  |
| | | Cases | Lead (yr) | $C$ | $C_{\text{demo}}$ | $\Delta C$ | AUROC ($n$) |
| Cardiomegaly | Cardiovascular | 68 | 4.6 | 0.731 | 0.704 | +0.026 | 0.663 (371) |
| **Vitamin B group deficiency** | Endocrine/Metab | 26 | 4.5 | 0.730 | 0.614 | +0.116 | 0.584 (107) |
| **Spondylosis** | Musculoskeletal | 37 | 4.4 | 0.729 | 0.656 | +0.073 | 0.630 (440) |
| **Abnormal findings on diagnostic imaging of** | Respiratory | 38 | 5.0 | 0.729 | 0.676 | +0.053 | 0.493 (279) |
| Hyposmolality and/or hyponatraemia | Endocrine/Metab | 62 | 5.2 | 0.728 | 0.682 | +0.045 | 0.680 (214) |
| **Respiratory failure** | Respiratory | 32 | 4.7 | 0.726 | 0.647 | +0.080 | 0.656 (120) |
| Other disorders of bone | Musculoskeletal | 50 | 4.8 | 0.724 | 0.721 | +0.003 | 0.565 (227) |
| Rheumatoid arthritis | Musculoskeletal | 31 | 3.5 | 0.723 | 0.711 | +0.012 | 0.701 (352) |
| **Carcinoma in situ of skin** | Neoplasms | 17 | 3.6 | 0.722 | 0.663 | +0.059 | 0.605 (143) |
| **Ascites** | Gastrointestinal | 22 | 4.6 | 0.720 | 0.643 | +0.077 | 0.545 (134) |
| **Myalgia** | Musculoskeletal | 35 | 4.4 | 0.720 | 0.661 | +0.059 | 0.662 (151) |
| Essential hypertension | Cardiovascular | 646 | 3.9 | 0.719 | 0.703 | +0.015 | 0.775 (13468) |
| Pathological fracture | Musculoskeletal | 17 | 3.7 | 0.717 | 0.684 | +0.033 | 0.551 (82) |
| Intestinal perforation | Gastrointestinal | 13 | 5.3 | 0.717 | 0.676 | +0.041 | 0.527 (177) |
| **Cardiac murmurs** | Cardiovascular | 22 | 4.4 | 0.716 | 0.658 | +0.058 | 0.497 (199) |
| Osteoarthritis of more than one site | Musculoskeletal | 70 | 4.4 | 0.716 | 0.703 | +0.013 | 0.697 (585) |
| Urinary tract infection | Genitourinary | 113 | 4.6 | 0.715 | 0.715 | +0.001 | 0.564 (1851) |
| Vitreous opacities | Sense organs | 21 | 5.2 | 0.713 | 0.696 | +0.017 | 0.520 (139) |
| Nonspecific low blood-pressure reading | Cardiovascular | 22 | 4.5 | 0.712 | 0.717 | -0.005 | 0.588 (162) |
| Cyst of kidney | Genitourinary | 36 | 4.2 | 0.712 | 0.674 | +0.038 | 0.554 (206) |
| Mucous polyp of cervix | Genitourinary | 10 | 3.5 | 0.711 | 0.783 | -0.072 | 0.660 (783) |
| Pure hypercholesterolaemia | Endocrine/Metab | 337 | 3.9 | 0.710 | 0.702 | +0.009 | 0.728 (5613) |
| Spinal stenosis | Musculoskeletal | 56 | 4.4 | 0.710 | 0.713 | -0.004 | 0.663 (590) |
| Umbilical hernia | Gastrointestinal | 27 | 3.7 | 0.710 | 0.726 | -0.016 | 0.648 (695) |
| Hallux valgus (Bunion) | Musculoskeletal | 28 | 3.2 | 0.710 | 0.677 | +0.032 | 0.677 (1421) |
| Polyp of corpus uteri | Genitourinary | 39 | 3.8 | 0.710 | 0.765 | -0.055 | 0.713 (1870) |
| **Anorexia** | Endocrine/Metab | 15 | 4.7 | 0.708 | 0.628 | +0.080 | 0.532 (107) |
| Fracture of upper limb | Musculoskeletal | 55 | 4.0 | 0.708 | 0.687 | +0.021 | 0.585 (1480) |
| **Pulmonary embolism** | Cardiovascular | 38 | 4.5 | 0.708 | 0.622 | +0.086 | 0.574 (524) |
| Hypotension | Cardiovascular | 84 | 4.7 | 0.707 | 0.659 | +0.048 | 0.583 (517) |
| Lobar pneumonia* | Respiratory | 103 | 4.3 | 0.705 | 0.669 | +0.036 | 0.573 (710) |
| **Amblyopia** | Sense organs | 14 | 4.3 | 0.705 | 0.650 | +0.055 | 0.541 (95) |
| **Back pain** | Musculoskeletal | 63 | 4.5 | 0.705 | 0.620 | +0.085 | 0.538 (358) |
| Transient cerebral ischaemic attack | Cardiovascular | 13 | 5.2 | 0.705 | 0.739 | -0.034 | 0.578 (286) |
| **Carpal tunnel syndrome** | Neurological | 50 | 3.2 | 0.704 | 0.588 | +0.117 | 0.680 (1858) |
| **Fracture of vertebrae** | Musculoskeletal | 16 | 4.3 | 0.704 | 0.572 | +0.133 | 0.530 (224) |
| Raynaud's syndrome | Cardiovascular | 20 | 4.1 | 0.704 | 0.663 | +0.041 | 0.606 (212) |
| Sepsis | Infections | 101 | 4.4 | 0.704 | 0.658 | +0.046 | 0.575 (411) |
| Polyneuropathies | Neurological | 28 | 4.7 | 0.704 | 0.662 | +0.042 | 0.613 (161) |
| **Aphasia and dysphasia** | Neurological | 15 | 4.9 | 0.702 | 0.643 | +0.059 | 0.678 (73) |
| **Vitamin D deficiency** | Endocrine/Metab | 38 | 5.2 | 0.702 | 0.554 | +0.148 | 0.555 (76) |
| Bacterial infections | Infections | 94 | 4.3 | 0.702 | 0.675 | +0.026 | 0.587 (1877) |
| Astigmatism | Sense organs | 76 | 5.8 | 0.701 | 0.701 | +0.000 | 0.527 (38) |
| Polymyalgia rheumatica | Musculoskeletal | 31 | 4.5 | 0.701 | 0.691 | +0.010 | 0.604 (201) |
| Constipation | Gastrointestinal | 194 | 4.6 | 0.700 | 0.659 | +0.042 | 0.582 (1585) |
| **Infection of the skin** | Dermatological | 12 | 3.3 | 0.700 | 0.611 | +0.089 | 0.481 (204) |
| **Pulmonary collapse** | Respiratory | 55 | 5.0 | 0.699 | 0.648 | +0.051 | 0.595 (242) |
| Abnormal findings on diagnostic imaging of | Gastrointestinal | 37 | 4.0 | 0.699 | 0.690 | +0.009 | 0.533 (301) |
| **Faecal incontinence and encopresis** | Gastrointestinal | 31 | 4.7 | 0.699 | 0.585 | +0.114 | 0.644 (261) |
| **Functional dyspepsia** | Gastrointestinal | 45 | 3.4 | 0.699 | 0.579 | +0.120 | 0.561 (2424) |
| **Heart valve replaced** | Cardiovascular | 19 | 4.6 | 0.697 | 0.634 | +0.063 | 0.688 (236) |
| Postmenopausal bleeding | Genitourinary | 29 | 3.1 | 0.696 | 0.773 | -0.077 | 0.738 (1971) |
| Palmar fascial fibromatosis | Musculoskeletal | 36 | 3.6 | 0.696 | 0.662 | +0.034 | 0.674 (472) |
| | | Cases | Lead (yr) | $C$ | $C_{\text{demo}}$ | $\Delta C$ | AUROC ($n$) |
| Paroxysmal atrial fibrillation* | Cardiovascular | 83 | 5.0 | 0.695 | 0.681 | +0.014 | – |
| Osteoarthritis | Musculoskeletal | 424 | 4.0 | 0.695 | 0.686 | +0.008 | 0.715 (6879) |
| **Angle-Closure Glaucoma** | Sense organs | 18 | 4.5 | 0.694 | 0.580 | +0.114 | 0.538 (137) |
| Hydronephrosis | Genitourinary | 38 | 4.8 | 0.694 | 0.646 | +0.048 | 0.518 (280) |
| Osteophyte* | Musculoskeletal | 34 | 4.4 | 0.694 | 0.660 | +0.034 | 0.578 (575) |
| Actinic keratosis | Dermatological | 51 | 4.2 | 0.694 | 0.647 | +0.046 | 0.534 (453) |
| Hearing impairment | Sense organs | 106 | 4.9 | 0.694 | 0.698 | -0.004 | 0.588 (535) |
| **Bursitis** | Musculoskeletal | 24 | 3.3 | 0.692 | 0.482 | +0.210 | 0.558 (390) |
| **Dizziness and giddiness** | Neurological | 76 | 4.6 | 0.691 | 0.629 | +0.062 | 0.554 (741) |
| **Cellulitis and abscess** | Dermatological | 98 | 4.3 | 0.691 | 0.635 | +0.056 | 0.605 (1546) |
| **Blood in stool** | Gastrointestinal | 31 | 3.5 | 0.691 | 0.638 | +0.053 | 0.548 (393) |
| Other diseases of stomach and duodenum | Gastrointestinal | 26 | 4.2 | 0.690 | 0.700 | -0.009 | 0.560 (491) |
| Gallstones | Gastrointestinal | 97 | 4.4 | 0.690 | 0.645 | +0.045 | 0.644 (2699) |
| **Disorders of calcium metabolism** | Endocrine/Metab | 27 | 5.3 | 0.690 | 0.581 | +0.109 | 0.583 (125) |
| Inflammatory spondylopathies | Musculoskeletal | 20 | 4.0 | 0.689 | 0.654 | +0.035 | 0.530 (175) |
| Renal failure | Genitourinary | 12 | 4.0 | 0.688 | 0.682 | +0.007 | 0.578 (153) |
| Pleural effusion | Respiratory | 94 | 4.6 | 0.688 | 0.685 | +0.003 | 0.584 (604) |
| **Myelopathies** | Neurological | 12 | 4.4 | 0.688 | 0.595 | +0.093 | 0.593 (129) |
| Low back pain | Musculoskeletal | 43 | 3.8 | 0.688 | 0.702 | -0.014 | 0.593 (960) |
| Bronchiectasis | Respiratory | 38 | 5.0 | 0.688 | 0.655 | +0.033 | 0.599 (282) |
| **Anxiety and anxiety disorders** | Mental | 126 | 4.5 | 0.687 | 0.577 | +0.110 | 0.624 (577) |
| Right bundle branch block | Cardiovascular | 37 | 4.6 | 0.687 | 0.679 | +0.008 | 0.587 (245) |
| Other respiratory disorders | Respiratory | 32 | 4.5 | 0.686 | 0.639 | +0.047 | 0.522 (318) |
| **Major depressive disorder** | Mental | 159 | 4.1 | 0.685 | 0.601 | +0.084 | 0.728 (1673) |
| Rotator cuff tear or rupture | Musculoskeletal | 26 | 3.1 | 0.683 | 0.633 | +0.049 | 0.614 (670) |
| Viral infections | Infections | 87 | 5.3 | 0.682 | 0.666 | +0.015 | 0.543 (800) |
| Anaemia | Blood/Immune | 161 | 4.4 | 0.681 | 0.643 | +0.038 | 0.585 (1942) |
| **Abnormal glucose** | Endocrine/Metab | 24 | 5.9 | 0.681 | 0.621 | +0.060 | 0.533 (58) |
| Hypothyroidism (not specified as secondary) | Endocrine/Metab | 164 | 3.8 | 0.680 | 0.662 | +0.018 | 0.670 (2617) |
| **Trigger finger** | Musculoskeletal | 20 | 3.7 | 0.679 | 0.584 | +0.095 | 0.581 (538) |
| Fracture of bones in thorax | Musculoskeletal | 16 | 3.7 | 0.679 | 0.638 | +0.041 | 0.582 (257) |
| **Tachycardia** | Cardiovascular | 34 | 5.0 | 0.677 | 0.512 | +0.165 | 0.554 (295) |
| Spondylolisthesis | Musculoskeletal | 23 | 4.1 | 0.676 | 0.673 | +0.002 | 0.644 (238) |
| Diverticular disease | Gastrointestinal | 106 | 4.8 | 0.673 | 0.651 | +0.022 | 0.633 (1105) |
| Angina pectoris | Cardiovascular | 105 | 3.9 | 0.673 | 0.660 | +0.013 | 0.727 (2826) |
| Tricuspid valve insufficiency* | Cardiovascular | 14 | 5.0 | 0.672 | 0.669 | +0.004 | 0.637 (49) |
| **Nonspecific abnormal results of function s** | Gastrointestinal | 56 | 4.2 | 0.672 | 0.599 | +0.072 | 0.511 (532) |
| Other disorders of bladder | Genitourinary | 81 | 4.3 | 0.672 | 0.660 | +0.012 | 0.610 (1311) |
| Dyspnoea | Respiratory | 81 | 4.4 | 0.670 | 0.624 | +0.046 | 0.593 (868) |
| **Enthesopathy/Enthesitis/Tendinopathy** | Musculoskeletal | 13 | 4.2 | 0.664 | 0.455 | +0.209 | 0.524 (191) |
| **Pain in knee** | Musculoskeletal | 14 | 4.6 | 0.663 | 0.603 | +0.060 | 0.560 (377) |
| Gastric ulcer | Gastrointestinal | 43 | 3.7 | 0.662 | 0.632 | +0.030 | 0.560 (656) |
| **Peritonitis** | Gastrointestinal | 16 | 5.2 | 0.661 | 0.561 | +0.100 | 0.538 (159) |
| Malignant neoplasm of the skin | Neoplasms | 168 | 4.1 | 0.661 | 0.664 | -0.003 | 0.597 (2347) |
| **Cervicalgia** | Musculoskeletal | 11 | 4.0 | 0.660 | 0.574 | +0.086 | 0.580 (151) |
| Other symptoms/disorders or the urinary sy | Genitourinary | 22 | 3.9 | 0.659 | 0.676 | -0.017 | 0.581 (857) |
| **Thrombocytopenia** | Blood/Immune | 24 | 4.6 | 0.657 | 0.521 | +0.136 | 0.455 (151) |
| Diaphragmatic hernia | Gastrointestinal | 253 | 4.0 | 0.657 | 0.620 | +0.037 | 0.653 (4902) |

| Disease | Category | Incident |  |  |  |  | Prevalent |  |
| --- | --- | --- | --- | --- | --- | --- | --- | --- |
| | | Cases | Lead (yr) | $C$ | $C_{\text{demo}}$ | $\Delta C$ | AUROC ($n$) | |
| Cancer (solid tumour, excluding BCC) | Neoplasms | 515 | 4.3 | 0.656 | 0.655 | +0.002 | 0.625 (8392) |  |
| Haematuria | Genitourinary | 77 | 4.1 | 0.655 | 0.636 | +0.018 | 0.568 (3114) |  |
| **Enlargement of lymph nodes** | Blood/Immune | 35 | 4.4 | 0.655 | 0.592 | +0.062 | 0.557 (384) |  |
| Mitral valve insufficiency | Cardiovascular | 50 | 4.6 | 0.654 | 0.649 | +0.005 | 0.608 (372) |  |
| **Dermatitis** | Dermatological | 40 | 4.9 | 0.653 | 0.440 | +0.214 | 0.538 (300) |  |
| **Pain in joint** | Musculoskeletal | 17 | 4.6 | 0.652 | 0.519 | +0.133 | 0.531 (378) |  |
| Supraventricular tachycardia | Cardiovascular | 21 | 4.7 | 0.652 | 0.636 | +0.016 | 0.529 (464) |  |
| Secondary malignant neoplasm | Neoplasms | 140 | 4.6 | 0.650 | 0.636 | +0.015 | 0.562 (927) |  |
| **Hyperparathyroidism** | Endocrine/Metab | 11 | 3.9 | 0.650 | 0.590 | +0.060 | 0.545 (127) |  |
| **Disorders of phosphorus metabolism** | Endocrine/Metab | 22 | 5.6 | 0.650 | 0.535 | +0.115 | 0.652 (21) |  |
| Frequency of urination and polyuria | Genitourinary | 16 | 3.9 | 0.649 | 0.641 | +0.008 | 0.543 (717) |  |
| Disorder of patella | Musculoskeletal | 12 | 2.9 | 0.646 | 0.636 | +0.010 | 0.540 (400) |  |
| Other disorders of the intestines | Gastrointestinal | 48 | 4.7 | 0.646 | 0.626 | +0.020 | 0.542 (472) |  |
| Other skin and subcutaneous disorders | Dermatological | 63 | 4.3 | 0.646 | 0.597 | +0.049 | 0.520 (1076) |  |
| **Disturbances of skin sensation** | Neurological | 17 | 3.4 | 0.646 | 0.475 | +0.171 | 0.516 (255) |  |
| **Fungal infections** | Infections | 42 | 4.7 | 0.645 | 0.594 | +0.051 | 0.527 (239) |  |
| Spinal disc displacement (herniation) | Musculoskeletal | 52 | 4.1 | 0.645 | 0.659 | -0.015 | 0.636 (890) |  |
| **Other disorders of liver** | Gastrointestinal | 33 | 4.8 | 0.645 | 0.571 | +0.073 | 0.576 (259) |  |
| Mononeuritis of lower limb | Neurological | 11 | 3.4 | 0.644 | 0.706 | -0.062 | 0.557 (346) |  |
| Cough | Respiratory | 40 | 4.0 | 0.644 | 0.607 | +0.036 | 0.566 (500) |  |
| Impingement syndrome of shoulder* | Musculoskeletal | 37 | 2.6 | 0.644 | 0.635 | +0.009 | 0.546 (980) |  |
| Varicose veins of lower extremities | Cardiovascular | 39 | 3.5 | 0.642 | 0.646 | -0.004 | 0.604 (2252) |  |
| Duodenitis | Gastrointestinal | 54 | 4.1 | 0.640 | 0.630 | +0.010 | 0.572 (1385) |  |
| Irritable bowel syndrome | Gastrointestinal | 74 | 4.5 | 0.639 | 0.636 | +0.004 | 0.589 (918) |  |
| Intestinal or peritoneal adhesions | Gastrointestinal | 73 | 4.6 | 0.639 | 0.618 | +0.021 | 0.538 (705) |  |
| Glaucoma | Sense organs | 66 | 4.0 | 0.637 | 0.617 | +0.021 | 0.615 (481) |  |
| Migraine | Neurological | 35 | 4.1 | 0.637 | 0.638 | -0.001 | 0.604 (513) |  |
| Abnormal weight loss | Endocrine/Metab | 109 | 4.2 | 0.637 | 0.622 | +0.014 | 0.546 (827) |  |
| Barrett's oesophagus | Gastrointestinal | 41 | 4.0 | 0.636 | 0.636 | +0.000 | 0.592 (444) |  |
| Iron deficiency | Endocrine/Metab | 154 | 4.7 | 0.636 | 0.595 | +0.041 | 0.563 (1420) |  |
| **Rectal prolapse** | Gastrointestinal | 14 | 2.7 | 0.636 | 0.569 | +0.067 | 0.579 (214) |  |
| Ptosis of eyelid | Sense organs | 18 | 4.5 | 0.636 | 0.605 | +0.030 | 0.539 (280) |  |
| Diverticula of colon | Gastrointestinal | 388 | 3.9 | 0.636 | 0.626 | +0.010 | 0.648 (4346) |  |
| **Secondary hypothyroidism** | Endocrine/Metab | 19 | 4.9 | 0.634 | 0.525 | +0.109 | 0.568 (230) |  |
| **Intervertebral disc disorder** | Musculoskeletal | 40 | 4.5 | 0.629 | 0.534 | +0.095 | 0.579 (835) |  |
| **Flatulence, eructation, and gas pain** | Gastrointestinal | 20 | 4.7 | 0.627 | 0.455 | +0.172 | 0.503 (297) |  |
| **Benign neoplasm of stomach** | Neoplasms | 108 | 4.3 | 0.626 | 0.560 | +0.066 | 0.569 (943) |  |
| Oesophageal obstruction (Stricture and sten) | Gastrointestinal | 23 | 4.5 | 0.624 | 0.612 | +0.012 | 0.557 (321) |  |
| Nasal polyps | Respiratory | 11 | 3.6 | 0.624 | 0.632 | -0.009 | 0.563 (667) |  |
| **Psoriasis** | Dermatological | 31 | 4.1 | 0.624 | 0.521 | +0.102 | 0.565 (283) |  |
| Kidney stone disease | Genitourinary | 37 | 3.7 | 0.623 | 0.622 | +0.000 | 0.612 (984) |  |
| Fracture of lower limb | Musculoskeletal | 16 | 2.8 | 0.622 | 0.717 | -0.095 | 0.534 (582) |  |
| Diseases of oesophagus | Gastrointestinal | 29 | 4.0 | 0.622 | 0.590 | +0.031 | 0.533 (267) |  |
| | | Cases | Lead (yr) | $C$ | $C_{\text{demo}}$ | $\Delta C$ | AUROC ($n$) |
| Rash and other nonspecific skin eruption | Dermatological | 23 | 4.0 | 0.621 | 0.641 | -0.020 | 0.498 (289) |
| **Myopia** | Sense organs | 38 | 4.3 | 0.621 | 0.442 | +0.179 | 0.493 (311) |
| **Aphagia and dysphagia** | Gastrointestinal | 104 | 4.2 | 0.620 | 0.548 | +0.072 | 0.555 (1048) |
| Gastro-oesophageal reflux disease | Gastrointestinal | 334 | 4.4 | 0.618 | 0.585 | +0.033 | 0.625 (4220) |
| **Acquired deformities of toe** | Musculoskeletal | 11 | 2.8 | 0.618 | 0.511 | +0.107 | 0.658 (391) |
| Lesions of mouth | Gastrointestinal | 12 | 3.9 | 0.615 | 0.589 | +0.026 | 0.510 (442) |
| **Benign neoplasm of the skin** | Neoplasms | 14 | 4.7 | 0.615 | 0.561 | +0.054 | 0.503 (957) |
| Corpus luteum cyst or haematoma | Genitourinary | 12 | 5.5 | 0.614 | 0.738 | -0.123 | 0.702 (999) |
| **Gastrointestinal haemorrhage** | Gastrointestinal | 110 | 4.1 | 0.614 | 0.542 | +0.071 | 0.525 (980) |
| **Ganglion cyst** | Musculoskeletal | 13 | 2.2 | 0.614 | 0.501 | +0.113 | 0.504 (670) |
| **Acute pancreatitis** | Gastrointestinal | 24 | 4.2 | 0.613 | 0.514 | +0.100 | 0.567 (339) |
| Oesophagitis | Gastrointestinal | 66 | 3.9 | 0.611 | 0.561 | +0.050 | 0.595 (1593) |
| **Localised swelling, mass and lump of skin** | Dermatological | 13 | 4.2 | 0.611 | 0.548 | +0.063 | 0.488 (416) |
| Haemo onc - by cell of origin | Neoplasms | 18 | 4.9 | 0.610 | 0.590 | +0.020 | 0.586 (210) |
| **Renal tubulo-interstitial diseases** | Genitourinary | 13 | 3.5 | 0.610 | 0.382 | +0.228 | 0.553 (169) |
| **Ulcerative colitis** | Gastrointestinal | 18 | 4.2 | 0.610 | 0.463 | +0.147 | 0.543 (677) |
| **Dental caries** | Gastrointestinal | 22 | 3.7 | 0.607 | 0.521 | +0.087 | 0.556 (644) |
| Duodenal ulcer | Gastrointestinal | 15 | 4.0 | 0.605 | 0.635 | -0.030 | 0.565 (429) |
| Seborrheic keratosis | Dermatological | 39 | 4.0 | 0.604 | 0.558 | +0.045 | 0.527 (702) |
| Abdominal pain | Gastrointestinal | 163 | 4.2 | 0.602 | 0.565 | +0.037 | 0.573 (3172) |
| **Radiculopathy** | Musculoskeletal | 16 | 4.1 | 0.601 | 0.504 | +0.098 | 0.556 (161) |
| **Psychoactive substance abuse** | Mental | 20 | 4.4 | 0.599 | 0.534 | +0.065 | 0.700 (322) |
| Benign neoplasm of the colon | Neoplasms | 295 | 4.2 | 0.599 | 0.581 | +0.018 | 0.625 (3502) |
| **Paresthesia of skin\*** | Neurological | 14 | 4.7 | 0.598 | 0.446 | +0.152 | 0.515 (254) |
| **Melanocytic nevi\*** | Neoplasms | 30 | 4.1 | 0.596 | 0.524 | +0.072 | 0.535 (1116) |
| Intestinal obstruction | Gastrointestinal | 22 | 3.7 | 0.596 | 0.621 | -0.024 | 0.559 (349) |
| Other disorders of the kidney and ureters | Genitourinary | 17 | 4.7 | 0.596 | 0.578 | +0.019 | 0.568 (170) |
| Neutropenia | Blood/Immune | 39 | 3.9 | 0.595 | 0.644 | -0.049 | 0.562 (432) |
| **Oesophageal ulcer** | Gastrointestinal | 15 | 3.0 | 0.595 | 0.512 | +0.083 | 0.606 (876) |
| Non-Hodgkin lymphoma | Neoplasms | 16 | 4.6 | 0.595 | 0.572 | +0.023 | 0.538 (238) |
| Abnormalities of breathing | Respiratory | 20 | 4.0 | 0.594 | 0.572 | +0.023 | 0.586 (537) |
| **Fracture of hand or wrist** | Musculoskeletal | 16 | 4.0 | 0.594 | 0.526 | +0.068 | 0.558 (455) |
| **Symptoms involving digestive system** | Gastrointestinal | 138 | 3.4 | 0.593 | 0.511 | +0.082 | 0.536 (2526) |
| **Asthma** | Respiratory | 188 | 3.6 | 0.592 | 0.516 | +0.076 | 0.655 (5073) |
| **Bacterial enteritis** | Gastrointestinal | 15 | 4.7 | 0.592 | 0.507 | +0.085 | 0.536 (335) |
| Other disorders of the eyelids | Sense organs | 20 | 4.5 | 0.592 | 0.559 | +0.033 | 0.502 (575) |
| **Retinal detachments and breaks** | Sense organs | 27 | 3.6 | 0.591 | 0.490 | +0.101 | 0.549 (592) |
| **Haemoptysis** | Respiratory | 15 | 3.5 | 0.590 | 0.527 | +0.063 | 0.559 (274) |
| Benign neoplasm of rectum and anus | Neoplasms | 120 | 3.8 | 0.589 | 0.588 | +0.001 | 0.579 (1921) |
| Haemorrhoids | Cardiovascular | 256 | 4.6 | 0.587 | 0.548 | +0.039 | – |
| Gastrointestinal inflammation | Gastrointestinal | 74 | 4.3 | 0.584 | 0.552 | +0.033 | 0.570 (2927) |
| Vomiting | Gastrointestinal | 122 | 4.6 | 0.583 | 0.541 | +0.042 | 0.589 (1797) |
| Fracture of pelvis | Musculoskeletal | 14 | 4.8 | 0.583 | 0.589 | -0.006 | 0.567 (143) |
| **Palpitations** | Cardiovascular | 39 | 4.5 | 0.582 | 0.404 | +0.178 | 0.568 (769) |
| Gastritis | Gastrointestinal | 225 | 3.7 | 0.581 | 0.574 | +0.007 | 0.607 (4266) |
| Acute Pharyngitis | Respiratory | 13 | 5.2 | 0.578 | 0.575 | +0.003 | 0.496 (146) |
| **Crohn's disease** | Gastrointestinal | 17 | 4.2 | 0.572 | 0.390 | +0.181 | 0.504 (365) |
| | | Cases | Lead (yr) | $C$ | $C_{\text{demo}}$ | $\Delta C$ | AUROC ($n$) |
| Disorders of the gallbladder | Gastrointestinal | 12 | 4.1 | 0.571 | 0.610 | -0.040 | 0.563 (240) |
| Sciatica | Musculoskeletal | 23 | 4.7 | 0.570 | 0.643 | -0.073 | 0.519 (205) |
| Other follicular disorders | Dermatological | 10 | 3.2 | 0.568 | 0.597 | -0.030 | 0.470 (369) |
| Abnormal intraocular pressure | Sense organs | 31 | 4.7 | 0.563 | 0.540 | +0.023 | 0.483 (197) |
| Deviated nasal septum | Respiratory | 12 | 2.5 | 0.561 | 0.525 | +0.037 | 0.579 (841) |
| Headache | Neurological | 61 | 4.2 | 0.554 | 0.518 | +0.036 | 0.589 (1299) |
| **Disease of anus and rectum** | Gastrointestinal | 42 | 4.1 | 0.554 | 0.490 | +0.064 | 0.530 (675) |
| Scar conditions and fibrosis of skin | Dermatological | 10 | 3.8 | 0.551 | 0.521 | +0.030 | 0.524 (517) |
| **Derangement of meniscus** | Musculoskeletal | 42 | 2.2 | 0.550 | 0.491 | +0.059 | 0.586 (2969) |
| **Coeliac disease** | Gastrointestinal | 15 | 4.3 | 0.545 | 0.493 | +0.053 | 0.542 (385) |
| Haemorrhage of rectum and anus | Gastrointestinal | 40 | 3.9 | 0.545 | 0.540 | +0.005 | 0.523 (2688) |
| Chronic rhinitis | Respiratory | 10 | 3.8 | 0.540 | 0.576 | -0.036 | 0.502 (165) |
| Nonspecific abnormal findings in stool con | Gastrointestinal | 50 | 4.5 | 0.534 | 0.545 | -0.011 | 0.536 (295) |
| **Adhesive capsulitis of shoulder** | Musculoskeletal | 11 | 3.0 | 0.531 | 0.426 | +0.105 | 0.575 (358) |
| **Diseases of tongue** | Gastrointestinal | 13 | 4.0 | 0.525 | 0.423 | +0.101 | 0.477 (158) |
| Haemangioma | Neoplasms | 10 | 3.8 | 0.512 | 0.547 | -0.035 | 0.462 (283) |
| Other symptoms and disorders of the soft t | Musculoskeletal | 13 | 5.2 | 0.508 | 0.480 | +0.028 | 0.497 (514) |
| Skull and face fracture | Musculoskeletal | 10 | 4.0 | 0.507 | 0.597 | -0.089 | 0.510 (243) |
| **Heartburn** | Gastrointestinal | 19 | 4.3 | 0.505 | 0.431 | +0.074 | 0.502 (383) |
| Chronic sinusitis | Respiratory | 19 | 3.0 | 0.504 | 0.569 | -0.065 | 0.502 (517) |
| Ulceration of the lower GI tract | Gastrointestinal | 13 | 4.4 | 0.500 | 0.645 | -0.145 | 0.498 (214) |
| Elevated blood pressure reading without di | Cardiovascular | 18 | 4.9 | 0.489 | 0.512 | -0.023 | 0.548 (259) |
| Other disorders of nose and nasal sinuses | Respiratory | 13 | 3.8 | 0.488 | 0.568 | -0.080 | 0.491 (549) |
| Pilar and trichodermal cyst | Dermatological | 14 | 3.0 | 0.460 | 0.580 | -0.120 | 0.512 (1004) |
| Lipoma of skin subcutaneous tissue | Neoplasms | 20 | 4.3 | 0.454 | 0.567 | -0.113 | 0.520 (1130) |
| **Type 1 diabetes** | Endocrine/Metab | 12 | 3.7 | 0.451 | 0.352 | +0.099 | 0.709 (383) |
| Visual disturbances | Sense organs | 17 | 4.1 | 0.434 | 0.469 | -0.035 | 0.494 (276) |
| Appendicitis | Gastrointestinal | 10 | 3.9 | 0.409 | 0.573 | -0.164 | 0.508 (174) |

**Supplementary Table 4.** Per-disease genetics fusion result (matched Standard PRS, European genetic ancestry, held-out test set). *C* values are leakage-safe out-of-fold concordance for the wrist embedding alone, the PRS alone, and the learned fusion [wrist hazard, PRS]. Δ*C* is fusion minus wrist with a 95% subject bootstrap interval; FDR is Benjamini–Hochberg across the 26 outcomes, and diseases in **bold** are significant at FDR *<* 0.05. The aggregate effect (+0.042, 95% CI +0.021 to +0.069) is the robust claim; per-<u>disease FDR is conservative on these incident-case counts.</u>

| Outcome | $n_{ev}$ | $C$ wrist | $C$ PRS | $C$ fusion | $\Delta C$ (95% CI) | FDR |
| --- | --- | --- | --- | --- | --- | --- |
| **Coeliac disease** | 12 | 0.586 | 0.868 | 0.869 | +0.283 (+0.093, +0.462) | 0.026 |
| Type 1 diabetes | 11 | 0.412 | 0.620 | 0.574 | +0.163 (-0.222, +0.489) | 0.557 |
| Glaucoma (open-angle) | 17 | 0.713 | 0.747 | 0.807 | +0.094 (+0.008, +0.177) | 0.097 |
| Hypertension (elevated BP) | 16 | 0.484 | 0.627 | 0.576 | +0.093 (-0.056, +0.246) | 0.416 |
| Crohn's disease | 15 | 0.586 | 0.714 | 0.673 | +0.086 (-0.056, +0.233) | 0.423 |
| Glaucoma | 55 | 0.625 | 0.650 | 0.684 | +0.059 (-0.019, +0.137) | 0.350 |
| Atrial fibrillation (persistent) | 14 | 0.748 | 0.708 | 0.807 | +0.059 (-0.045, +0.164) | 0.432 |
| Atrial fibrillation (paroxysmal) | 74 | 0.675 | 0.646 | 0.715 | +0.040 (+0.003, +0.074) | 0.098 |
| **Atrial fibrillation** | 163 | 0.743 | 0.641 | 0.779 | +0.036 (+0.011, +0.060) | 0.026 |
| Psoriasis | 29 | 0.653 | 0.629 | 0.688 | +0.035 (-0.051, +0.128) | 0.612 |
| Coronary atherosclerosis | 70 | 0.769 | 0.664 | 0.801 | +0.032 (+0.008, +0.056) | 0.062 |
| Acute myocardial infarction | 48 | 0.746 | 0.648 | 0.775 | +0.028 (-0.015, +0.068) | 0.416 |
| Asthma | 160 | 0.588 | 0.592 | 0.615 | +0.027 (-0.009, +0.063) | 0.333 |
| Type 2 diabetes | 128 | 0.780 | 0.634 | 0.802 | +0.022 (+0.002, +0.040) | 0.097 |
| Myocardial infarction | 85 | 0.762 | 0.638 | 0.783 | +0.021 (+0.002, +0.043) | 0.098 |
| Pulmonary embolism | 32 | 0.682 | 0.617 | 0.696 | +0.014 (-0.063, +0.088) | 0.841 |
| Ulcerative colitis | 14 | 0.564 | 0.676 | 0.578 | +0.014 (-0.178, +0.167) | 0.879 |
| Essential hypertension | 541 | 0.717 | 0.594 | 0.730 | +0.014 (+0.004, +0.025) | 0.052 |
| Ischaemic heart disease | 105 | 0.771 | 0.603 | 0.782 | +0.010 (-0.007, +0.027) | 0.423 |
| Melanoma/skin cancer | 144 | 0.650 | 0.559 | 0.653 | +0.002 (-0.019, +0.023) | 0.897 |
| Rheumatoid arthritis | 25 | 0.726 | 0.529 | 0.728 | +0.002 (-0.047, +0.057) | 0.942 |
| Macular degeneration | 68 | 0.777 | 0.621 | 0.777 | +0.000 (-0.035, +0.032) | 0.986 |
| Parkinson's disease | 23 | 0.914 | 0.544 | 0.913 | -0.001 (-0.006, +0.003) | 0.705 |
| Dementia | 23 | 0.788 | 0.526 | 0.784 | -0.004 (-0.022, +0.014) | 0.758 |
| Ischaemic stroke | 27 | 0.791 | 0.394 | 0.779 | -0.012 (-0.024, -0.002) | 0.062 |
| Alzheimer's disease | 11 | 0.835 | 0.620 | 0.819 | -0.016 (-0.072, +0.041) | 0.705 |

**Supplementary Table 5.** Disease-matched PRS to phecode crosswalk. Each polygenic risk score (UK Biobank Standard release field) matched by hand to the corresponding incident phecode(s) in the panel. Cancer scores without a specific malignant phecode in the panel were excluded (melanoma, matched to CA_1<u>03, was retained).</u>

| PRS trait | Standard / Enhanced field | Matched phecode(s) |
| --- | --- | --- |
| alzheimers | 26206 / 26207 | NS_328.11, NS_328.1 |
| asthma | 26210 / 26211 | RE_475 |
| atrial fibrillation | 26212 / 26213 | CV_416.2, CV_416.211, CV_416.212 |
| coeliac disease | 26225 / 26226 | GI_525.1 |
| coronary artery disease | 26227 / 26228 | CV_404, CV_404.1, CV_404.11, CV_404.2 |
| crohns disease | 26229 / - | GI_522.11 |
| glaucoma | 26265 / 26266 | SO_375.1, SO_375.11 |
| hypertension | 26244 / 26245 | CV_401.1, CV_402 |
| ischaemic stroke | 26248 / 26249 | CV_431.11 |
| macular degeneration | 26204 / 26205 | SO_374.5 |
| melanoma | 26252 / 26253 | CA_103 |
| multiple sclerosis | 26254 / 26255 | NS_326.1 |
| parkinsons | 26260 / 26261 | NS_324.11 |
| psoriasis | 26269 / 26270 | DE_664.4 |
| rheumatoid arthritis | 26273 / 26274 | MS_705.1 |
| type1 diabetes | 26283 / 26284 | EM_202.1 |
| type2 diabetes | 26285 / 26286 | EM_202.2 |
| ulcerative colitis | 26287 / - | GI_522.12 |
| venous thromboembolism | 26289 / 26290 | CV_440.3 |

**Supplementary Table 6.** Per-disease concordance for evaluable outcomes with at least five incident test cases. , ordered by concordance, with 95% subject-level bootstrap confidence intervals. Events are incident cases in the held-out test set; estimates for outcomes with few events are unstable and carry wide intervals. Outcomes with fewer than five incident test cases are not shown, in line with UK Biobank’s small-numbers policy.

| Disease | Category | Events | C | 95% CI |
| --- | --- | --- | --- | --- |
| Parkinson’s disease (Primary) | Neurological | 25 | 0.909 | [0.816, 0.966] |
| Chronic prostatitis | Genitourinary | 7 | 0.882 | [0.835, 0.921] |
| Memory loss | Neurological | 8 | 0.876 | [0.759, 0.966] |
| Hypoglycaemia | Endocrine/Metab | 9 | 0.873 | [0.784, 0.967] |
| Disorders of penis | Genitourinary | 19 | 0.864 | [0.789, 0.926] |
| Benign prostatic hyperplasia | Genitourinary | 168 | 0.863 | [0.846, 0.878] |
| Heart failure | Cardiovascular | 37 | 0.862 | [0.818, 0.914] |
| Atrioventricular block, first degree | Cardiovascular | 36 | 0.857 | [0.799, 0.903] |
| Hyperkalaemia | Endocrine/Metab | 21 | 0.850 | [0.771, 0.927] |
| Psychoactive substance dependence | Mental | 7 | 0.849 | [0.778, 0.927] |
| Alzheimer’s disease | Neurological | 15 | 0.846 | [0.711, 0.946] |
| Disorders of prostate | Genitourinary | 9 | 0.843 | [0.769, 0.895] |
| Other diseases of biliary tract | Gastrointestinal | 15 | 0.837 | [0.770, 0.908] |
| Pressure ulcer | Dermatological | 29 | 0.836 | [0.778, 0.890] |
| Hydrocele | Genitourinary | 14 | 0.836 | [0.768, 0.898] |
| Aortic valve disorders | Cardiovascular | 18 | 0.833 | [0.751, 0.903] |
| Non-pressure chronic ulcer | Dermatological | 16 | 0.831 | [0.720, 0.924] |
| Repeated falls* | Neurological | 97 | 0.827 | [0.785, 0.870] |
| Aortic stenosis | Cardiovascular | 39 | 0.827 | [0.769, 0.880] |
| Atrioventricular block, complete | Cardiovascular | 13 | 0.826 | [0.725, 0.923] |
| Acidosis | Endocrine/Metab | 25 | 0.825 | [0.756, 0.890] |
| Occlusion and stenosis of cerebral arteries | Cardiovascular | 12 | 0.824 | [0.733, 0.912] |
| Hemiplegia and hemiparesis | Neurological | 23 | 0.822 | [0.747, 0.881] |
| Epilepsy | Neurological | 15 | 0.819 | [0.725, 0.904] |
| Obesity | Endocrine/Metab | 222 | 0.819 | [0.787, 0.843] |
| Diverticulum of bladder | Genitourinary | 11 | 0.812 | [0.616, 0.925] |
| Fluid overload | Endocrine/Metab | 12 | 0.811 | [0.698, 0.908] |
| Diseases of the pancreas | Gastrointestinal | 12 | 0.811 | [0.698, 0.913] |
| Chronic obstructive pulmonary disease | Respiratory | 92 | 0.808 | [0.763, 0.849] |
| Uterine/Uterovaginal prolapse | Genitourinary | 50 | 0.808 | [0.772, 0.837] |
| Sequelae of cancer | Neoplasms | 27 | 0.806 | [0.738, 0.858] |
| Bladder neck obstruction | Genitourinary | 10 | 0.806 | [0.717, 0.881] |
| End stage renal disease | Genitourinary | 11 | 0.805 | [0.624, 0.964] |
| Pneumonia | Respiratory | 60 | 0.805 | [0.746, 0.855] |
| Sleep apnoea | Neurological | 57 | 0.805 | [0.739, 0.863] |
| Inguinal hernia | Gastrointestinal | 128 | 0.803 | [0.776, 0.829] |
| Acute kidney failure | Genitourinary | 136 | 0.802 | [0.768, 0.829] |
| Thyrotoxicosis | Endocrine/Metab | 10 | 0.801 | [0.675, 0.906] |
| Dementias | Neurological | 27 | 0.801 | [0.693, 0.884] |
| Left heart failure | Cardiovascular | 52 | 0.801 | [0.744, 0.853] |
| Cerebral degeneration | Neurological | 16 | 0.800 | [0.702, 0.892] |
| Aspiration pneumonia | Respiratory | 20 | 0.800 | [0.699, 0.876] |
| Arterial embolism and thrombosis | Cardiovascular | 14 | 0.798 | [0.700, 0.873] |
| Pulmonary hypertension | Cardiovascular | 14 | 0.798 | [0.658, 0.906] |
| Atrophic vaginitis | Genitourinary | 13 | 0.797 | [0.705, 0.898] |
| Abdominal aortic aneurysm | Cardiovascular | 18 | 0.797 | [0.685, 0.891] |
| Cerebral infarction | Cardiovascular | 33 | 0.793 | [0.717, 0.852] |
| Pain in hip | Musculoskeletal | 27 | 0.789 | [0.710, 0.867] |
| Ischaemic heart disease | Cardiovascular | 128 | 0.787 | [0.749, 0.819] |
| Lump or mass in breast | Neoplasms | 5 | 0.785 | [0.595, 0.960] |
| Symptoms and signs involving cognitive function | Neurological | 19 | 0.785 | [0.703, 0.866] |
| Epiphora | Sense organs | 6 | 0.785 | [0.564, 0.958] |
| Presence of cardiac device | Cardiovascular | 41 | 0.785 | [0.720, 0.849] |
| Other arthropathies | Musculoskeletal | 6 | 0.784 | [0.546, 0.990] |
| Type 2 diabetes | Endocrine/Metab | 159 | 0.777 | [0.743, 0.813] |
| Ventricular premature depolarisation* | Cardiovascular | 15 | 0.777 | [0.705, 0.843] |
| Macular degeneration | Sense organs | 77 | 0.777 | [0.728, 0.821] |
| Pleurisy | Respiratory | 17 | 0.775 | [0.656, 0.876] |
| Vitamin B12 deficiency | Endocrine/Metab | 7 | 0.775 | [0.575, 0.931] |
| Coronary atherosclerosis | Cardiovascular | 86 | 0.774 | [0.729, 0.806] |
| Chronic kidney disease | Genitourinary | 151 | 0.772 | [0.728, 0.806] |
| Aortic insufficiency | Cardiovascular | 24 | 0.772 | [0.699, 0.836] |
| Urinary incontinence and enuresis | Genitourinary | 54 | 0.769 | [0.697, 0.833] |
| Orthostatic hypotension | Cardiovascular | 50 | 0.769 | [0.710, 0.825] |
| Complications and ill-defined descriptions of | Cardiovascular | 72 | 0.768 | [0.719, 0.808] |
| Entropion and trichiasis of eyelid | Sense organs | 6 | 0.767 | [0.618, 0.899] |
| All-cause mortality | Mortality | 213 | 0.767 | [0.729, 0.800] |
| Obstruction of bile duct | Gastrointestinal | 14 | 0.767 | [0.681, 0.851] |
| Disorders of magnesium metabolism | Endocrine/Metab | 18 | 0.767 | [0.675, 0.850] |
| Urethral stricture | Genitourinary | 22 | 0.766 | [0.673, 0.847] |
| Epidermal thickening | Dermatological | 12 | 0.764 | [0.631, 0.880] |
| Testicular limpoma | Neoplasms | 11 | 0.764 | [0.702, 0.834] |
| Retention of urine | Genitourinary | 129 | 0.764 | [0.722, 0.802] |
| Altered mental status, unspecified | Neurological | 41 | 0.764 | [0.695, 0.827] |
| Myocardial infarction | Cardiovascular | 104 | 0.763 | [0.723, 0.805] |
| Other mental and behavioural problems | Mental | 18 | 0.763 | [0.611, 0.876] |
| Leiomyoma of uterus | Neoplasms | 31 | 0.763 | [0.712, 0.818] |
| Haematemesis | Gastrointestinal | 10 | 0.762 | [0.636, 0.904] |
| Mononeuritis of upper limb | Neurological | 9 | 0.761 | [0.599, 0.898] |
| Abnormality of gait and mobility | Neurological | 60 | 0.760 | [0.691, 0.822] |
| Interstitial pulmonary diseases | Respiratory | 26 | 0.759 | [0.668, 0.845] |
| Diplopia | Sense organs | 17 | 0.759 | [0.628, 0.874] |
| Pneumothorax and air leak | Respiratory | 13 | 0.758 | [0.589, 0.885] |
| Bradycardia | Cardiovascular | 53 | 0.758 | [0.705, 0.811] |
| Tricuspid valve disorders | Cardiovascular | 53 | 0.758 | [0.701, 0.814] |
| Cholangitis | Gastrointestinal | 15 | 0.755 | [0.673, 0.835] |
| Hammer toe | Musculoskeletal | 13 | 0.754 | [0.645, 0.865] |
| Scoliosis | Musculoskeletal | 16 | 0.753 | [0.623, 0.865] |
| Peripheral vascular disease NOS | Cardiovascular | 26 | 0.752 | [0.625, 0.843] |
| Other disorders of male genital organs | Genitourinary | 27 | 0.751 | [0.704, 0.799] |
| Acute myocardial infarction | Cardiovascular | 60 | 0.750 | [0.692, 0.806] |
| Persistent atrial fibrillation* | Cardiovascular | 18 | 0.750 | [0.649, 0.836] |
| Fatty liver disease (FLD) | Gastrointestinal | 42 | 0.750 | [0.680, 0.808] |
| Gout | Musculoskeletal | 53 | 0.750 | [0.702, 0.799] |
| Pain in shoulder | Musculoskeletal | 20 | 0.750 | [0.686, 0.821] |
| Hypokalaemia | Endocrine/Metab | 47 | 0.750 | [0.681, 0.810] |
| Disorders of the retina | Sense organs | 25 | 0.750 | [0.661, 0.831] |
| Volume depletion | Endocrine/Metab | 63 | 0.749 | [0.696, 0.808] |
| Stress incontinence | Genitourinary | 15 | 0.748 | [0.666, 0.844] |
| Atrial fibrillation and flutter | Cardiovascular | 183 | 0.748 | [0.715, 0.778] |
| Nuclear cataract | Sense organs | 168 | 0.747 | [0.713, 0.776] |
| Cystitis | Genitourinary | 11 | 0.747 | [0.647, 0.843] |
| Other cerebrovascular disease | Cardiovascular | 54 | 0.746 | [0.694, 0.792] |
| Phlebitis and thrombophlebitis | Cardiovascular | 33 | 0.746 | [0.663, 0.814] |
| Emphysema | Respiratory | 16 | 0.744 | [0.591, 0.881] |
| Gastrointestinal angiodysplasia | Gastrointestinal | 19 | 0.744 | [0.656, 0.824] |
| Cataract | Sense organs | 383 | 0.743 | [0.720, 0.765] |
| Cardiac arrhythmia and conduction disorders | Cardiovascular | 24 | 0.743 | [0.647, 0.824] |
| Epistaxis | Respiratory | 18 | 0.743 | [0.627, 0.868] |
| Blepharitis | Sense organs | 8 | 0.742 | [0.566, 0.887] |
| Left bundle branch block | Cardiovascular | 34 | 0.742 | [0.665, 0.802] |
| Ventral hernia | Gastrointestinal | 13 | 0.741 | [0.587, 0.867] |
| Acute lower respiratory infection | Respiratory | 64 | 0.738 | [0.671, 0.801] |
| Phobic disorders | Mental | 16 | 0.738 | [0.648, 0.830] |
| Sensorineural hearing loss | Sense organs | 30 | 0.737 | [0.673, 0.803] |
| Fracture of femur | Musculoskeletal | 34 | 0.737 | [0.657, 0.811] |
| Disorders of uterus, except cervix | Genitourinary | 16 | 0.737 | [0.642, 0.832] |
| Open angle glaucoma | Sense organs | 19 | 0.736 | [0.639, 0.829] |
| Haemorrhagic stroke | Cardiovascular | 14 | 0.735 | [0.628, 0.833] |
| Hyperlipidaemia | Endocrine/Metab | 70 | 0.734 | [0.681, 0.784] |
| Effusion of joint | Musculoskeletal | 17 | 0.734 | [0.606, 0.854] |
| Corneal degenerations | Sense organs | 29 | 0.733 | [0.663, 0.804] |
| Benign neoplasms of the ovary | Neoplasms | 10 | 0.732 | [0.606, 0.851] |
| Atherosclerosis | Cardiovascular | 145 | 0.732 | [0.698, 0.768] |
| Viral pneumonia | Respiratory | 24 | 0.731 | [0.593, 0.850] |
| Cardiomegaly | Cardiovascular | 68 | 0.731 | [0.679, 0.778] |
| Vitamin B group deficiency | Endocrine/Metab | 26 | 0.730 | [0.647, 0.823] |
| Spondylosis | Musculoskeletal | 37 | 0.729 | [0.644, 0.795] |
| Abnormal findings on diagnostic imaging of lun | Respiratory | 38 | 0.729 | [0.655, 0.804] |
| Hyposmolality and/or hyponatraemia | Endocrine/Metab | 62 | 0.728 | [0.662, 0.784] |
| Respiratory failure | Respiratory | 32 | 0.726 | [0.640, 0.800] |
| Other disorders of bone | Musculoskeletal | 50 | 0.724 | [0.660, 0.777] |
| Rheumatoid arthritis | Musculoskeletal | 31 | 0.723 | [0.597, 0.820] |
| Carcinoma in situ of skin | Neoplasms | 17 | 0.722 | [0.570, 0.852] |
| Ascites | Gastrointestinal | 22 | 0.720 | [0.618, 0.828] |
| Myalgia | Musculoskeletal | 35 | 0.720 | [0.627, 0.807] |
| Essential hypertension | Cardiovascular | 646 | 0.719 | [0.700, 0.736] |
| Pathological fracture | Musculoskeletal | 17 | 0.717 | [0.527, 0.845] |
| Intestinal perforation | Gastrointestinal | 13 | 0.717 | [0.614, 0.807] |
| Cardiac murmurs | Cardiovascular | 22 | 0.716 | [0.591, 0.799] |
| Osteoarthritis of more than one site | Musculoskeletal | 70 | 0.716 | [0.660, 0.766] |
| Urinary tract infection | Genitourinary | 113 | 0.715 | [0.667, 0.762] |
| Vitreous opacities | Sense organs | 21 | 0.713 | [0.600, 0.809] |
| Pericardial effusion (noninflammatory)* | Cardiovascular | 9 | 0.713 | [0.542, 0.854] |
| Nonspecific low blood-pressure reading | Cardiovascular | 22 | 0.712 | [0.642, 0.787] |
| Cyst of kidney | Genitourinary | 36 | 0.712 | [0.638, 0.782] |
| Carotid artery stenosis | Cardiovascular | 9 | 0.711 | [0.543, 0.888] |
| Mucous polyp of cervix | Genitourinary | 10 | 0.711 | [0.604, 0.813] |
| Pure hypercholesterolaemia | Endocrine/Metab | 337 | 0.710 | [0.685, 0.732] |
| Disorders of lacrimal system | Sense organs | 9 | 0.710 | [0.519, 0.832] |
| Spinal stenosis | Musculoskeletal | 56 | 0.710 | [0.636, 0.776] |
| Umbilical hernia | Gastrointestinal | 27 | 0.710 | [0.613, 0.802] |
| Hallux valgus (Bunion) | Musculoskeletal | 28 | 0.710 | [0.635, 0.780] |
| Polyp of corpus uteri | Genitourinary | 39 | 0.710 | [0.659, 0.764] |
| Anorexia | Endocrine/Metab | 15 | 0.708 | [0.578, 0.803] |
| Fracture of upper limb | Musculoskeletal | 55 | 0.708 | [0.640, 0.773] |
| Pulmonary embolism | Cardiovascular | 38 | 0.708 | [0.622, 0.784] |
| Hypotension | Cardiovascular | 84 | 0.707 | [0.650, 0.762] |
| Lobar pneumonia* | Respiratory | 103 | 0.705 | [0.650, 0.753] |
| Amblyopia | Sense organs | 14 | 0.705 | [0.552, 0.848] |
| Back pain | Musculoskeletal | 63 | 0.705 | [0.636, 0.777] |
| Transient cerebral ischaemic attack | Cardiovascular | 13 | 0.705 | [0.540, 0.848] |
| Carpal tunnel syndrome | Neurological | 50 | 0.704 | [0.650, 0.771] |
| Fracture of vertebrae | Musculoskeletal | 16 | 0.704 | [0.584, 0.804] |
| Raynaud's syndrome | Cardiovascular | 20 | 0.704 | [0.580, 0.852] |
| Sepsis | Infections | 101 | 0.704 | [0.645, 0.753] |
| Polyneuropathies | Neurological | 28 | 0.704 | [0.606, 0.824] |
| Aphasia and dysphasia | Neurological | 15 | 0.702 | [0.576, 0.814] |
| Vitamin D deficiency | Endocrine/Metab | 38 | 0.702 | [0.610, 0.792] |
| Bacterial infections | Infections | 94 | 0.702 | [0.639, 0.758] |
| Astigmatism | Sense organs | 76 | 0.701 | [0.648, 0.758] |
| Polymyalgia rheumatica | Musculoskeletal | 31 | 0.701 | [0.600, 0.785] |
| Constipation | Gastrointestinal | 194 | 0.700 | [0.665, 0.730] |
| Multiple sclerosis | Neurological | 7 | 0.700 | [0.374, 0.955] |
| Infection of the skin | Dermatological | 12 | 0.700 | [0.494, 0.883] |
| Pulmonary collapse | Respiratory | 55 | 0.699 | [0.646, 0.762] |
| Abnormal findings on diagnostic imaging of the | Gastrointestinal | 37 | 0.699 | [0.623, 0.776] |
| Faecal incontinence and encopresis | Gastrointestinal | 31 | 0.699 | [0.597, 0.799] |
| Functional dyspepsia | Gastrointestinal | 45 | 0.699 | [0.640, 0.773] |
| Heart valve replaced | Cardiovascular | 19 | 0.697 | [0.561, 0.844] |
| Postmenopausal bleeding | Genitourinary | 29 | 0.696 | [0.635, 0.756] |
| Palmar fascial fibromatosis | Musculoskeletal | 36 | 0.696 | [0.620, 0.776] |
| Paroxysmal atrial fibrillation* | Cardiovascular | 83 | 0.695 | [0.644, 0.753] |
| Osteoarthritis | Musculoskeletal | 424 | 0.695 | [0.669, 0.717] |
| Angle-Closure Glaucoma | Sense organs | 18 | 0.694 | [0.568, 0.796] |
| Hydronephrosis | Genitourinary | 38 | 0.694 | [0.610, 0.785] |
| Osteophyte* | Musculoskeletal | 34 | 0.694 | [0.624, 0.774] |
| Actinic keratosis | Dermatological | 51 | 0.694 | [0.631, 0.757] |
| Hearing impairment | Sense organs | 106 | 0.694 | [0.649, 0.738] |
| Bursitis | Musculoskeletal | 24 | 0.692 | [0.598, 0.774] |
| Dizziness and giddiness | Neurological | 76 | 0.691 | [0.640, 0.741] |
| Cellulitis and abscess | Dermatological | 98 | 0.691 | [0.633, 0.738] |
| Intestinal fistula | Gastrointestinal | 8 | 0.691 | [0.525, 0.846] |
| Blood in stool | Gastrointestinal | 31 | 0.691 | [0.602, 0.770] |
| Other diseases of stomach and duodenum | Gastrointestinal | 26 | 0.690 | [0.605, 0.773] |
| Gallstones | Gastrointestinal | 97 | 0.690 | [0.637, 0.741] |
| Disorders of calcium metabolism | Endocrine/Metab | 27 | 0.690 | [0.575, 0.783] |
| Inflammatory spondylopathies | Musculoskeletal | 20 | 0.689 | [0.552, 0.783] |
| Renal failure | Genitourinary | 12 | 0.688 | [0.551, 0.810] |
| Pleural effusion | Respiratory | 94 | 0.688 | [0.636, 0.738] |
| Myelopathies | Neurological | 12 | 0.688 | [0.511, 0.864] |
| Low back pain | Musculoskeletal | 43 | 0.688 | [0.608, 0.757] |
| Bronchiectasis | Respiratory | 38 | 0.688 | [0.609, 0.762] |
| Anxiety and anxiety disorders | Mental | 126 | 0.687 | [0.642, 0.737] |
| Right bundle branch block | Cardiovascular | 37 | 0.687 | [0.602, 0.786] |
| Other respiratory disorders | Respiratory | 32 | 0.686 | [0.593, 0.774] |
| Major depressive disorder | Mental | 159 | 0.685 | [0.647, 0.729] |
| Ventricular tachycardia | Cardiovascular | 8 | 0.684 | [0.490, 0.859] |
| Pelvic peritoneal adhesions, female (postopera | Genitourinary | 5 | 0.683 | [0.498, 0.908] |
| Rotator cuff tear or rupture | Musculoskeletal | 26 | 0.683 | [0.584, 0.776] |
| Viral infections | Infections | 87 | 0.682 | [0.616, 0.735] |
| Anaemia | Blood/Immune | 161 | 0.681 | [0.638, 0.720] |
| Abnormal glucose | Endocrine/Metab | 24 | 0.681 | [0.578, 0.766] |
| Hypothyroidism (not specified as secondary) | Endocrine/Metab | 164 | 0.680 | [0.643, 0.723] |
| Trigger finger | Musculoskeletal | 20 | 0.679 | [0.552, 0.797] |
| Fracture of bones in thorax | Musculoskeletal | 16 | 0.679 | [0.535, 0.786] |
| Hallux rigidus | Musculoskeletal | 8 | 0.678 | [0.534, 0.820] |
| Tachycardia | Cardiovascular | 34 | 0.677 | [0.584, 0.769] |
| Spondylolisthesis | Musculoskeletal | 23 | 0.676 | [0.569, 0.757] |
| Diverticular disease | Gastrointestinal | 106 | 0.673 | [0.623, 0.725] |
| Angina pectoris | Cardiovascular | 105 | 0.673 | [0.626, 0.715] |
| Tricuspid valve insufficiency* | Cardiovascular | 14 | 0.672 | [0.556, 0.789] |
| Nonspecific abnormal results of function study | Gastrointestinal | 56 | 0.672 | [0.612, 0.724] |
| Other disorders of bladder | Genitourinary | 81 | 0.672 | [0.605, 0.735] |
| Dyspnoea | Respiratory | 81 | 0.670 | [0.616, 0.724] |
| Enthesopathy/Enthesitis/Tendinopathy | Musculoskeletal | 13 | 0.664 | [0.491, 0.820] |
| Diseases of vocal cords and larynx, not elsewh | Respiratory | 9 | 0.664 | [0.483, 0.842] |
| Pain in knee | Musculoskeletal | 14 | 0.663 | [0.525, 0.792] |
| Gastric ulcer | Gastrointestinal | 43 | 0.662 | [0.590, 0.733] |
| Noninflammatory disorders of vulva and perineu | Genitourinary | 6 | 0.662 | [0.502, 0.793] |
| Peritonitis | Gastrointestinal | 16 | 0.661 | [0.547, 0.783] |
| Malignant neoplasm of the skin | Neoplasms | 168 | 0.661 | [0.624, 0.703] |
| Cervicalgia | Musculoskeletal | 11 | 0.660 | [0.505, 0.822] |
| Anal fissure | Gastrointestinal | 8 | 0.660 | [0.393, 0.937] |
| Other symptoms/disorders or the urinary system | Genitourinary | 22 | 0.659 | [0.539, 0.759] |
| Thrombocytopenia | Blood/Immune | 24 | 0.657 | [0.546, 0.769] |
| Diaphragmatic hernia | Gastrointestinal | 253 | 0.657 | [0.624, 0.694] |
| Cardiomyopathy | Cardiovascular | 8 | 0.657 | [0.432, 0.844] |
| Cancer (solid tumour, excluding BCC) | Neoplasms | 515 | 0.656 | [0.635, 0.677] |
| Haematuria | Genitourinary | 77 | 0.655 | [0.591, 0.722] |
| Enlargement of lymph nodes | Blood/Immune | 35 | 0.655 | [0.578, 0.729] |
| Mitral valve insufficiency | Cardiovascular | 50 | 0.654 | [0.576, 0.723] |
| Sebaceous cyst | Dermatological | 7 | 0.654 | [0.381, 0.869] |
| Dermatitis | Dermatological | 40 | 0.653 | [0.563, 0.733] |
| Chronic cholecystitis | Gastrointestinal | 9 | 0.653 | [0.459, 0.821] |
| Pain in joint | Musculoskeletal | 17 | 0.652 | [0.503, 0.777] |
| Supraventricular tachycardia | Cardiovascular | 21 | 0.652 | [0.545, 0.757] |
| Secondary malignant neoplasm | Neoplasms | 140 | 0.650 | [0.603, 0.698] |
| Hyperparathyroidism | Endocrine/Metab | 11 | 0.650 | [0.469, 0.833] |
| Disorders of phosphorus metabolism | Endocrine/Metab | 22 | 0.650 | [0.565, 0.736] |
| Frequency of urination and polyuria | Genitourinary | 16 | 0.649 | [0.470, 0.800] |
| Disorder of patella | Musculoskeletal | 12 | 0.646 | [0.506, 0.753] |
| Other disorders of the intestines | Gastrointestinal | 48 | 0.646 | [0.575, 0.723] |
| Other skin and subcutaneous disorders | Dermatological | 63 | 0.646 | [0.590, 0.697] |
| Disturbances of skin sensation | Neurological | 17 | 0.646 | [0.475, 0.781] |
| Fungal infections | Infections | 42 | 0.645 | [0.576, 0.712] |
| Spinal disc displacement (herniation) | Musculoskeletal | 52 | 0.645 | [0.578, 0.725] |
| Other disorders of liver | Gastrointestinal | 33 | 0.645 | [0.532, 0.741] |
| Mononeuritis of lower limb | Neurological | 11 | 0.644 | [0.479, 0.797] |
| Cough | Respiratory | 40 | 0.644 | [0.548, 0.737] |
| Impingement syndrome of shoulder* | Musculoskeletal | 37 | 0.644 | [0.560, 0.728] |
| Varicose veins of lower extremities | Cardiovascular | 39 | 0.642 | [0.559, 0.732] |
| Periodontitis | Gastrointestinal | 7 | 0.641 | [0.424, 0.838] |
| Duodenitis | Gastrointestinal | 54 | 0.640 | [0.577, 0.703] |
| Irritable bowel syndrome | Gastrointestinal | 74 | 0.639 | [0.575, 0.699] |
| Intestinal or peritoneal adhesions | Gastrointestinal | 73 | 0.639 | [0.577, 0.699] |
| Glaucoma | Sense organs | 66 | 0.637 | [0.575, 0.695] |
| Migraine | Neurological | 35 | 0.637 | [0.546, 0.719] |
| Abnormal weight loss | Endocrine/Metab | 109 | 0.637 | [0.580, 0.688] |
| Barrett's oesophagus | Gastrointestinal | 41 | 0.636 | [0.541, 0.722] |
| Iron deficiency | Endocrine/Metab | 154 | 0.636 | [0.595, 0.677] |
| Rectal prolapse | Gastrointestinal | 14 | 0.636 | [0.485, 0.812] |
| Ptois of eyelid | Sense organs | 18 | 0.636 | [0.505, 0.770] |
| Diverticula of colon | Gastrointestinal | 388 | 0.636 | [0.607, 0.661] |
| Loose body in joint | Musculoskeletal | 6 | 0.635 | [0.447, 0.835] |
| Secondary hypothyroidism | Endocrine/Metab | 19 | 0.634 | [0.499, 0.765] |
| Intervertebral disc disorder | Musculoskeletal | 40 | 0.629 | [0.532, 0.715] |
| Flatulence, eructation, and gas pain | Gastrointestinal | 20 | 0.627 | [0.509, 0.743] |
| Benign neoplasm of stomach | Neoplasms | 108 | 0.626 | [0.574, 0.675] |
| Oesophageal obstruction (Stricture and stenosis) | Gastrointestinal | 23 | 0.624 | [0.530, 0.722] |
| Nasal polyps | Respiratory | 11 | 0.624 | [0.444, 0.757] |
| Psoriasis | Dermatological | 31 | 0.624 | [0.509, 0.717] |
| Kidney stone disease | Genitourinary | 37 | 0.623 | [0.520, 0.703] |
| Fracture of lower limb | Musculoskeletal | 16 | 0.622 | [0.469, 0.748] |
| Diseases of oesophagus | Gastrointestinal | 29 | 0.622 | [0.527, 0.735] |
| Rash and other nonspecific skin eruption | Dermatological | 23 | 0.621 | [0.494, 0.751] |
| Myopia | Sense organs | 38 | 0.621 | [0.535, 0.689] |
| Aphagia and dysphagia | Gastrointestinal | 104 | 0.620 | [0.563, 0.672] |
| Gastro-oesophageal reflux disease | Gastrointestinal | 334 | 0.618 | [0.587, 0.647] |
| Acquired deformities of toe | Musculoskeletal | 11 | 0.618 | [0.461, 0.774] |
| Lesions of mouth | Gastrointestinal | 12 | 0.615 | [0.439, 0.782] |
| Benign neoplasm of the skin | Neoplasms | 14 | 0.615 | [0.486, 0.720] |
| Corpus luteum cyst or haematoma | Genitourinary | 12 | 0.614 | [0.556, 0.676] |
| Gastrointestinal haemorrhage | Gastrointestinal | 110 | 0.614 | [0.567, 0.668] |
| Ganglion cyst | Musculoskeletal | 13 | 0.614 | [0.453, 0.770] |
| Acute pancreatitis | Gastrointestinal | 24 | 0.613 | [0.467, 0.728] |
| Oesophagitis | Gastrointestinal | 66 | 0.611 | [0.545, 0.672] |
| Localised swelling, mass and lump of skin and | Dermatological | 13 | 0.611 | [0.457, 0.744] |
| Haemo onc - by cell of origin | Neoplasms | 18 | 0.610 | [0.471, 0.792] |
| Renal tubulo-Interstitial diseases | Genitourinary | 13 | 0.610 | [0.413, 0.791] |
| Ulcerative colitis | Gastrointestinal | 18 | 0.610 | [0.489, 0.716] |
| Dental caries | Gastrointestinal | 22 | 0.607 | [0.497, 0.733] |
| Chronic cystitis | Genitourinary | 8 | 0.606 | [0.413, 0.799] |
| Duodenal ulcer | Gastrointestinal | 15 | 0.605 | [0.444, 0.760] |
| Seborrheic keratosis | Dermatological | 39 | 0.604 | [0.534, 0.670] |
| Abdominal pain | Gastrointestinal | 163 | 0.602 | [0.556, 0.644] |
| Radiculopathy | Musculoskeletal | 16 | 0.601 | [0.434, 0.747] |
| Tinnitus | Sense organs | 9 | 0.600 | [0.438, 0.777] |
| Psychoactive substance abuse | Mental | 20 | 0.599 | [0.484, 0.704] |
| Benign neoplasm of the colon | Neoplasms | 295 | 0.599 | [0.568, 0.628] |
| Paresthesia of skin* | Neurological | 14 | 0.598 | [0.449, 0.757] |
| Melanocytic nevi* | Neoplasms | 30 | 0.596 | [0.499, 0.704] |
| Intestinal obstruction | Gastrointestinal | 22 | 0.596 | [0.479, 0.717] |
| Other disorders of the kidney and ureters | Genitourinary | 17 | 0.596 | [0.448, 0.725] |
| Neutropenia | Blood/Immune | 39 | 0.595 | [0.510, 0.667] |
| Oesophageal ulcer | Gastrointestinal | 15 | 0.595 | [0.473, 0.713] |
| Non-Hodgkin lymphoma | Neoplasms | 16 | 0.595 | [0.417, 0.766] |
| Abnormalities of breathing | Respiratory | 20 | 0.594 | [0.468, 0.734] |
| Fracture of hand or wrist | Musculoskeletal | 16 | 0.594 | [0.409, 0.779] |
| Symptoms involving digestive system | Gastrointestinal | 138 | 0.593 | [0.548, 0.637] |
| Asthma | Respiratory | 188 | 0.592 | [0.544, 0.635] |
| Bacterial enteritis | Gastrointestinal | 15 | 0.592 | [0.410, 0.742] |
| Other disorders of the eyelids | Sense organs | 20 | 0.592 | [0.489, 0.721] |
| Retinal detachments and breaks | Sense organs | 27 | 0.591 | [0.489, 0.683] |
| Haemoptysis | Respiratory | 15 | 0.590 | [0.435, 0.726] |
| Benign neoplasm of rectum and anus | Neoplasms | 120 | 0.589 | [0.535, 0.635] |
| Haemorrhoids | Cardiovascular | 256 | 0.587 | [0.555, 0.620] |
| Gastrointestinal inflammation | Gastrointestinal | 74 | 0.584 | [0.513, 0.642] |
| Vomiting | Gastrointestinal | 122 | 0.583 | [0.529, 0.631] |
| Fracture of pelvis | Musculoskeletal | 14 | 0.583 | [0.376, 0.761] |
| Chalazion | Sense organs | 6 | 0.582 | [0.420, 0.773] |
| Palpitations | Cardiovascular | 39 | 0.582 | [0.491, 0.671] |
| Gastritis | Gastrointestinal | 225 | 0.581 | [0.542, 0.620] |
| Acute Pharyngitis | Respiratory | 13 | 0.578 | [0.414, 0.745] |
| Crohn's disease | Gastrointestinal | 17 | 0.572 | [0.435, 0.689] |
| Disorders of the gallbladder | Gastrointestinal | 12 | 0.571 | [0.454, 0.703] |
| Sciatica | Musculoskeletal | 23 | 0.570 | [0.440, 0.707] |
| Other follicular disorders | Dermatological | 10 | 0.568 | [0.415, 0.701] |
| Impacted teeth* | Gastrointestinal | 5 | 0.564 | [0.367, 0.804] |
| Abnormal intraocular pressure | Sense organs | 31 | 0.563 | [0.474, 0.626] |
| Deviated nasal septum | Respiratory | 12 | 0.561 | [0.382, 0.689] |
| Symptoms related to joints | Musculoskeletal | 5 | 0.554 | [0.361, 0.703] |
| Headache | Neurological | 61 | 0.554 | [0.487, 0.628] |
| Disease of anus and rectum | Gastrointestinal | 42 | 0.554 | [0.484, 0.634] |
| Scar conditions and fibrosis of skin | Dermatological | 10 | 0.551 | [0.336, 0.733] |
| Derangement of meniscus | Musculoskeletal | 42 | 0.550 | [0.447, 0.642] |
| Coeliac disease | Gastrointestinal | 15 | 0.545 | [0.409, 0.693] |
| Haemorrhage of rectum and anus | Gastrointestinal | 40 | 0.545 | [0.461, 0.623] |
| Chronic rhinitis | Respiratory | 10 | 0.540 | [0.329, 0.728] |
| Nonspecific abnormal findings in stool content | Gastrointestinal | 50 | 0.534 | [0.454, 0.612] |
| Adhesive capsulitis of shoulder | Musculoskeletal | 11 | 0.531 | [0.319, 0.687] |
| Diseases of tongue | Gastrointestinal | 13 | 0.525 | [0.358, 0.659] |
| Haemangioma | Neoplasms | 10 | 0.512 | [0.346, 0.678] |
| Otitis media | Sense organs | 8 | 0.508 | [0.321, 0.694] |
| Other symptoms and disorders of the soft tissue | Musculoskeletal | 13 | 0.508 | [0.344, 0.688] |
| Skull and face fracture | Musculoskeletal | 10 | 0.507 | [0.327, 0.683] |
| Heartburn | Gastrointestinal | 19 | 0.505 | [0.365, 0.632] |
| Chronic sinusitis | Respiratory | 19 | 0.504 | [0.377, 0.658] |
| Ulceration of the lower GI tract | Gastrointestinal | 13 | 0.500 | [0.316, 0.667] |
| Elevated blood pressure reading without diagnosis | Cardiovascular | 18 | 0.489 | [0.378, 0.605] |
| Other disorders of nose and nasal sinuses | Respiratory | 13 | 0.488 | [0.313, 0.661] |
| Pilar and trichodermal cyst | Dermatological | 14 | 0.460 | [0.328, 0.586] |
| Serous retinal detachment | Sense organs | 8 | 0.460 | [0.294, 0.653] |
| Lipoma of skin subcutaneous tissue | Neoplasms | 20 | 0.454 | [0.330, 0.569] |
| Type 1 diabetes | Endocrine/Metab | 12 | 0.451 | [0.263, 0.653] |
| Dysuria | Genitourinary | 7 | 0.451 | [0.153, 0.743] |
| Visual disturbances | Sense organs | 17 | 0.434 | [0.321, 0.567] |
| Appendicitis | Gastrointestinal | 10 | 0.409 | [0.242, 0.644] |
| Periapical abscess | Gastrointestinal | 5 | 0.406 | [0.115, 0.732] |

**Supplementary Table 7.**
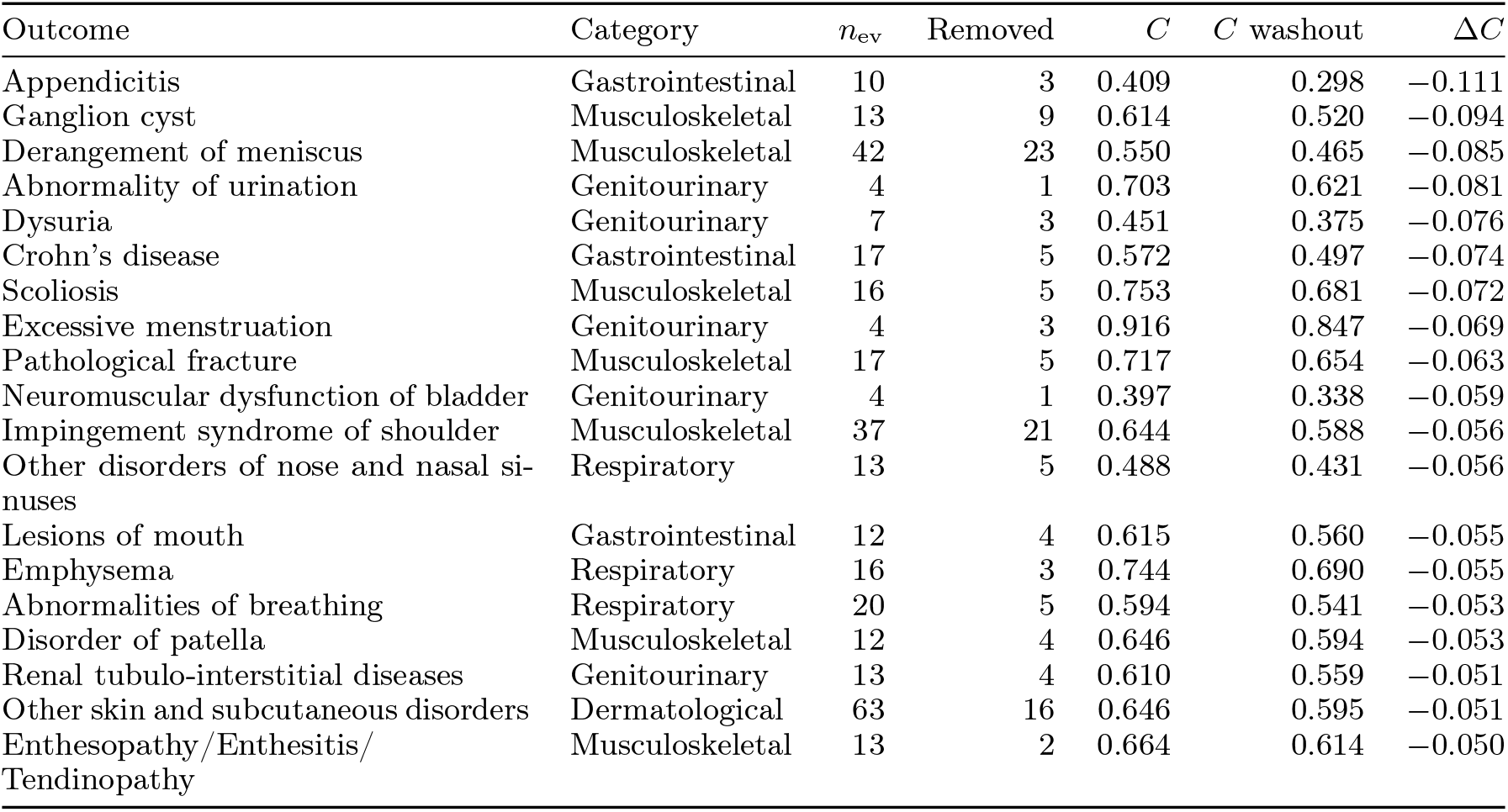
Outcomes with the largest concordance loss under the two-year washout. Of the 388 evaluable outcomes, we show those whose six-year concordance fell by more than 0.05 when every incident case diagnosed within two years of the wrist recording was excluded (the reverse-causality check). Across all 388 outcomes the median change was *−*0.004 and the phenome-wide mean was almost unchanged (0.685 versus 0.688); one outcome, appendicitis, fell by more than 0.10. The larger drops are confined to uncommon conditions: each rests on at most 63 incident test cases, most on fewer than 20, several start from a baseline concordance near chance, and none is a neurodegenerative or cardiometabolic outcome featured in the analysis, so on few events these per-outcome estimates are imprecise. Columns give the incident test cases, the cases removed by the washout, the baseline and washout concordance, and their difference; outcomes are ordered by the size of the drop. Outcomes with fewer than five incident test cases, or fewer than five remaining after the washout, are not shown, and any case count below five is shown as *<* 5, in line with UK Biob<u>ank’s small-numbers policy.</u>

| Outcome | Category | $n_{ev}$ | Removed | $C$ | $C$ washout | $\Delta C$ |
| --- | --- | --- | --- | --- | --- | --- |
| Appendicitis | Gastrointestinal | 10 | $< 5$ | 0.409 | 0.298 | $-0.111$ |
| Derangement of meniscus | Musculoskeletal | 42 | 23 | 0.550 | 0.465 | $-0.085$ |
| Crohn’s disease | Gastrointestinal | 17 | 5 | 0.572 | 0.497 | $-0.074$ |
| Scoliosis | Musculoskeletal | 16 | 5 | 0.753 | 0.681 | $-0.072$ |
| Pathological fracture | Musculoskeletal | 17 | 5 | 0.717 | 0.654 | $-0.063$ |
| Impingement syndrome of shoulder | Musculoskeletal | 37 | 21 | 0.644 | 0.588 | $-0.056$ |
| Other disorders of nose and nasal sinuses | Respiratory | 13 | 5 | 0.488 | 0.431 | $-0.056$ |
| Lesions of mouth | Gastrointestinal | 12 | $< 5$ | 0.615 | 0.560 | $-0.055$ |
| Emphysema | Respiratory | 16 | $< 5$ | 0.744 | 0.690 | $-0.055$ |
| Abnormalities of breathing | Respiratory | 20 | 5 | 0.594 | 0.541 | $-0.053$ |
| Disorder of patella | Musculoskeletal | 12 | $< 5$ | 0.646 | 0.594 | $-0.053$ |
| Renal tubulo-interstitial diseases | Genitourinary | 13 | $< 5$ | 0.610 | 0.559 | $-0.051$ |
| Other skin and subcutaneous disorders | Dermatological | 63 | 16 | 0.646 | 0.595 | $-0.051$ |
| Enthesopathy/Enthesitis/Tendinopathy | Musculoskeletal | 13 | $< 5$ | 0.664 | 0.614 | $-0.050$ |

**Supplementary Table 8.**
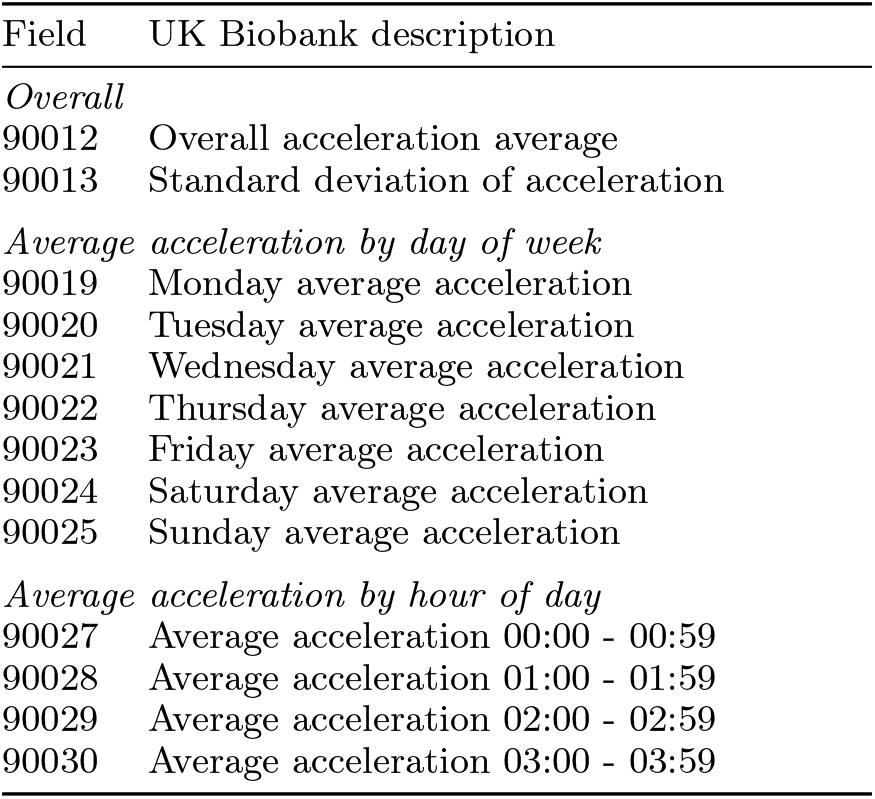
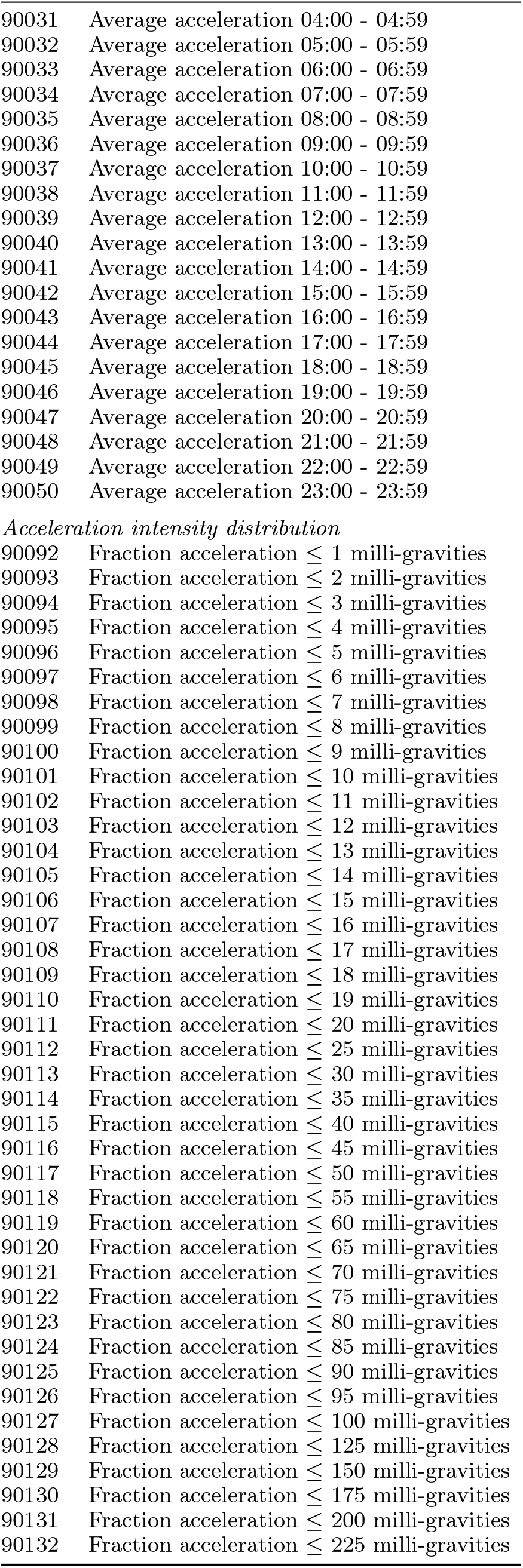
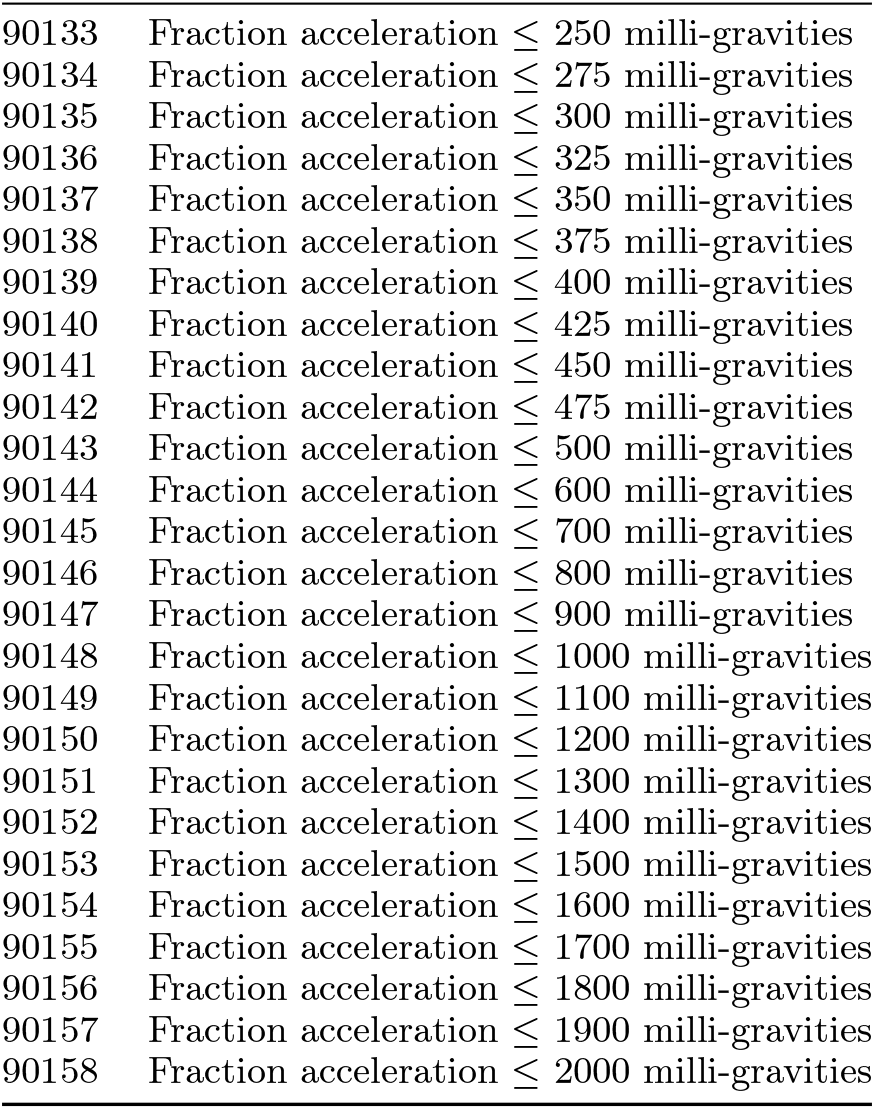
UK Biobank accelerometer summary fields used as the activity baseline. The 100 UK Biobank data-fields that constitute the activity-summary panel compared against the wrist embedding (Figure 3c; Methods). Field identifiers and descriptions are taken verbatim from the UK Biobank Showcase. UK Biobank derives these fields from each participant’s raw wrist recording with the Oxford accelerometer-processing pipeline [13]. Acceleration averages are reported in milli-gravities; each intensity-distribution field is the fraction of time during which acceleration was at or below the stated threshold.

| Field | UK Biobank description |
| --- | --- |
| *Overall* |  |
| 90012 | Overall acceleration average |
| 90013 | Standard deviation of acceleration |
| *Average acceleration by day of week* |  |
| 90019 | Monday average acceleration |
| 90020 | Tuesday average acceleration |
| 90021 | Wednesday average acceleration |
| 90022 | Thursday average acceleration |
| 90023 | Friday average acceleration |
| 90024 | Saturday average acceleration |
| 90025 | Sunday average acceleration |
| *Average acceleration by hour of day* |  |
| 90027 | Average acceleration 00:00 - 00:59 |
| 90028 | Average acceleration 01:00 - 01:59 |
| 90029 | Average acceleration 02:00 - 02:59 |
| 90030 | Average acceleration 03:00 - 03:59 |
| 90031 | Average acceleration 04:00 - 04:59 |
| 90032 | Average acceleration 05:00 - 05:59 |
| 90033 | Average acceleration 06:00 - 06:59 |
| 90034 | Average acceleration 07:00 - 07:59 |
| 90035 | Average acceleration 08:00 - 08:59 |
| 90036 | Average acceleration 09:00 - 09:59 |
| 90037 | Average acceleration 10:00 - 10:59 |
| 90038 | Average acceleration 11:00 - 11:59 |
| 90039 | Average acceleration 12:00 - 12:59 |
| 90040 | Average acceleration 13:00 - 13:59 |
| 90041 | Average acceleration 14:00 - 14:59 |
| 90042 | Average acceleration 15:00 - 15:59 |
| 90043 | Average acceleration 16:00 - 16:59 |
| 90044 | Average acceleration 17:00 - 17:59 |
| 90045 | Average acceleration 18:00 - 18:59 |
| 90046 | Average acceleration 19:00 - 19:59 |
| 90047 | Average acceleration 20:00 - 20:59 |
| 90048 | Average acceleration 21:00 - 21:59 |
| 90049 | Average acceleration 22:00 - 22:59 |
| 90050 | Average acceleration 23:00 - 23:59 |
| *Acceleration intensity distribution* |  |
| 90092 | Fraction acceleration $\leq 1$ milli-gravities |
| 90093 | Fraction acceleration $\leq 2$ milli-gravities |
| 90094 | Fraction acceleration $\leq 3$ milli-gravities |
| 90095 | Fraction acceleration $\leq 4$ milli-gravities |
| 90096 | Fraction acceleration $\leq 5$ milli-gravities |
| 90097 | Fraction acceleration $\leq 6$ milli-gravities |
| 90098 | Fraction acceleration $\leq 7$ milli-gravities |
| 90099 | Fraction acceleration $\leq 8$ milli-gravities |
| 90100 | Fraction acceleration $\leq 9$ milli-gravities |
| 90101 | Fraction acceleration $\leq 10$ milli-gravities |
| 90102 | Fraction acceleration $\leq 11$ milli-gravities |
| 90103 | Fraction acceleration $\leq 12$ milli-gravities |
| 90104 | Fraction acceleration $\leq 13$ milli-gravities |
| 90105 | Fraction acceleration $\leq 14$ milli-gravities |
| 90106 | Fraction acceleration $\leq 15$ milli-gravities |
| 90107 | Fraction acceleration $\leq 16$ milli-gravities |
| 90108 | Fraction acceleration ≤ 17 milli-gravities |
| 90109 | Fraction acceleration ≤ 18 milli-gravities |
| 90110 | Fraction acceleration ≤ 19 milli-gravities |
| 90111 | Fraction acceleration ≤ 20 milli-gravities |
| 90112 | Fraction acceleration ≤ 25 milli-gravities |
| 90113 | Fraction acceleration ≤ 30 milli-gravities |
| 90114 | Fraction acceleration ≤ 35 milli-gravities |
| 90115 | Fraction acceleration ≤ 40 milli-gravities |
| 90116 | Fraction acceleration ≤ 45 milli-gravities |
| 90117 | Fraction acceleration ≤ 50 milli-gravities |
| 90118 | Fraction acceleration ≤ 55 milli-gravities |
| 90119 | Fraction acceleration ≤ 60 milli-gravities |
| 90120 | Fraction acceleration ≤ 65 milli-gravities |
| 90121 | Fraction acceleration ≤ 70 milli-gravities |
| 90122 | Fraction acceleration ≤ 75 milli-gravities |
| 90123 | Fraction acceleration ≤ 80 milli-gravities |
| 90124 | Fraction acceleration ≤ 85 milli-gravities |
| 90125 | Fraction acceleration ≤ 90 milli-gravities |
| 90126 | Fraction acceleration ≤ 95 milli-gravities |
| 90127 | Fraction acceleration ≤ 100 milli-gravities |
| 90128 | Fraction acceleration ≤ 125 milli-gravities |
| 90129 | Fraction acceleration ≤ 150 milli-gravities |
| 90130 | Fraction acceleration ≤ 175 milli-gravities |
| 90131 | Fraction acceleration ≤ 200 milli-gravities |
| 90132 | Fraction acceleration ≤ 225 milli-gravities |
| 90133 | Fraction acceleration ≤ 250 milli-gravities |
| 90134 | Fraction acceleration ≤ 275 milli-gravities |
| 90135 | Fraction acceleration ≤ 300 milli-gravities |
| 90136 | Fraction acceleration ≤ 325 milli-gravities |
| 90137 | Fraction acceleration ≤ 350 milli-gravities |
| 90138 | Fraction acceleration ≤ 375 milli-gravities |
| 90139 | Fraction acceleration ≤ 400 milli-gravities |
| 90140 | Fraction acceleration ≤ 425 milli-gravities |
| 90141 | Fraction acceleration ≤ 450 milli-gravities |
| 90142 | Fraction acceleration ≤ 475 milli-gravities |
| 90143 | Fraction acceleration ≤ 500 milli-gravities |
| 90144 | Fraction acceleration ≤ 600 milli-gravities |
| 90145 | Fraction acceleration ≤ 700 milli-gravities |
| 90146 | Fraction acceleration ≤ 800 milli-gravities |
| 90147 | Fraction acceleration ≤ 900 milli-gravities |
| 90148 | Fraction acceleration ≤ 1000 milli-gravities |
| 90149 | Fraction acceleration ≤ 1100 milli-gravities |
| 90150 | Fraction acceleration ≤ 1200 milli-gravities |
| 90151 | Fraction acceleration ≤ 1300 milli-gravities |
| 90152 | Fraction acceleration ≤ 1400 milli-gravities |
| 90153 | Fraction acceleration ≤ 1500 milli-gravities |
| 90154 | Fraction acceleration ≤ 1600 milli-gravities |
| 90155 | Fraction acceleration ≤ 1700 milli-gravities |
| 90156 | Fraction acceleration ≤ 1800 milli-gravities |
| 90157 | Fraction acceleration ≤ 1900 milli-gravities |
| 90158 | Fraction acceleration ≤ 2000 milli-gravities |

**Supplementary Table 9.**
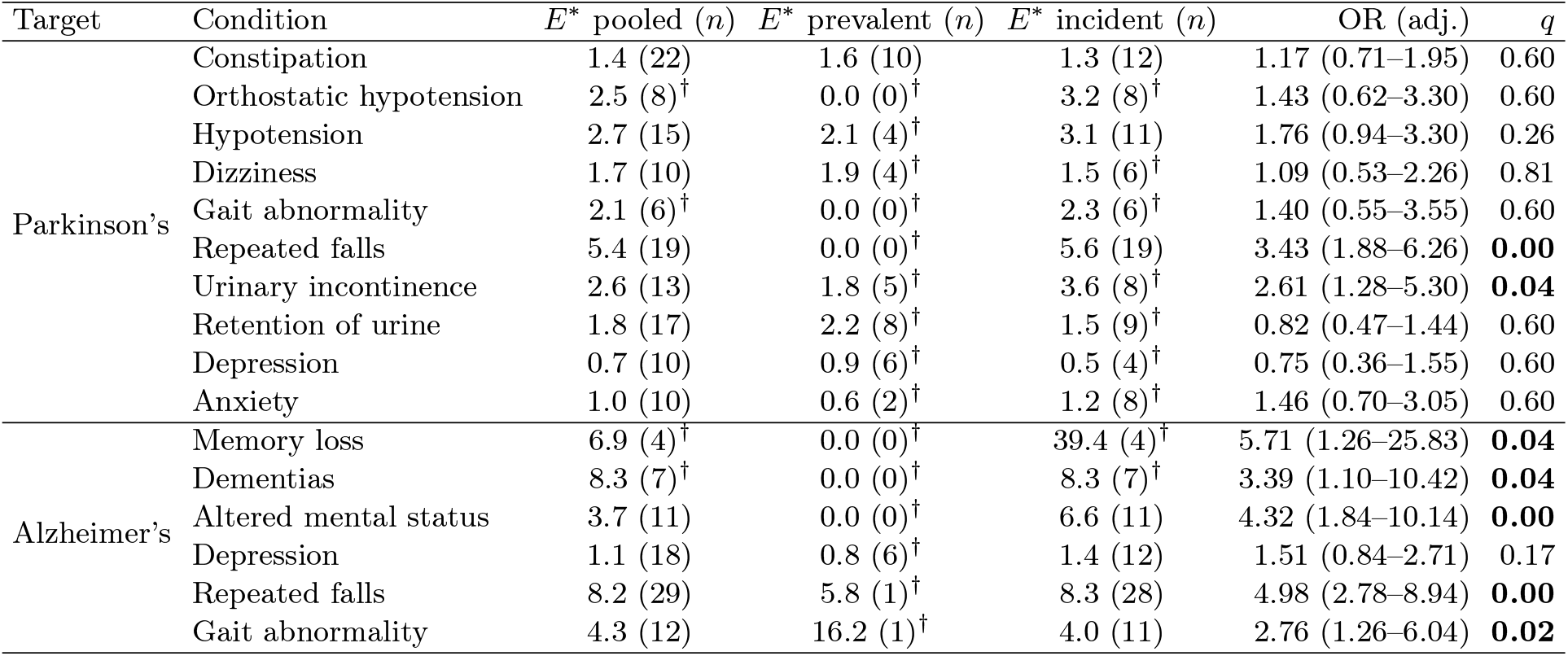
Frailty-adjusted false-positive enrichment for the pre-registered Parkinson’s and Alzheimer’s prodrome, by diagnosis timing. For each pre-registered prodrome condition we report the burden-matched enrichment *E^∗^* (rate among false positives divided by the rate in comorbidity-burden-matched controls) among incident false positives, with the number carrying the condition; a dagger marks under-powered cells (fewer than ten false positives). Almost no false positive carried the condition already at baseline, so the enrichment is essentially all incident. The final columns give the age-, sex-and comorbidity-burden-adjusted logistic odds ratio (over all false positives ever carrying the condition) and its Benjamini–Hochberg false-discovery-rate *q* within each disease family; bold *q* marks survival at *q <* 0.10. Under this adjusted test the Parkinson’s survivors are repeated falls and urinary incontinence, whereas constipation is a well-powered null and orthostatic hypotension is under-powered; the Alzheimer’s survivors are cognitive and motor. Conditions with fewer than five incident false positives are not shown, in line with UK Biobank’s small-numbers policy. European-ancestry test set.

**Supplementary Table 10.** The three neurodegenerative diseases are preferentially confused with one another, beyond the SHARC. For participants whose true diagnosis is Parkinson’s disease, Alzheimer’s disease or dementia (European-ancestry test set; 23, 11 and 23 incident cases, respectively), the mean over-ranking for each disease, expressed as an excess above the cohort median (0 = at the median), on the age-and sex-residualised six-year-risk percentile scale of the misdiagnosis analysis (Methods), shown for each of the three diseases and, as a baseline, averaged over all non-neurological diagnoses (“other systems”). **(a)** Raw. **(b)** After projecting out the SHARC. The within-triad over-ranking stays well above the “other systems” baseline, which falls to zero once the SHARC is removed, so the confusion among the three is specific to them rather than a reflection of the SHARC. Estimates rest on few events.

| True diagnosis | Over-ranked for |  |  | Other systems<br>(baseline) |
| --- | --- | --- | --- | --- |
|  | Parkinson's | Alzheimer's | Dementia |  |
| *(a) Raw (excess above the cohort median)* |  |  |  |  |
| Parkinson's disease | – | +0.28 | +0.30 | +0.11 |
| Alzheimer's disease | +0.18 | – | +0.21 | +0.09 |
| Dementia | +0.14 | +0.16 | – | +0.10 |
| *(b) After removing the SHARC* |  |  |  |  |
| Parkinson's disease | – | +0.24 | +0.31 | –0.01 |
| Alzheimer's disease | +0.13 | – | +0.13 | 0.00 |
| Dementia | +0.08 | +0.09 | – | –0.01 |

**Supplementary Table 11.** High-specificity screening for Parkinson’s disease, incident and prevalent. Area under the curve, partial area under the curve in the high-specificity region (false-positive rate at most 0.10), average precision (AP, the area under the precision–recall curve), and sensitivity at 90, 95 and 99% specificity, for six-year incident screening and for cross-sectional prevalent detection, European ancestry. Average precision reflects precision at the rare-disease base rate: the wrist model reaches an AP of 0.70 (prevalent) and 0.50 (incident) against at most 0.01 for the polygenic score alone. The incident framing has 17 incident Parkinson’s cases in this European-ancestry test set, so these are small-sample fixed-horizon AUROCs and differ from the full-cohort time-dependent AUROC of 0.90 in the main text. In both framings, adding the polygenic score to the wrist model does not raise sensitivity at any operating point, and the polygenic score alone is far below the wrist model. Across all matched diseases, adding the polygenic score to the wrist model raises sensitivity at 90% specificity on average by 0.05 (incident) and 0.10 (prevalent), helping a small number of cardiometabolic and autoimmune outcomes, but it adds nothing at 99% specificity in either framing. “Wrist” is the frozen wrist-embedding six-year hazard; “Clinical” is an age, sex and body-mass-index model; “+ PRS” adds the matched Parkinson’s polygenic score through a leakage-safe out-of-fold fusion (Cox for the incident framing, logistic for prevalent detection).

| Framing | Arm | AUROC | pAUC<sub>FPR≤0.1</sub> | AP | Sens@90% | Sens@95% | Sens@99% |
| --- | --- | --- | --- | --- | --- | --- | --- |
| Incident (6-yr) | Wrist | 0.96 | 0.85 | 0.50 | 0.82 | 0.71 | 0.59 |
|  | Wrist + PRS | 0.96 | 0.84 | 0.48 | 0.76 | 0.71 | 0.53 |
|  | PRS only | 0.48 | 0.47 | 0.00 | 0.00 | 0.00 | 0.00 |
|  | Clinical | 0.88 | 0.65 | 0.03 | 0.59 | 0.29 | 0.06 |
|  | Clinical + PRS | 0.87 | 0.64 | 0.03 | 0.53 | 0.24 | 0.18 |
| Prevalent | Wrist | 0.91 | 0.90 | 0.70 | 0.82 | 0.82 | 0.73 |
|  | Wrist + PRS | 0.90 | 0.90 | 0.69 | 0.82 | 0.82 | 0.73 |
|  | PRS only | 0.67 | 0.56 | 0.01 | 0.18 | 0.18 | 0.18 |
|  | Clinical | 0.65 | 0.49 | 0.00 | 0.09 | 0.00 | 0.00 |
|  | Clinical + PRS | 0.70 | 0.54 | 0.01 | 0.36 | 0.09 | 0.00 |

**Extended Data Figure 1.**
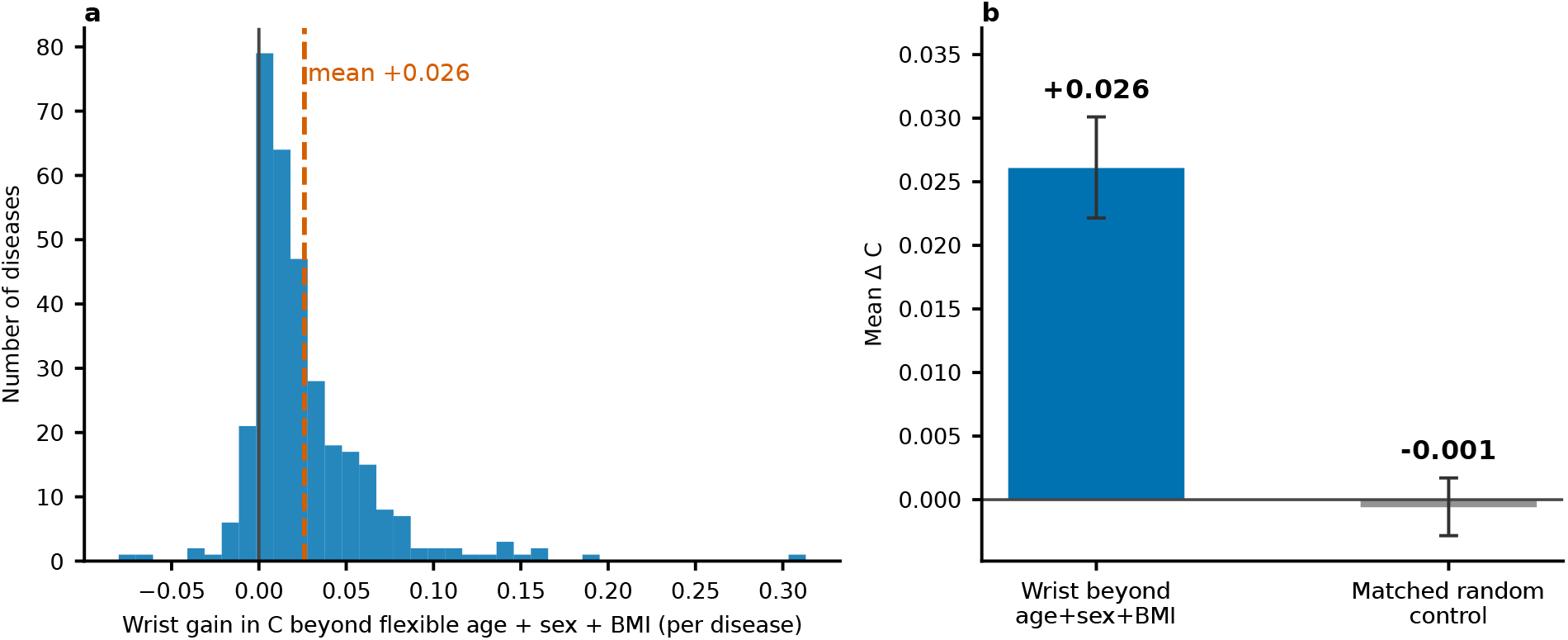
The wrist embedding adds disease signal beyond a flexible demographic model. Per-disease gain in concordance from adding the wrist embedding on top of a flexible age, sex and body-mass-index model, in which age and body-mass index enter with quadratic terms and all pairwise interactions (in contrast to the linear baseline of Figure 3b), mean +0.026 on 87% of the 331 diseases with at least ten incident cases (mortality excluded); a matched, signal-free control column added essentially nothing. Error bars in panel b are 95% confidence intervals from a disease-panel bootstrap.

**Extended Data Figure 2.**
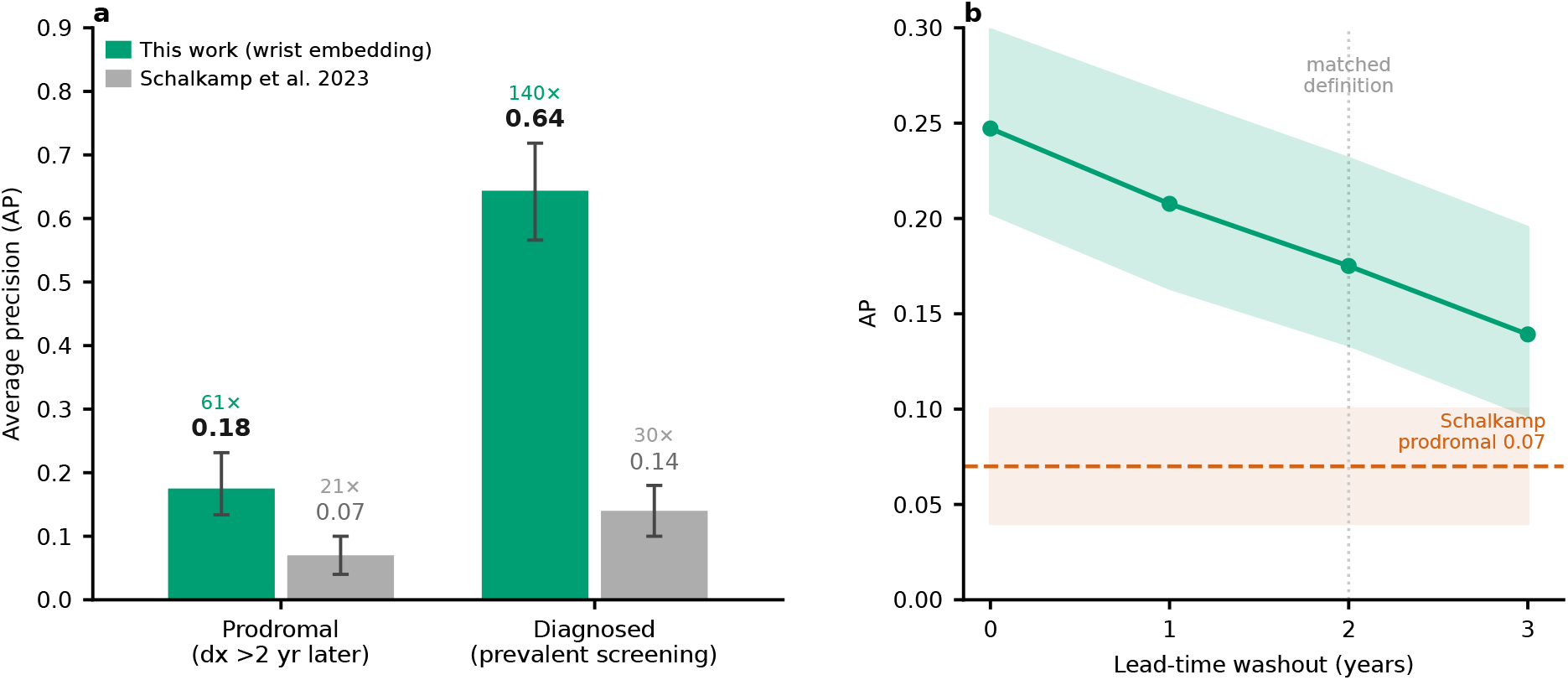
Parkinson’s disease: a general-purpose wrist embedding improves on a bespoke accelerometry model’s precision, for prodromal prediction and for diagnosed screening. a, Average precision (AP) for identifying Parkinson’s disease against the general population in two regimes: green, our wrist-embedding readout (Methods); grey, the accelerometry model of the prior study [8]. In the *prodromal* regime (diagnosed more than two years after the recording) our embedding reaches 0.18 (95% CI 0.13–0.23; 282 cases, prevalence 0.29%) against 0.07 for the prior model (prevalence 0.34%), so our lower base rate makes the comparison conservative. In the *diagnosed* regime (prevalent disease at the time of wear) it reaches 0.64 (95% CI 0.57–0.72; AUROC 0.916; 132 cases) against 0.14, both at the prior study’s diagnosed base rate (0.46%). Numerals are enrichment over the no-skill floor (fold above chance): 61 against 21 for prodromal, and 140 against 30 for diagnosed. Error bars are the 95% CI (ours) or the reported s.d. (prior study). b, Lead-time stability: AP of the prodromal readout as cases diagnosed within one, two and three years of the recording are progressively excluded (green line, 95% CI shaded), against the prior prodromal AP (orange dashed, mean *±* s.d.); the two-year washout (dotted) is the matched definition. Discrimination declines gradually and stays several times the prior value, so it is a genuine pre-diagnostic signature rather than near-onset detection. On the fully held-out test set the same model reaches an AP of 0.41 (95% CI 0.22–0.60) for any incident Parkinson’s disease (25 cases, prevalence 0.48%), essentially unchanged without demographic covariates.

**Extended Data Figure 3.**
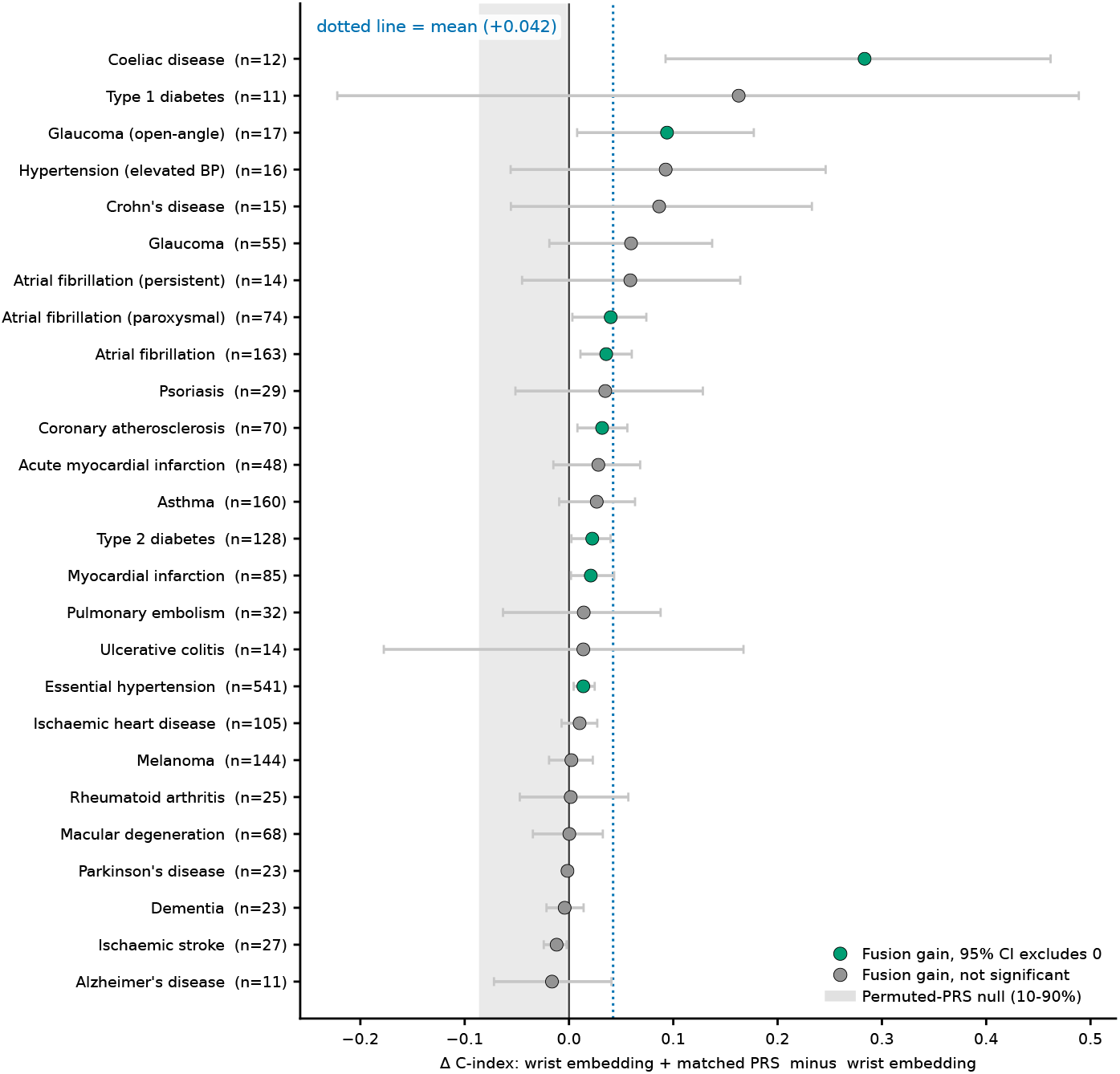
Genetics fusion across all 26 disease-matched outcomes. Change in concordance from adding a disease-matched Standard polygenic risk score to the wrist-embedding hazard, in participants of European genetic ancestry, with a leakage-safe out-of-fold cross-fit on the held-out test set (Methods). Because outcomes are phecodes (a hierarchical clinical coding), some conditions appear as several related phecodes at different levels of specificity, for example atrial fibrillation and its paroxysmal and persistent forms, or ischaemic heart disease, myocardial infarction and coronary atherosclerosis; each is evaluated as a separate outcome with its own incident-case count. Green points mark outcomes whose 95% disease-level bootstrap interval excludes zero; grey points do not reach significance. The grey shaded band is a permuted-PRS control with the same number of covariates (10th to 90th percentile across outcomes); the band lies at or below zero, so genuine gains lie to its right. The dotted line is the mean (+0.042, 95% CI +0.021 to +0.069). Per-disease intervals are wide on these incident-case counts, so the aggregate gain is the robust claim; per-disease Benjamini–Hochberg false-discovery rates are in the supplementary table.

**Extended Data Figure 4.**
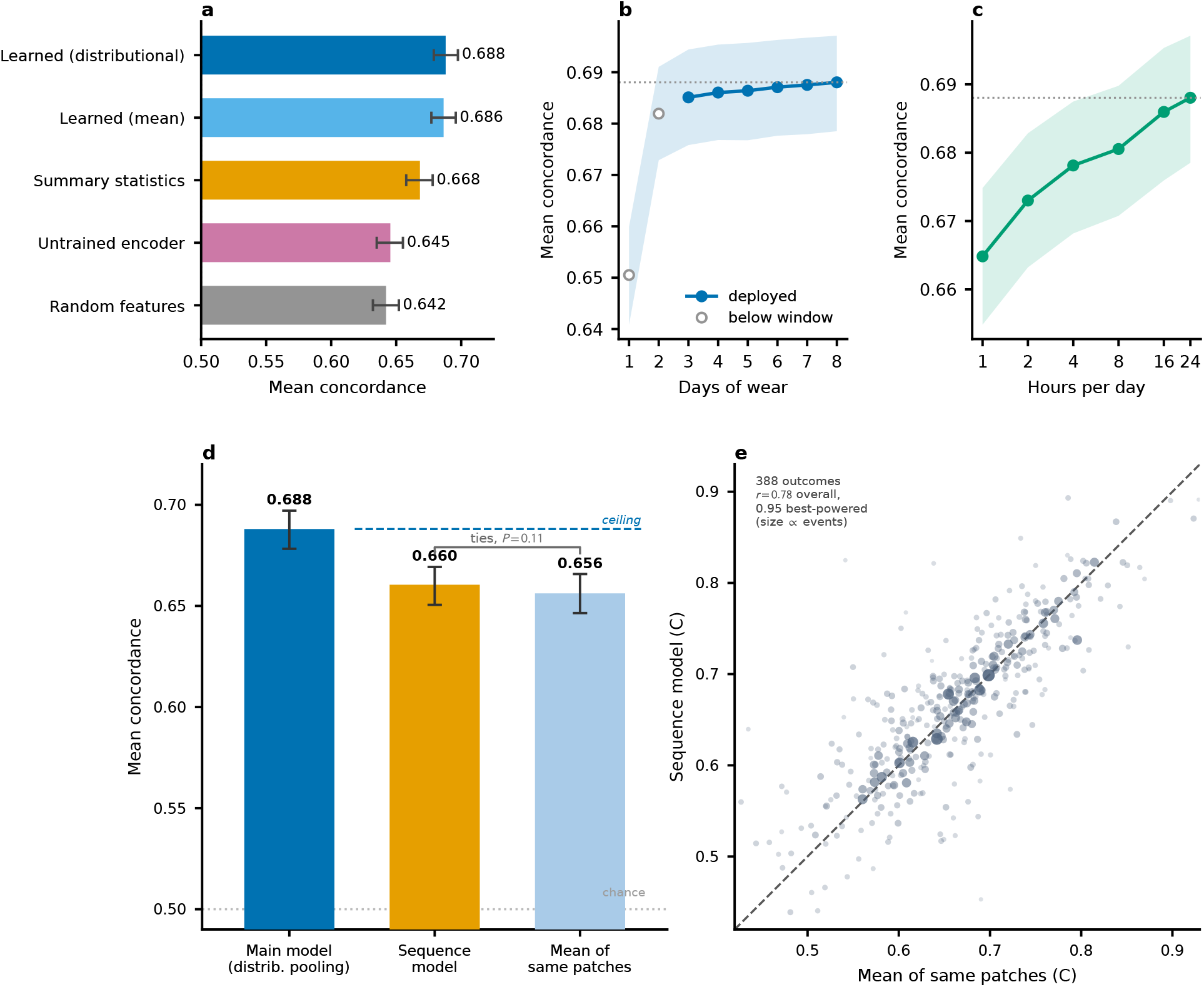
Modelling ablations: the learned representation is necessary, the model needs little input, and an age-pretrained sequence model does not improve on pooling. a, Feature-source ladder: with the survival model, split, covariates and evaluation held fixed, the learned daily embedding is replaced by alternative feature sources of matched dimensionality (Methods). Bars are mean concordance with 95% subject-level bootstrap confidence intervals; the random floor coincides with the age-and-sex baseline retained by the pipeline. b, Days-of-wear titration: with the trained model fixed, each recording is truncated to its first *k* days at inference; filled points are within the deployed three-day window and open points extrapolate below it. c, Hours-per-day titration: each day is re-pooled from only its first *H* hours. d, An age-pretrained sequence model reads the per-patch embeddings of both encoders in time order; it is first pretrained by self-supervision to predict chronological age from input masked in six-hour blocks at 75% (no disease labels are used), then fine-tuned to the 390-outcome panel (Methods). It does no better (0.660) than a simple mean of the same patches (0.656, paired *P* = 0.11); both fall below the main model (0.688, dashed ceiling), whose advantage is the richer daily distributional pooling and the Cox loss, not temporal modelling. Error bars are 95% confidence intervals from a disease-panel bootstrap; the paired *P* = 0.11 is a separate subject-level test. e, Across all 388 outcomes the sequence model’s per-disease concordance tracks a simple mean of the same patches (Pearson *r* = 0.78, rising to 0.95 among the best-powered outcomes); point size is proportional to the number of incident events.

**Extended Data Figure 5.**
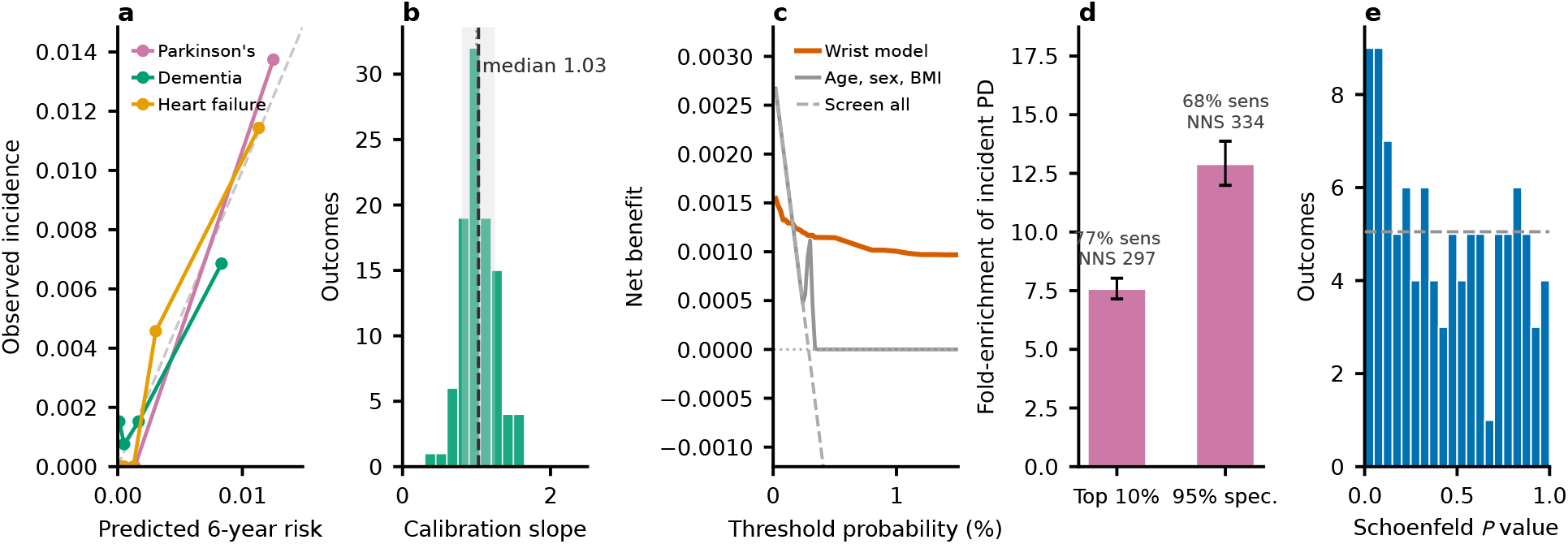
The predicted risks are calibrated, satisfy proportional hazards, and are clinically useful. **a**, Calibration of the six-year predicted risk for Parkinson’s disease, dementia and heart failure: observed incidence (Kaplan-Meier) against predicted risk by quantile bin, with the identity line; the curves show calibration shape and the reported metric is the slope in **b**. **b**, Distribution of calibration slopes across the 101 outcomes with at least 50 incident cases (dark line, median 1.03; shaded band, 0.8 to 1.25; dotted line, 1.0). **c**, Decision-curve analysis for incident Parkinson’s disease: net benefit of the wrist readout against threshold probability, compared with screening everyone, screening no one, and an age, sex and body-mass-index baseline; the baseline’s maximum predicted six-year risk is about 0.4%, above which it flags no one. **d**, Screening yield for prodromal Parkinson’s disease on the unenriched cohort at the top-10% and 95%-specificity operating points: sensitivity, fold-enrichment of the incident rate (bars), and number-needed-to-screen; error bars are 95% confidence intervals from subject-level bootstrap resampling. **e**, Proportional-hazards check: the distribution of scaled Schoenfeld residual test *P* values across the same 101 well-powered outcomes, each entered as the single covariate in a Cox model on the frozen risk score (Grambsch and Therneau, rank-transformed time; Methods). Under proportional hazards these *P* values are uniform, and the dashed line is the count expected per bin. The assumption was rejected for 9 of the 101 outcomes at nominal *P <* 0.05 and for none after Benjamini–Hochberg false-discovery control (about 5 nominal rejections are expected by chance); the nine nominal rejections, which include type 2 diabetes, mostly reflect a risk-score effect that weakens across follow-up, and no flagship outcome violated the assumption after correction.

**Extended Data Figure 6.**
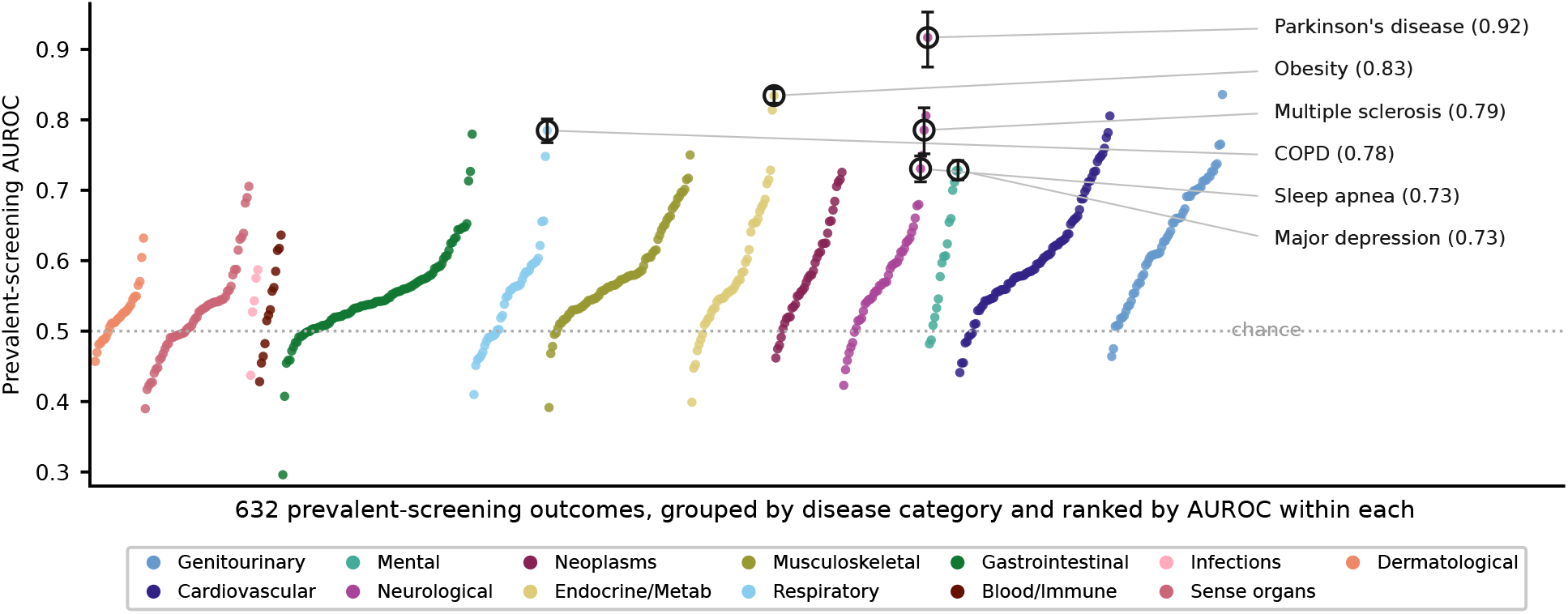
The wrist embedding screens for prevalent disease, led by Parkinson’s disease. Per-disease wrist-embedding prevalent-screening AUROC for all 632 screened outcomes, coloured by disease category and ranked by AUROC within each category; the dotted line marks chance. Six leaders are ringed, with 95% confidence intervals from a 1,000-resample bootstrap over participants (Methods). Prevalent Parkinson’s disease is the strongest single screening signal in the phenome (AUROC 0.916, 95% CI 0.872–0.952), and the ringed leaders span movement, mood and respiratory disease. The three-arm comparison against an age, sex and body-mass-index baseline on the 19 labelled outcomes is reported in the Prevalent-disease screening subsection.

**Extended Data Figure 7.**
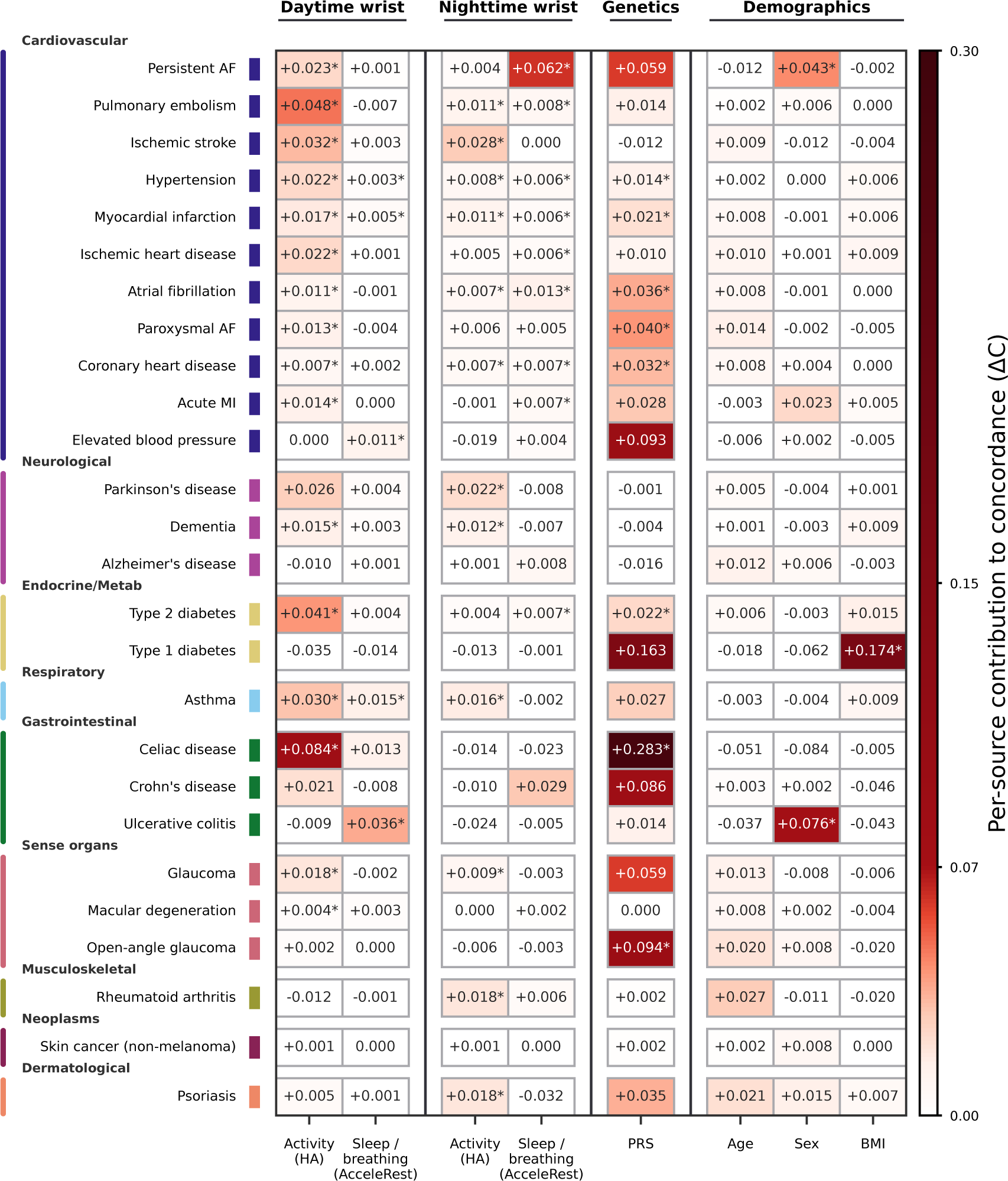
Per-disease source attribution for every outcome with a matched polygenic score. The decomposition of Figure 4 for all 26 outcomes that carry a matched polygenic risk score (rows, grouped by organ system and ordered within each by total wrist contribution), so every cell is populated. Columns and encoding are as in Figure 4. An asterisk marks a statistically significant contribution, and the criterion differs by source: for the wrist components, the subject-level 95% confidence interval excludes zero under Benjamini–Hochberg false-discovery-rate control; for the polygenic score, the 95% confidence interval excludes zero; and for age, sex and body-mass index, the contribution exceeds the 95th percentile of a matched covariate-permutation null. The colour scale matches Figure 4 exactly up to its maximum (a concordance contribution of 0.07, marked on the colour bar) and then continues into darker reds for the larger genetics-driven contributions that appear only in this fuller panel, so a disease shared with Figure 4 is shown in the same colour in both.

**Extended Data Figure 8.**
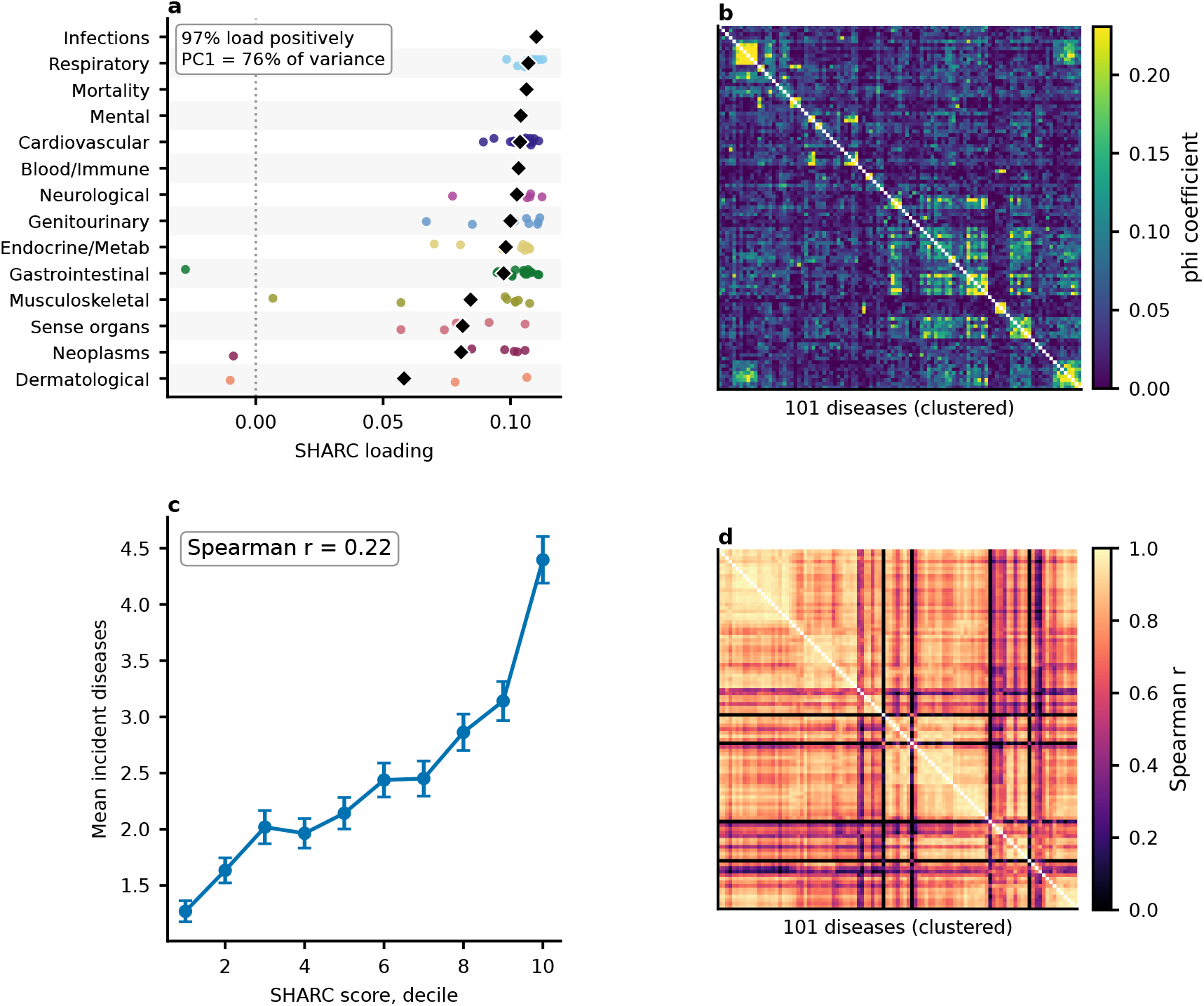
Supplementary anatomy of the Shared Actigraphic Risk Component (SHARC). Companion to Figure 2, for the 101 outcomes with at least 50 incident cases (Methods). (a) SHARC (first-principal-component) loading for every disease, grouped by organ system and ordered by mean loading (black diamonds mark each group’s mean); almost all loadings are positive (97%), so the SHARC is a general risk gradient, with the highest loadings for infectious and respiratory disease and the lowest for skin conditions and neoplasms. (b) Incident co-occurrence (comorbidity, phi coefficient) for the same diseases in the same clustered order as the predicted-risk correlation matrix in panel (d), on a much smaller colour scale (maximum near 0.2 versus 1.0): real co-occurrence is sparse (mean phi 0.045) and does not reproduce the dense predicted-risk correlation. (c) Mean number of incident diseases per participant across deciles of the SHARC score (error bars, *±*1 standard error of the mean); burden rises steadily with the score (Spearman 0.22). (d) Spearman correlation of the age-and sex-residualised predicted six-year risks between every disease pair (mean pairwise correlation 0.69; 0.70 without residualising), for the same 101 outcomes ordered by hierarchical clustering; the colour scale runs from 0 to 1, and the 5.6% of pairs that are weakly negative render at the floor of the scale. This full matrix underlies the per-system variance partition promoted to Figure 2d.

**Extended Data Figure 9.**
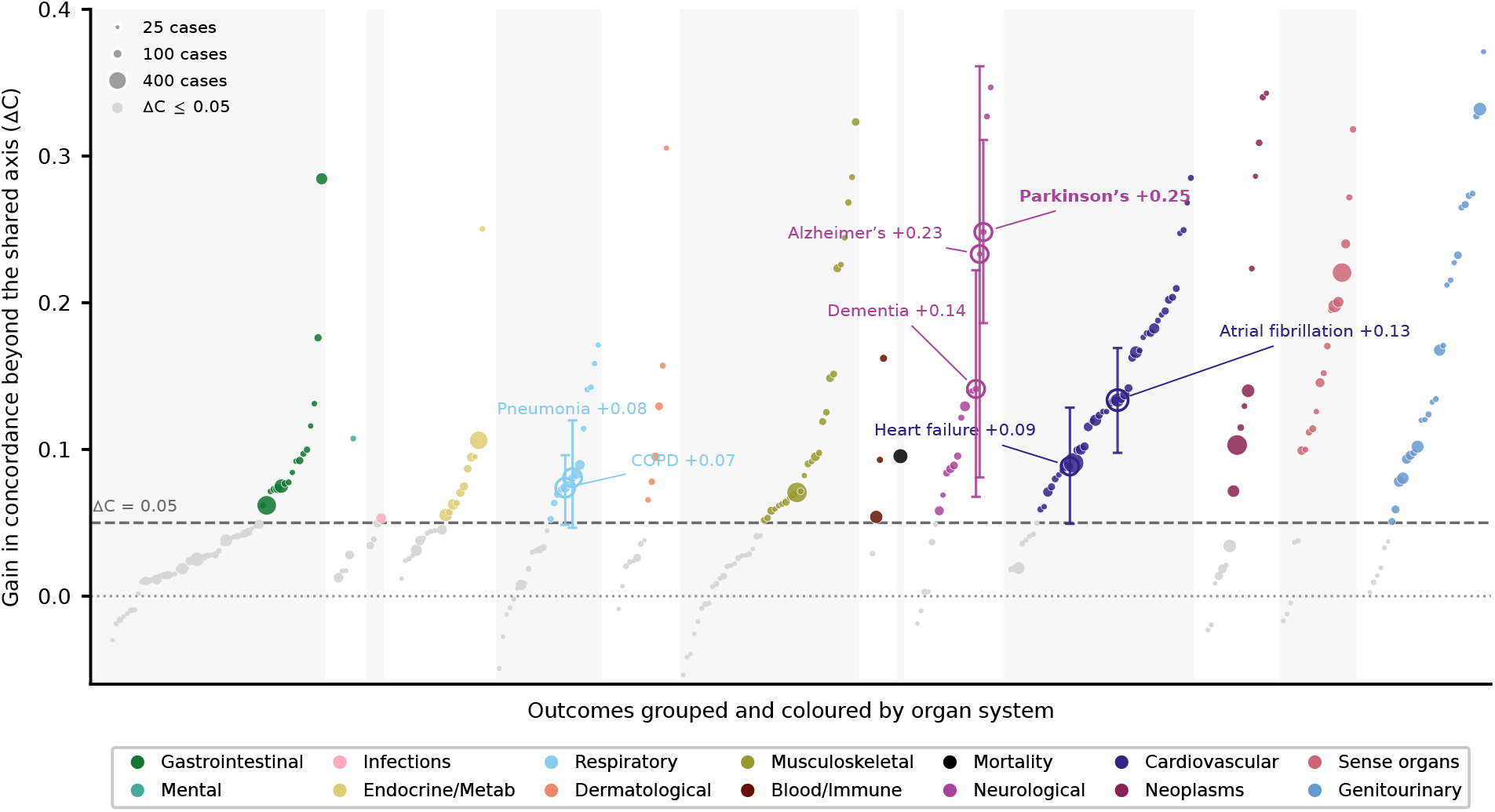
Disease-specific discrimination beyond the SHARC across the phenome. For each of the 332 outcomes with at least 10 incident test cases, the gain in six-year concordance from adding the disease-specific hazard to the SHARC, extending the representative comparison in Figure 2c to the full phenome. Each point is one outcome, positioned along the horizontal axis in organ-system groups and coloured by system; point area scales with the incident-case count, so lower-power outcomes are visibly down-weighted. The dashed line marks a gain of 0.05, and outcomes above it are drawn in full colour with the rest in grey. Of the 332 outcomes, 188 exceed 0.05; among the 101 well-powered outcomes with at least 50 incident cases, 66 do. Parkinson’s disease (+0.25), Alzheimer’s disease (+0.23), dementia (+0.14), atrial fibrillation (+0.13), heart failure (+0.09), pneumonia (+0.08) and chronic obstructive pulmonary disease (+0.07) are labelled. Error bars on the labelled outcomes are 95% confidence intervals from a 300-resample subject-level bootstrap.

**Extended Data Figure 10.**
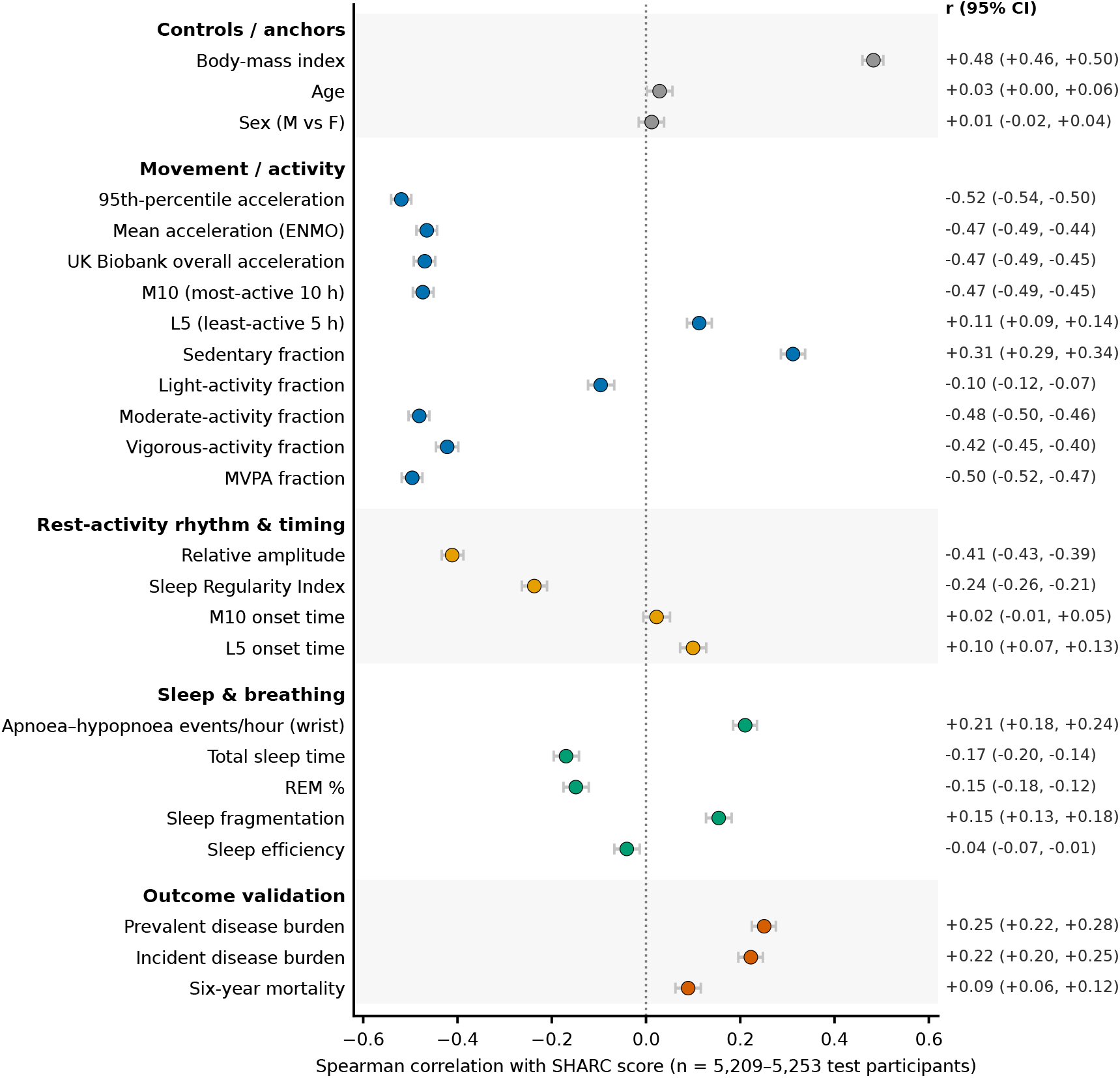
The Shared Actigraphic Risk Component relative to established health descriptors. Each row is the Spearman correlation between the SHARC score and one health descriptor, measured across the held-out test participants (n = 5,209–5,253, depending on missingness). Each point is a correlation and the bars are 95% confidence intervals from 2,000 subject-level percentile bootstrap resamples. Descriptors fall into five groups: control anchors, wrist-derived movement and activity, the rest–activity rhythm and its timing, sleep and breathing, and the reference outcomes the SHARC is built to rank. By construction the SHARC is near-orthogonal to age and sex, and it correlates moderately with body-mass index (*r* = 0.48). It correlates strongly and inversely with the volume and intensity of movement, most strongly with the 95th percentile of acceleration (*r* = *−*0.52), and across mean acceleration, moderate-to-vigorous activity and the UK Biobank overall-acceleration average (*r* between *−*0.47 and *−*0.50), whereas more sedentary time goes with a higher SHARC score (*r* = 0.31). A higher SHARC score also goes with a weaker day–night rest–activity rhythm (relative amplitude *r* = *−*0.41) and more irregular sleep timing (Sleep Regularity Index *r* = *−*0.24), while the clock time of the active and rest windows barely correlates with it. Among the wrist-derived sleep and breathing measures a higher SHARC score goes with more wrist-detected apnoea–hypopnoea events (*r* = 0.21) and more fragmented sleep (*r* = 0.15) and with shorter total sleep time (*r* = *−*0.17) and a lower percentage of REM sleep (*r* = *−*0.15), while sleep efficiency is essentially uncorrelated. As reference anchors, the SHARC rises with prevalent (*r* = 0.25) and incident (*r* = 0.22) disease burden and with six-year mortality (*r* = 0.09). The movement associations survive adjustment for body-mass index (partial Spearman *r* between *−*0.39 and *−*0.46). Smoking, alcohol and physical-activity questionnaires are not available in this data extract.

